# Interventions for treatment of COVID-19: second edition of a living systematic review with meta-analyses and trial sequential analyses (The LIVING Project)

**DOI:** 10.1101/2020.11.22.20236448

**Authors:** Sophie Juul, Emil Eik Nielsen, Joshua Feinberg, Faiza Siddiqui, Caroline Kamp Jørgensen, Emily Barot, Johan Holgersson, Niklas Nielsen, Peter Bentzer, Areti Angeliki Veroniki, Lehana Thabane, Fanlong Bu, Sarah Klingenberg, Christian Gluud, Janus Christian Jakobsen

**Author notes:** Contact information: Emil Eik Nielsen Joshua Feinberg Faiza Siddiqui Caroline Kamp Jørgensen Emily Barot Johan Holgersson Niklas Nielsen Peter Bentzer Areti Angeliki Veroniki Lehana Thabane Fanlong Bu Sarah Klingenberg Christian Gluud Janus Christian Jakobsen.

## Abstract

**Background:** COVID-19 is a rapidly spreading disease that has caused extensive burden to individuals, families, countries, and the world. Effective treatments of COVID-19 are urgently needed. This is the second edition of a living systematic review of randomized clinical trials assessing the effects of all treatment interventions for participants in all age groups with COVID-19.

**Methods and findings:** We planned to conduct aggregate data meta-analyses, trial sequential analyses, network meta-analysis, and individual patient data meta-analyses. Our systematic review was based on PRISMA and Cochrane guidelines, and our eight-step procedure for better validation of clinical significance of meta-analysis results. We performed both fixed-effect and random-effects meta-analyses. Primary outcomes were all-cause mortality and serious adverse events. Secondary outcomes were admission to intensive care, mechanical ventilation, renal replacement therapy, quality of life, and non-serious adverse events. According to the number of outcome comparisons, we adjusted our threshold for significance to *p* = 0.033. We used GRADE to assess the certainty of evidence. We searched relevant databases and websites for published and unpublished trials until November 2, 2020. Two reviewers independently extracted data and assessed trial methodology.

We included 82 randomized clinical trials enrolling a total of 40,249 participants. 81 out of 82 trials were at overall high risk of bias.

Meta-analyses showed no evidence of a difference between corticosteroids versus control on all-cause mortality (risk ratio [RR] 0.89; 95% confidence interval [CI] 0.79 to 1.00; *p* = 0.05; I^2^ = 23.1%; eight trials; very low certainty), on serious adverse events (RR 0.89; 95% CI 0.80 to 0.99; *p* = 0.04; I^2^ = 39.1%; eight trials; very low certainty), and on mechanical ventilation (RR 0.86; 95% CI 0.55 to 1.33; *p* = 0.49; I^2^ = 55.3%; two trials; very low certainty). The fixed-effect meta-analyses showed indications of beneficial effects. Trial sequential analyses showed that the required information size for all three analyses was not reached.

Meta-analysis (RR 0.93; 95% CI 0.82 to 1.07; *p* = 0.31; I^2^ = 0%; four trials; moderate certainty) and trial sequential analysis (boundary for futility crossed) showed that we could reject that remdesivir versus control reduced the risk of death by 20%. Meta-analysis (RR 0.82; 95% CI 0.68 to 1.00; *p* = 0.05; I^2^ = 38.9%; four trials; very low certainty) and trial sequential analysis (required information size not reached) showed no evidence of difference between remdesivir versus control on serious adverse events. Fixed-effect meta-analysis showed indications of a beneficial effect of remdesivir on serious adverse events.

Meta-analysis (RR 0.40; 95% CI 0.19 to 0.87; *p* = 0.02; I^2^ = 0%; two trials; very low certainty) showed evidence of a beneficial effect of intravenous immunoglobulin versus control on all-cause mortality, but trial sequential analysis (required information size not reached) showed that the result was severely underpowered to confirm or reject realistic intervention effects.

Meta-analysis (RR 0.63; 95% CI 0.35 to 1.14; *p* = 0.12; I^2^ = 77.4%; five trials; very low certainty) and trial sequential analysis (required information size not reached) showed no evidence of a difference between tocilizumab versus control on serious adverse events. Fixed-effect meta-analysis showed indications of a beneficial effect of tocilizumab on serious adverse events. Meta-analysis (RR 0.70; 95% CI 0.51 to 0.96; *p* = 0.02; I^2^ = 0%; three trials; very low certainty) showed evidence of a beneficial effect of tocilizumab versus control on mechanical ventilation, but trial sequential analysis (required information size not reached) showed that the result was severely underpowered to confirm of reject realistic intervention effects.

Meta-analysis (RR 0.32; 95% CI 0.15 to 0.69; *p* < 0.00; I^2^ = 0%; two trials; very low certainty) showed evidence of a beneficial effect of bromhexidine versus standard care on non-serious adverse events, but trial sequential analysis (required information size not reached) showed that the result was severely underpowered to confirm or reject realistic intervention effects.

Meta-analyses and trial sequential analyses (boundary for futility crossed) showed that we could reject that hydroxychloroquine versus control reduced the risk of death and serious adverse events by 20%.

Meta-analyses and trial sequential analyses (boundary for futility crossed) showed that we could reject that lopinavir-ritonavir versus control reduced the risk of death, serious adverse events, and mechanical ventilation by 20%.

All remaining outcome comparisons showed that we did not have enough information to confirm or reject realistic intervention effects. Nine single trials showed statistically significant results on our outcomes, but were underpowered to confirm or reject realistic intervention effects. Due to lack of data, it was not relevant to perform network meta-analysis or possible to perform individual patient data meta-analyses.

**Conclusions:** No evidence-based treatment for COVID-19 currently exists. Very low certainty evidence indicates that corticosteroids might reduce the risk of death, serious adverse events, and mechanical ventilation; that remdesivir might reduce the risk of serious adverse events; that intraveneous immunoglobin might reduce the risk of death and serious adverse events; that tocilizumab might reduce the risk of serious adverse events and mechanical ventilation; and that bromhexidine might reduce the risk of non-serious adverse events. More trials with low risks of bias and random errors are urgently needed. This review will continuously inform best practice in treatment and clinical research of COVID-19.

**Systematic review registration** PROSPERO CRD42020178787

**Author summary:** **Why was this study done?**

- Severe acute respiratory syndrome coronavirus 2 (SARS-CoV-2) infection has spread rapidly worldwide, causing an international outbreak of the corona virus disease 2019 (COVID-19).
- There is a need for a living systematic review evaluating the beneficial and harmful effects of all possible interventions for treatment of COVID-19.

**What did the researchers do and find?**

- We conducted the second edition of our living systematic review with meta-analyses and Trial sequential analyses to compare the effects of all treatment interventions for COVID-19.
- Very low certainty evidence indicated that corticosteroids might reduce the risk of death, serious adverse events, and mechanical ventilation; that remdesivir might reduce the risk of serious adverse events; that intraveneous immunoglobin might reduce the risk of death and serious adverse events; that tocilizumab might reduce the risk of serious adverse events and mechanical ventilation; and that bromhexidine might reduce the risk of non-serious adverse events.
- Nine single trials showed statistically significant results on our predefined outcomes but were underpowered to confirm or reject realistic intervention effects.
- None of the remaining trials showed evidence of a difference of the experimental interventions on our predefined outcomes.

**What do these findings mean?**

- No evidence-based treatment for COVID-19 currently exists
- More high quality, low risk of bias randomized clinical trials are urgently needed.

## Introduction

In 2019, a novel coronavirus named severe acute respiratory syndrome coronavirus 2 (SARS-CoV-2) caused an international outbreak of the respiratory illness COVID-19 [1]. Since the initial outbreak in China, SARS-CoV-2 has spread globally and COVID-19 is labeled a public health emergency of global concern by the World Health Organization [2]. The full spectrum of COVID-19 ranges from subclinical infection over mild, self-limiting respiratory tract illness to severe progressive pneumonia, multiorgan failure, and death [3]. Severe disease onset might result in death due to massive alveolar damage and progressive respiratory failure [4–6].

Many randomized clinical trials assessing the effects of different potential treatments for COVID-19 are currently underway. However, a single trial can rarely validly assess the effects of any intervention and there is an urgent need to continuously surveil and update the aggregated evidence base so that effective interventions, if such exist, are implemented in clinical practice [7].

The present living systematic review with aggregate meta-analyses and trial sequential analyses aims to continuously inform evidence-based guideline recommendations for the treatment of COVID-19, taking risks of systematic errors (‘bias’), risks of random errors (‘play of chance’), and certainty of the findings into consideration [8].

## Methods

We report this systematic review based on the Preferred Reporting Items for Systematic Reviews and Meta-Analysis (PRISMA) guidelines (**S1 Text**) [9, 10]. The updated methodology used in this living systematic review is according to the Cochrane Handbook of Systematic Reviews of Interventions [11] and described in our protocol [8], which was registered in the PROSPERO database (ID: CRD42020178787) prior to the systematic literature search.

### Search strategy and selection criteria

#### Electronic searches

An information specialist searched the Cochrane Central Register of Controlled Trials (CENTRAL) in The Cochrane Library, Medical Literature Analysis and Retrieval System Online (MEDLINE Ovid), Excerpta Medica database (Embase Ovid), Latin American and Caribbean Health Sciences Literature (LILACS; Bireme), Science Citation Index Expanded (SCI-EXPANDED; Web of Science), Conference Proceedings Citation Index – Science (CPCI-S; Web of Science), BIOSIS (Web of Science), CINAHL (EBSCO host), Chinese Biomedical Literature Database (CBM), China Network Knowledge Information (CNKI), Chinese Science Journal Database (VIP), and Wanfang Database to identify relevant trials. We searched all databases from their inception and until November 2, 2020. Trials were included irrespective of language, publication status, publication year, and publication type. For the detailed search strategies for all electronic searches, see **S2 Text.**

#### Searching other resources

The reference lists of relevant trial publications were checked for any unidentified randomized clinical trials. To identify unpublished trials, we searched clinical trial registries (e.g. clinicaltrials.gov, clinicaltrialregister.eu, who.int/ictrp, chictr.org.cn) of Europe, USA, and China, and websites of pharmaceutical companies and of U.S. Food and Drug Administration (FDA) and European Medicines Agency (EMA). We also searched the COVID-19 Study Registry [12] and the real-time dashboard of randomized trials [13].

We included unpublished and grey literature trials and assessed relevant retraction statements and errata for included trials. We also searched preprint servers (bioRxiv, medRxiv) for unpublished trials. We contacted all corresponding authors to obtain individual patient data.

### Living systematic review

In this living systematic review, two independent investigators receive a weekly updated literature search file, and continuously include relevant newly published or unpublished trials. The relevant meta-analyses, trial sequential analyses, and network meta-analysis will be continuously updated, and if new evidence is available (judged by the author group), the results will be submitted for publication. Every month, the author group will discuss whether searching once a week is necessary. For a detailed overview of the living systematic review work flow, see our protocol [8]. As this is a living systematic review analyzing results of randomized clinical trials, no ethical approval is required.

### Data extraction

Two authors (EEN and JF) independently screened relevant trials. Seven authors in pairs (SJ, EEN, JF, FS, CKJ, EB, JH) independently extracted data using a standardized data extraction sheet. Any discrepancies were resolved through discussion, or if required, through discussion with a third author (JCJ). We contacted corresponding authors if relevant data were unclear or missing.

### Risk of bias assessment

Risk of bias was assessed with the Cochrane Risk of Bias tool – version 2 (RoB 2) [11, 14]. Seven authors in pairs (SJ, EEN, JF, FS, CKJ, EB, JH) independently assessed risk of bias. Any discrepancies were resolved through discussion or, if required, through discussion with a third author (JCJ). Bias was assessed with the following domains: bias arising from the randomization process, bias due to deviations from the intended interventions, bias due to missing outcome data, bias in measurement of outcomes, and bias arising from selective reporting of results [11, 14]. We contacted corresponding authors of trials with unclear or missing data.

### Outcomes and subgroup analyses

Primary and secondary outcomes were predefined in our protocol [8]. Primary outcomes were all-cause mortality and serious adverse events (as defined by the ICH-GCP guidelines) [8, 15]. Secondary outcomes were admission to intensive care (as defined by trialists), mechanical ventilation (as defined by trialists), renal replacement therapy (as defined by trialists), quality of life, and non-serious adverse events. We classified non-serious adverse events as any adverse event not assessed as serious according to the ICH-GCP definition.

We chose to add time to clinical improvement as a post hoc outcome. We planned several subgroup analyses, which are described in detail in our protocol [8]. For all outcomes, we used the trial results reported at maximum follow-up.

### Assessment of statistical and clinical significance

We performed our aggregate data meta-analyses according to Cochrane [11], Keus et al. [16], and the eight-step assessment by Jakobsen et al. [17] for better validation of meta-analytic results in systematic reviews. Review Manager version 5.4 [18] and Stata 16 (StataCorp LLC, College Station, TX, USA) [19] were used for all statistical analyses. We used risk ratios (RR) for dichotomous outcomes. We assessed a total of two primary outcomes per comparison, and we therefore adjusted our thresholds for significance [17] and considered a *p*-value of 0.033 or less as the threshold for statistical significance [8, 17]. Because we primarily considered results of secondary outcomes as hypothesis generating, we did not adjust the *p*-value for secondary outcomes. We conducted both random-effects (DerSimonian-Laird) and fixed-effect (Mantel-Haenszel) meta-analyses for all analyses and chose the most conservative result as our primary result [11,17,20,21]. We used trial sequential analysis to control for random errors [22–30]. Trial sequential analysis estimates the diversity-adjusted required information size (DARIS), which is the number of participants needed in a meta-analysis to detect or reject a certain intervention effect. Statistical heterogeneity was quantified by calculating heterogeneity (I^2^) for traditional meta-analyses and diversity (D^2^) for trial sequential analysis. We used Grading Recommendations Assessment Development Evaluation (GRADE) to assess the certainty of evidence. We downgraded imprecision in GRADE by two levels if the accrued number of participants were below 50% of the DARIS, and one level if between 50% and 100% of DARIS [17]. We did not downgrade if benefit, harm, futility or DARIS were reached. We used Fisher’s exact test to calculate *p*-values for all single trial results.

## Results

### Study characteristics

On November 2, 2020 our literature searches identified 15,359 records after duplicates were removed. We included a total of 82 clinical trials randomizing 40,249 participants (**Fig 1**) [31–114]. We identified several trials including participants *suspected* of COVID-19 [115, 116]. None of the trials reported separate data on COVID-19 positive participants compared to the remaining participants. We included trials if approximately 50% or more participants had a confirmed COVID-19 diagnosis. We wrote to all authors requesting separate data on COVID-19 confirmed participants, but we have received no responses yet. For at detailed overview of excluded trials, see **S1 Table.**

**Fig 1.**
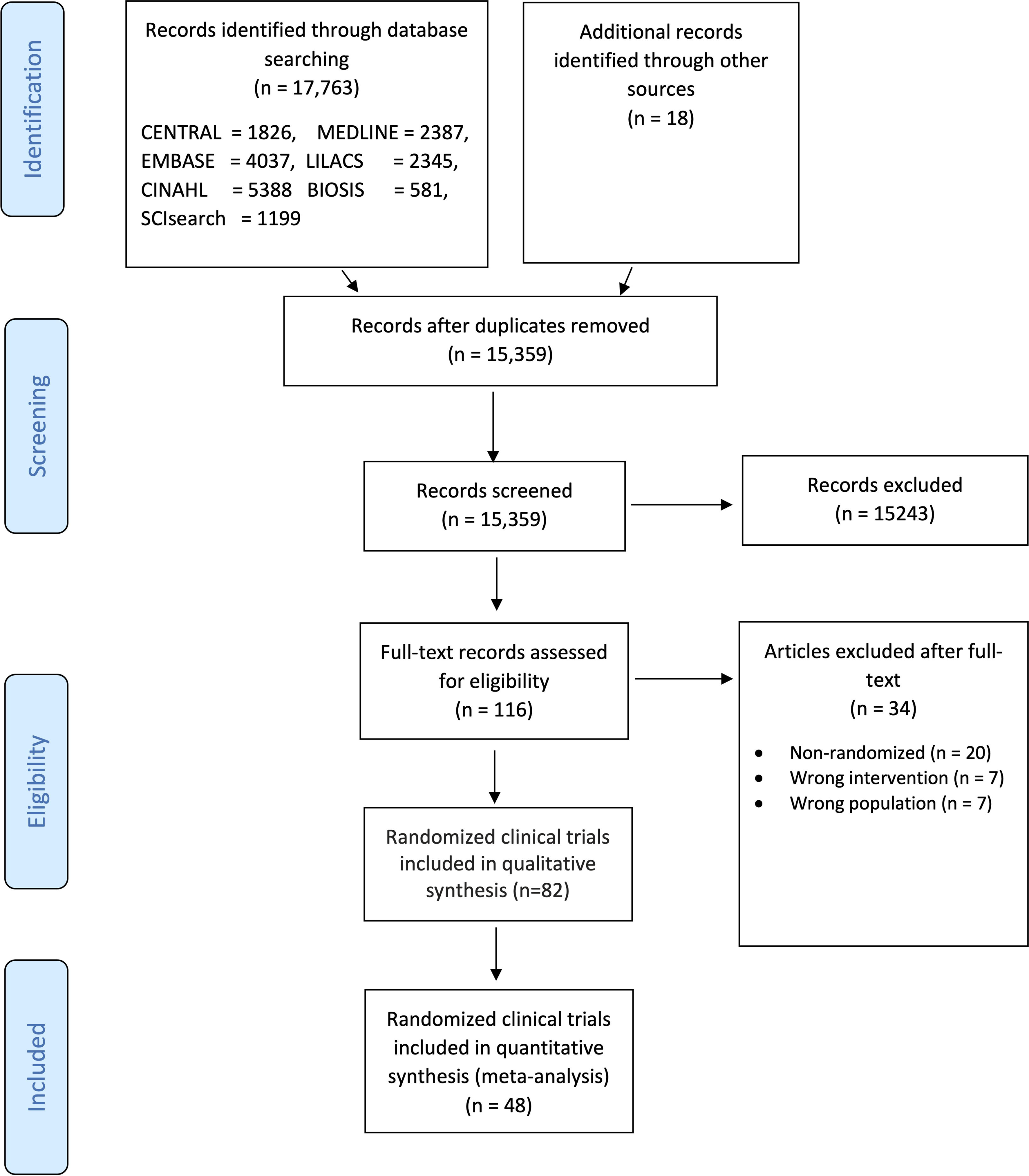
PRISMA flow diagram

Characteristics of included trials and the trial results can be found in **S2 Table**. Most trials were at high risk of bias (**S3 Table**).

The identified trials compared the following interventions: 10 trials compared corticosteroids versus standard care [52,56,87,89,96–99] or placebo [68, 88]; four trials compared remdesivir versus standard care [86, 110] or placebo [43, 65]; 13 trials compared hydroxychloroquine versus standard care [34,35,42,48,54,55,58,59,105,110], or placebo [53, 108]; five trials compared lopinavir-ritonavir versus standard care [32,40,106,110] or a co-intervention alone [45]; two trials compared interferon beta-1a versus standard care [36, 110]; four trials compared convalescent plasma versus standard care [39,51,78,91]; three trials compared azithromycin versus standard care [83] or co-interventions with standard care [54] or without standard care [82]; three trials compared colchicine versus standard care [49], placebo plus standard care [92], or placebo plus a co-intervention [107]; two trials compared immunoglobulin versus standard care [57] or placebo [95]; six trials compared tocilizumab versus standard care [93,111–113], placebo with standard care [90] or favipiravir alone as co-intervention [114];two trials compared bromhexine versus standard care [94, 104]; and three trials compared favipiravir versus standard care [41, 67] or a co-intervention alone [114].

The remaining trial comparisons included: favipiravir versus umifenovir [33]; umifenovir versus lopinavir-ritonavir [40]; umifenovir versus standard care [40]; novaferon versus novaferon plus lopinavir-ritonavir [45]; novaferon plus lopinavir-ritonavir versus lopinavir-ritonavir [45]; novaferon versus lopinavir-ritonavir [45]; alpha lipotic acid versus placebo [46]; baloxavir marboxil versus favipavir [41]; baloxavir marboxil versus standard care [41]; triple combination of interferon beta-1b plus lopinavir-ritonavir plus ribavirin versus lopinavir-ritonavir [38]; remdesivir for 5 days versus remdesivir for 10 days [37]; high-flow nasal oxygenation versus standard bag-valve oxygenation [44]; hydroxychloroquine versus chloroquine [48]; chloroquine versus standard care [48]; high dosage chloroquine diphosphate versus low dosage chloroquine diphosphate [50]; hydroxychloroquine plus azithromycin versus standard care [54]; triple combination of darunavir plus cobicistat plus interferon alpha-2b versus interferon alpha-2b [117]; lopinavir-ritonavir plus interferon alpha versus ribavirin plus interferon alpha [61]; ribavirin plus lopinavir-ritonavir plus interferon alpha versus ribavirin plus interferon alpha [61]; ribavirin plus lopinavir-ritonavir plus interferon alpha versus lopinavir-ritonavir plus interferon alpha [61]; lincomycin versus azithromycin [62];, 99-mTc-methyl diphosphonate (99mTc-MDP) injection versus standard care [63]; interferon alpha-2b plus interferon gamma versus interferon alpha-2b [64]; telmisartan versus standard care [66]; avifavir 1800/800 versus avifavir 1600/600 [67]; dexamethasone plus aprepitant versus dexamethasone [69]; anti-C5a antibody versus standard care [70]; azvudine versus standard care [73]; human plasma-derived C1 esterase/kallikrein inhibitor versus standard care [72]; icatibant acetate versus standard care [72]; icatibant acetate versus human plasma-derived C1 esterase/kallikrein inhibitor [72]; pulmonary rehabilitation program versus isolation at home [71]; auxora (calcium release-activated calcium channel inhibitors) versus standard care [74]; umbilical cord stem cell infusion versus standard care [75]; vitamin C versus placebo [76]; sofosbuvir plus daclatasvir versus standard care [80]; sofosbuvir plus daclatasvir plus ribavirin versus hydroxychloroquine plus lopinavir-ritonavir with or without ribavirin [79]; interferon beta-1b versus standard care [81]; calcifediol versus standard care [84]; recombinant human granulocyte colony–stimulating factor versus standard care [85]; intravenous and/or nebulized electrolyzed saline with dose escalation versus standard care [100]; nasal irrigation with hypertonic saline plus surfactant versus no intervention [101]; nasal irrigation with hypertonic saline plus surfactant versus nasal irrigation with hypertonic saline [101]; nasal irrigation with hypertonic saline versus no intervention [101]; triazavirin versus placebo [102]; N-acetylcysteine versus placebo [103]; tocilizumab versus favipiravir [114].

The maximum follow-up time ranged from five [34, 35] to 60 days [90, 99] after randomization. For several of our outcomes it was not possible to conduct meta-analysis due to insufficient data.

### Corticosteroids versus control

We identified 10 trials (11 comparisons) randomizing 7,918 participants to corticosteroids versus standard care [52,56,87,89,96–99] or placebo [68, 88]. One trial was assessed at low risk of bias [88]. The remaining trials were assessed at high risk of bias (**S3 Table**). Five trials assessed the effects of methylprednisolone [56,68,96–98], three trials assessed the effects of dexamethasone [52,87,99], and two trials (three comparisons) assessed the effects of hydrocortisone [88, 89]. One trial assessing the effects of methylprednisolone was not eligible for meta-analysis, as approximately half of the participants in the experimental group were non-randomized [56]. We contacted the trial authors and asked for separate data for all randomized participants, but did not receive any response. Another trial assessing the effects of methylprednisolone was not eligible for meta-analysis, as the trial did not report on any of our review outcomes [96]. We requested data for our review outcomes from the trial authors but did not receive a response.

#### Meta-analysis of all-cause mortality

Random-effects meta-analysis showed no evidence of a difference between corticosteroids and control on all-cause mortality (RR 0.89; 95% CI 0.79 to 1.00; *p* = 0.05; I^2^ = 23.1%; eight trials; very low certainty) (**Fig 2, S4 Table**). Fixed-effect meta-analysis showed evidence of a beneficial effect of corticosteroids versus control on all-cause mortality (RR 0.88; 95% CI 0.82 to 0.95; *p* = 0.00; I^2^ = 23.6%, eight trials) (**S1 Fig**). Visual inspection of the forest plot and measures to quantify heterogeneity (I^2^ = 23.1%) indicated no substantial heterogeneity. The time-points of assessment varied from 21 [88] to 30 days after randomization [97, 118]. The trial sequential analysis showed that we did not have enough information to confirm or reject that corticosteroids versus control reduce the risk of all-cause mortality with a relative risk reduction of 20% (**Fig 3**). The subgroup analysis assessing the effects of the different corticosteroids versus control showed no significant subgroup differences (*p* = 0.57) (**Fig 2**). The subgroup analysis assessing the effects of disease severity as defined by trialists (mild, moderate, severe, or a combination) showed no significant subgroup differences (*p* = 0.42) (**S2 Fig**).

**Fig 2.**
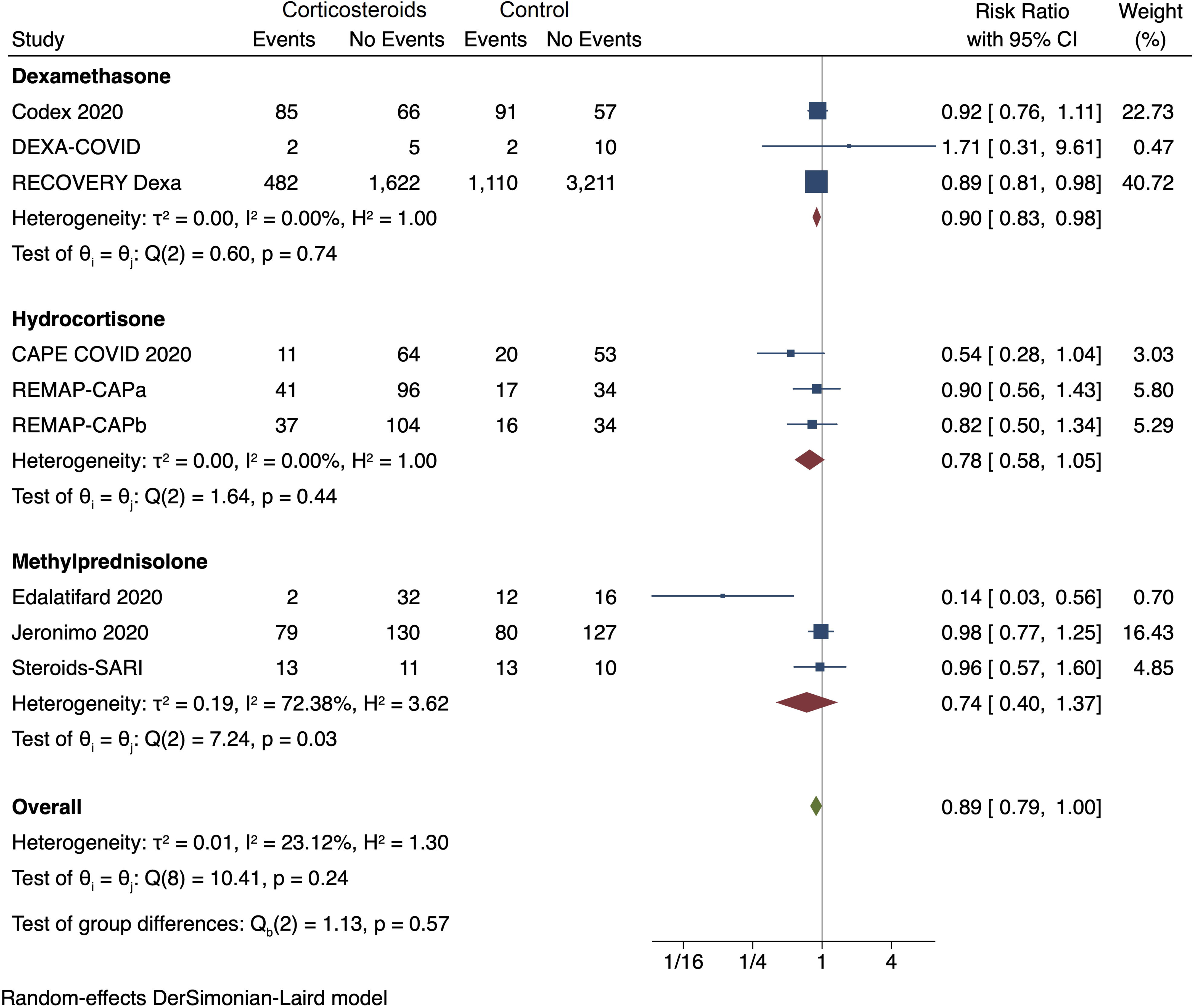
Random-effects meta-analysis for corticosteroids versus control (standard care or placebo) on all-cause mortality

**Fig 3.**
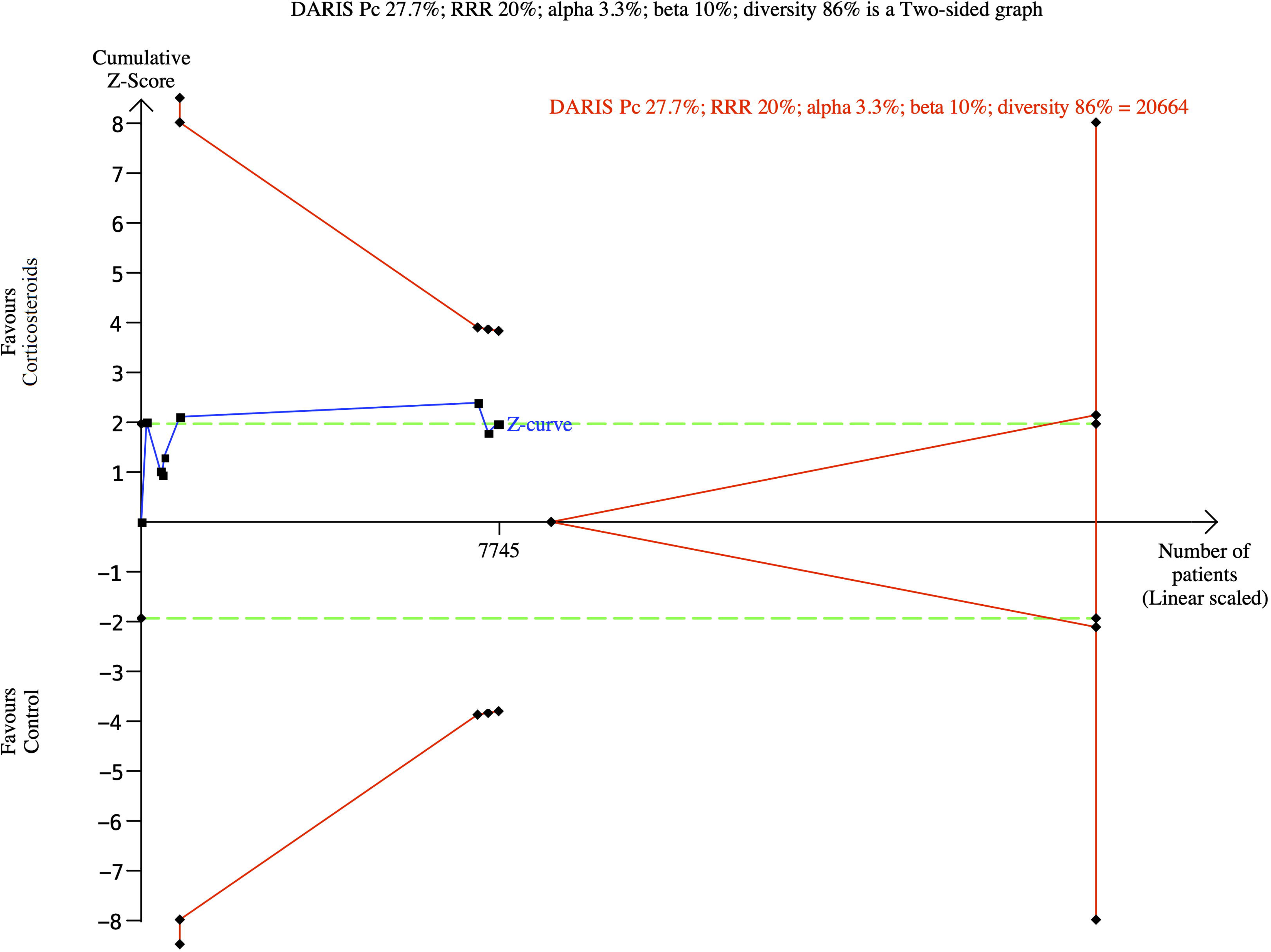
Trial Sequential Analysis for corticosteroids versus control (standard care or placebo) on all-cause mortality

#### Meta-analysis of serious adverse events

Random-effects meta-analysis showed no evidence of a difference between corticosteroids and control on serious adverse events (RR 0.89; 95% CI 0.80 to 0.99; *p* = 0.04; I^2^ = 39.1%; eight trials; very low certainty) (**S3 Fig, S4 Table**). Fixed-effect meta-analysis showed evidence of a beneficial effect of corticosteroids versus control on serious adverse events (RR 0.88; 95% CI 0.82 to 0.95; *p* = 0.00; I^2^ = 43.1%; eight trials) (**S4 Fig**). Visual inspection of the forest plot and measures to quantify heterogeneity (I^2^ = 39.1%) indicated no substantial heterogeneity. The time-points of assessment varied from 21 [88] to 30 days after randomization [97, 118]. The trial sequential analysis showed that we did not have enough information to confirm or reject that corticosteroids versus control reduce the risk of serious adverse events with a relative risk reduction of 20% (**S5 Fig**). The subgroup analysis assessing the effects of the different corticosteroids versus control showed no significant subgroup differences (*p* = 0.71) (**S3 Fig**). The serious adverse event data is predominately based on mortality data, and assessed according to the ICH-GCP definition of a serious adverse event [15].

#### Meta-analysis of mechanical ventilation

Random-effects meta-analysis showed no evidence of a difference between corticosteroids versus control on mechanical ventilation (RR 0.86; 95% CI 0.55 to 1.33; *p* = 0.49; I^2^ = 55.3%; two trials; very low certainty) (**S6 Fig, S4 Table**). Fixed-effect meta-analysis showed evidence of a beneficial effect of corticosteroids versus control on mechanical ventilation (RR 0.77; 95% CI 0.63 to 0.94; *p* = 0.01; I^2^ = 55.4%, two trials) (**S7 Fig**). Visual inspection of the forest plot and measures to quantify heterogeneity (I^2^ = 55.3%) indicated moderate heterogeneity. The time-points of assessment varied from 7 days [68] to 28 days after randomization [47]. The trial sequential analysis showed that we did not have enough information to confirm or reject that corticosteroids versus control reduce the risk of receiving mechanical ventilation with a relative risk reduction of 20% (**S8 Fig**). The subgroup analysis assessing the effects of the different corticosteroids versus control showed no significant subgroup differences (*p* = 0.13) (**S6 Fig**). One of the two trials [68] had substantial missing data for this outcome, but it was a small trial that did not contribute with much data compared to the second trial.

### Remdesivir versus control

We identified four trials randomizing 7,370 participants to remdesivir versus standard care [86, 110] or placebo [43, 65]. All trials were assessed at high risk of bias (**S3 Table**). One trial assessed two different dosages of remdesivir versus standard care [86], and the two comparisons were both included in the meta-analysis. We halved the control group to avoid double counting [11].

#### Meta-analysis of all-cause mortality

Random-effects meta-analysis showed no evidence of a difference between remdesivir versus control on all-cause mortality (RR 0.93; 95% CI 0.82 to 1.07; *p* = 0.31; I^2^ = 0%; four trials; moderate certainty) (**Fig 4**, **S5 Table**). Visual inspection of the forest plot and measures to quantify heterogeneity (I^2^ = 0%) indicated no heterogeneity. The assessment time points were 28 [43,86,110] and 29 days after randomization [65]. The trial sequential analysis showed that we had enough information to reject that remdesivir versus control reduces the risk of all-cause mortality with a relative risk reduction of 20% (**Fig 5**). The subgroup analysis assessing the effects of the different control interventions showed no significant subgroup differences (*p* = 0.21) (**Fig 4**). The subgroup analysis assessing the effects of early versus late intervention (defined as no oxygen versus oxygen or respiratory support at baseline) showed no significant subgroup differences (*p* = 0.85) (**S9 Fig**).

**Fig 4.**
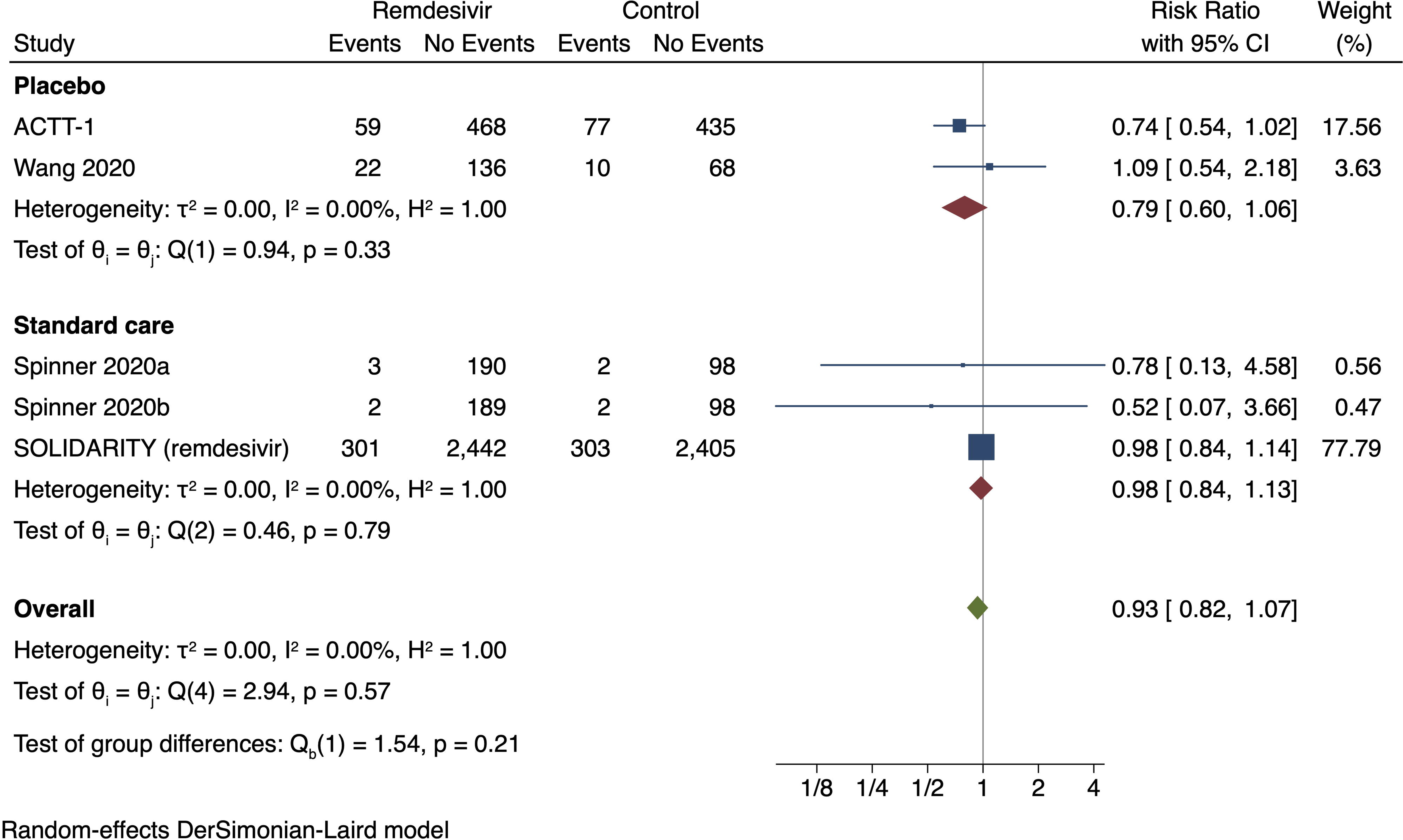
Random-effects meta-analysis for remdesivir versus control (standard care or placebo) on all-cause mortality

**Fig 5.**
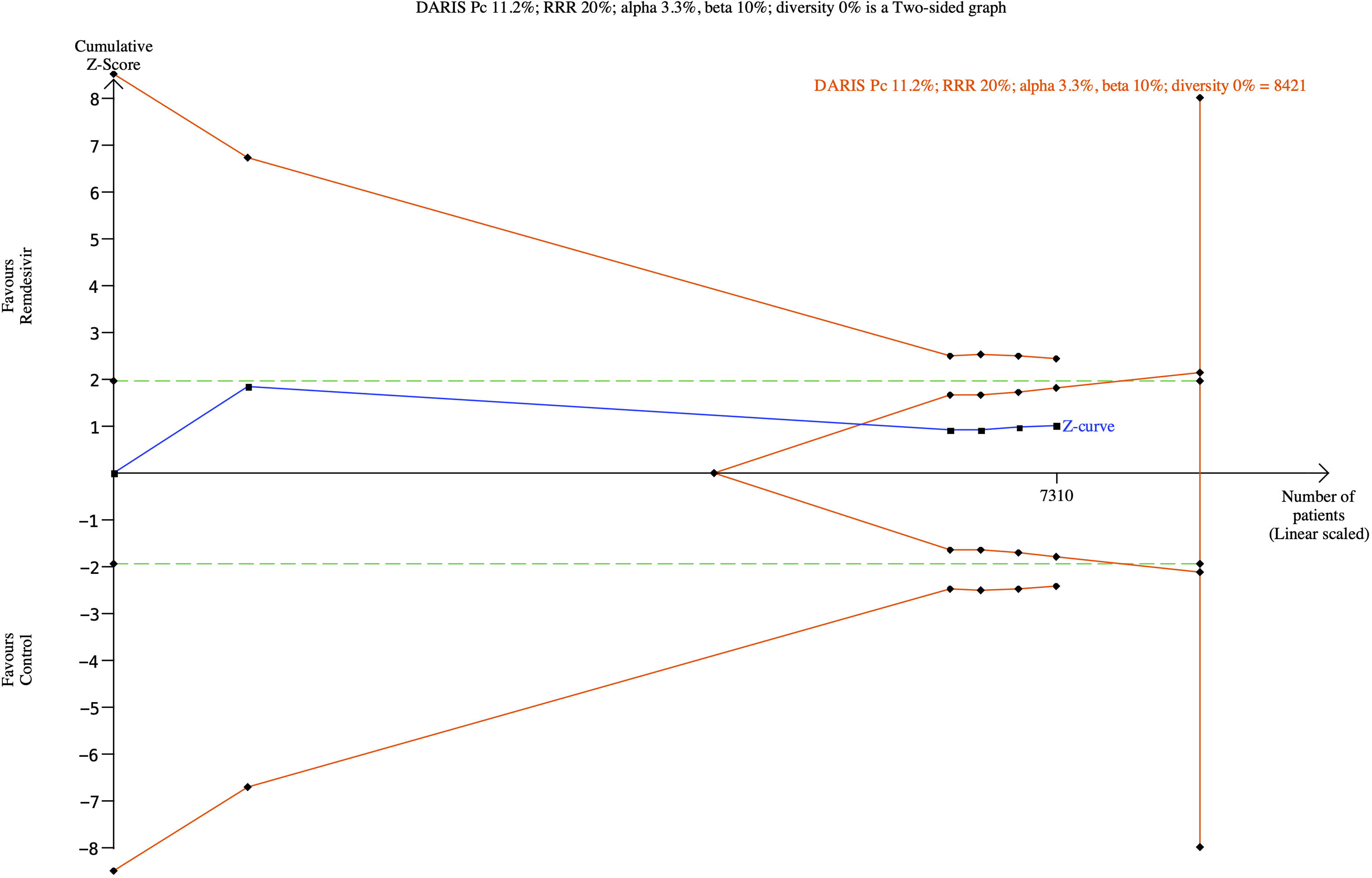
Trial Sequential Analysis for remdesivir versus control (standard care or placebo) on all-cause mortality

#### Meta-analysis of serious-adverse events

Random-effects meta-analysis showed no evidence of difference between remdesivir versus control on serious adverse events (RR 0.82; 95% CI 0.68 to 1.00; *p* = 0.05; I^2^ = 38.9%; four trials; very low certainty) (**S10 Fig, S5 Table**). Fixed-effect meta-analysis showed evidence of a beneficial effect of remdesivir versus control on serious adverse events (RR 0.88; 95% CI 0.79 to 0.99; *p* = 0.03; I^2^ = 39.2%; four trials) (**S11 Fig**). Visual inspection of the forest plot and measures to quantify heterogeneity (I^2^ = 38.9%) indicated some heterogeneity. The assessment time points were 28 [43,86,110] and 29 days after randomization [65]. The trial sequential analysis showed that we did not have enough information to confirm or reject that remdesivir versus control reduces the risk of serious adverse events with a relative risk reduction of 20% (**S12 Fig**). The subgroup analysis assessing the effects of the different control interventions showed no significant subgroup differences (*p* = 0.83) (**S10 Fig**).

#### Meta-analysis of mechanical ventilation

Random-effects meta-analysis showed no evidence of a difference between remdesivir versus control on mechanical ventilation (RR 0.73; 95% CI 0.42 to 1.27; *p* = 0.27; I^2^ = 83.1%; three trials; very low certainty) (**S13 Fig, S5 Table**). Visual inspection of the forest plot and measures to quantify heterogeneity (I^2^ = 83.1%) indicated substantial heterogeneity. The assessment time points were 28 [43, 110] and 29 days after randomization [65]. The trial sequential analysis showed that we did not have enough information to confirm or reject that remdesivir versus control reduces the risk of receiving mechanical ventilation with a relative risk reduction of 20% (**S14 Fig**). The subgroup analysis assessing the effects of the different control interventions showed evidence of a significant subgroup difference between placebo and standard care (*p* = 0.00) (**S13 Fig**).

#### Meta-analysis of non-serious adverse events

Fixed-effect meta-analysis showed no evidence of a difference between remdesivir versus control on non-serious adverse events (RR 0.99; 95% CI 0.91 to 1.08; *p* = 0.86; I^2^ = 56.4%; three trials; very low certainty) (**S15 Fig, S5 Table**). Visual inspection of the forest plot and measures to quantify heterogeneity (I^2^ = 56.4%) indicated moderate heterogeneity. The assessment time point was 28 days after randomization [43,65,86]. The Trial Sequential Analysis showed that we had enough information to reject that remdesivir versus control reduces the risk of non-serious adverse events with a relative risk reduction of 20% (**S16 Fig**). The subgroup analysis assessing the effects of the different control interventions showed evidence of subgroup difference between placebo and standard care (*p* = 0.02) (**S15 Fig**).

### Hydroxychloroquine versus control interventions

We identified 13 trials randomizing 10,276 participants to hydroxychloroquine versus standard care [34,35,42,48,54,55,58,59,105,110], or placebo [53, 108]. All trials were assessed at high risk of bias (**S3 Table**). One trial was not eligible for meta-analysis, as the results were not reported in a usable way; i.e., the results were reported as per-protocol and several participants crossed over [42].

#### Meta-analysis of all-cause mortality

Fixed-effect meta-analysis showed no evidence of a difference between hydroxychloroquine versus control on all-cause mortality (RR 1.09; 95% CI 0.99 to 1.20; *p* = 0.08; I^2^ = 0%; 10 trials; moderate certainty) (**S17 Fig, S6 Table**). Visual inspection of the forest plot and measures to quantify heterogeneity (I^2^ = 0%) indicated no heterogeneity. The assessment time points varied from five days after randomization [34, 35] to 30 days after randomization [108]. The trial sequential analysis showed that we had enough information to reject that hydroxychloroquine versus control reduces the risk of all-cause mortality with a relative risk reduction of 20% (**S18 Fig**). The subgroup analysis assessing the effects of hydroxychloroquine versus different control interventions showed no significant subgroup differences (*p* = 0.92) (**S17 Fig**).

#### Meta-analysis of serious adverse events

Fixed-effect meta-analysis showed no evidence of a difference between hydroxychloroquine versus control on serious adverse events (RR 1.08; 95% CI 0.98 to 1.19; p = 0.11; I^2^ = 0%; 11 trials; moderate certainty) (**S19 Fig, S6 Table**). Visual inspection of the forest plot and measures to quantify heterogeneity (I^2^ = 0%) indicated no heterogeneity. The assessment time points varied from five days after randomization [34, 35] to 30 days after randomization [108]. The trial sequential analysis showed that we had enough information to reject that hydroxychloroquine versus control reduces the risk of serious adverse events with a relative risk reduction of 20% (**S20 Fig).** The subgroup analysis assessing the effects of hydroxychloroquine versus different control interventions showed no significant subgroup differences (*p* = 0.90) (**S19 Fig**).

#### Meta-analysis of admission to intensive care

Fixed-effect meta-analysis showed no evidence of a difference between hydroxychloroquine versus control on admission to intensive care (RR 0.74; 95% CI 0.44 to 1.25; p = 0.26; I^2^ = 0%; two trials; very low certainty) (**S21 Fig, S6 Table**). Visual inspection of the forest plot and measures to quantify heterogeneity (I^2^ = 0%) indicated no substantial heterogeneity. The assessment time points were 28 days after randomization [77] and 30 days after randomization [108]. The trial sequential analysis showed that we did not have enough information to confirm or reject that hydroxychloroquine versus control reduces the risk of admission to intensive care with a relative risk reduction of 20% (**S22 Fig**). The subgroup analysis assessing the effects of hydroxychloroquine versus different control interventions showed no significant subgroup differences (*p* = 0.61) (**S21 Fig**)

#### Meta-analysis of mechanical ventilation

Fixed-effect meta-analysis showed no evidence of a difference between hydroxychloroquine versus control on mechanical ventilation (RR 1.10; 95% CI 0.84 to 1.45; p = 0.48; I^2^ = 0.00%; four trials; very low certainty) (**S23 Fig, S6 Table**). Visual inspection of the forest plot and measures to quantify heterogeneity (I^2^ = 0%) indicated no heterogeneity. The assessment time points were 15 days after randomization [54] and 30 days after randomization [108]. The trial sequential analysis showed that we did not have enough information to confirm or reject that hydroxychloroquine versus control reduces the risk of receiving mechanical ventilation with a relative risk reduction of 20% (**S24 Fig**). The subgroup analysis assessing the effects of hydroxychloroquine versus different control interventions showed no significant subgroup differences (*p* = 0.84) (**S23 Fig**).

#### Meta-analysis of non-serious adverse events

Random-effects meta-analysis showed evidence of a harmful effect of hydroxychloroquine versus control on non-serious adverse events (RR 2.09; 95% CI 1.14 to 3.80; p = 0.02; I^2^ = 92.1%; six trials; very low certainty) (**S25 Fig, S6 Table**). Visual inspection of the forest plot and measures to quantify heterogeneity (I^2^ = 92.1%) indicated substantial heterogeneity. The assessment time points were five days after randomization [34] and 30 days after randomization [108]. The trial sequential analysis showed that we did not have enough information to confirm or reject that hydroxychloroquine versus control reduces the risk of non-serious adverse events with a relative risk reduction of 20%. The subgroup analysis assessing the effects of hydroxychloroquine versus different control interventions showed a significant subgroup difference between standard care and placebo (*p* = 0.39) (**S25 Fig**).

### Lopinavir-ritonavir versus control interventions

We identified four trials randomizing 8,081 participants to lopinavir-ritonavir versus standard care [32,40,106,110]. We also identified one trial randomising 60 participants to lopinavir-ritonavir and novaferon versus novaferon alone [45]. All trials were assessed at high risk of bias (**S3 Table**).

#### Meta-analysis of all-cause mortality

Fixed-effect meta-analysis showed no evidence of a difference between lopinavir-ritonavir versus control on all-cause mortality (RR 1.01; 95% CI 0.92 to 1.12; *p* = 0.77; I^2^ = 0.0%; five trials; moderate certainty) (**S26 Fig, S7 Table)**. Visual inspection of the forest plot and measures to quantify heterogeneity (I^2^ = 0.0%) indicated no heterogeneity. The time-points of assessment were nine days after randomization in one trial [45], 21 days after randomization in one trial [40], 28 days after randomization in two trials [32, 110], and 28 days or until discharge or death in one trial [106]. The trial sequential analysis showed that we had enough information to reject that lopinavir-ritonavir versus control reduces the risk of all-cause mortality with a relative risk reduction of 20% (**S27 Fig**). The subgroup analysis assessing the effects of lopinavir-ritonavir in combination with novaferon versus lopinavir-ritonavir alone showed no significant subgroup differences (p = 0.99) (**S26 Fig**).

#### Meta-analysis of serious adverse events

Random-effects meta-analysis showed no evidence of a difference between lopinavir-ritonavir versus control on serious adverse events (RR 1.00; 95% CI 0.91 to 1.11; *p* = 0.93; I^2^ = 1.2%; five trials; moderate certainty) (**S28 Fig, S7 Table**). Visual inspection of the forest plot and measures to quantify heterogeneity (I^2^ = 1.2%) indicated no substantial heterogeneity. The time-points of assessment were nine days after randomization in one trial [45], 21 days after randomization in one trial [40], 28 days after randomization in two trials [32, 110], and 28 days or until discharge or death in one trial [106]. The trial sequential analysis showed that we had enough information to reject that lopinavir-ritonavir versus control reduces the risk of serious adverse events with a relative risk reduction of 20% (**S29 Fig**). The subgroup analysis assessing the effects of lopinavir-ritonavir in combination with novaferon versus lopinavir-ritonavir alone showed no significant subgroup differences (p = 0.99) (**S28 Fig**).

#### Meta-analysis of mechanical ventilation

Random-effects meta-analysis showed no evidence of a difference between lopinavir-ritonavir versus control on mechanical ventilation (RR 1.08; 95% CI 0.94 to 1.25; *p* = 0.29; I^2^ = 0.0%; three trials; moderate certainty) (**S30** **Fig 2, S7 Table**). Visual inspection of the forest plot and measures to quantify heterogeneity (I^2^ = 0.0%) indicated no heterogeneity. The time-points of assessment were 28 days after randomization in two trials [32, 79] and 28 days or until discharge or death for one trial [106]. The trial sequential analysis showed that we had enough information to reject that lopinavir-ritonavir versus control reduces the risk of receiving mechanical ventilation with a relative risk reduction of 20% (**S31 Fig**).

#### Meta-analysis of renal replacement therapy

Random-effects meta-analysis showed no evidence of a difference between lopinavir-ritonavir versus control on renal replacement therapy (RR 0.97; 95% CI 0.73 to 1.28; *p* = 0.81; I^2^ = 0.0%; two trials; very low certainty) (**S32 Fig, S7 Table**). Visual inspection of the forest plot and measures to quantify heterogeneity (I^2^ = 0.0%) indicated no heterogeneity. The time-points of assessment was 28 days for the first trial [32] and 28 days or until discharge or death for the second trial [106]. The trial sequential analysis showed that we did not have enough information to confirm or reject that lopinavir-ritonavir versus control reduces the risk of renal replacement therapy with a relative risk reduction of 20% (**S33 Fig**).

#### Meta-analysis of non-serious adverse events

Random-effects meta-analysis showed no evidence of a difference between lopinavir–ritonavir versus standard care on non-serious adverse events (RR 1.14; 95% CI 0.85–1.53; *p* = 0.38, I^2^ = 75%; two trials; very low certainty) (**S34 Fig**; **S7 Table**). Visual inspection of the forest plot and measures to quantify heterogeneity (I^2^ = 75%) indicated substantial heterogeneity. The assessment time point was 21 days after randomization in the first trial [40] and 28 days after randomization in the second trial [3]. The trial sequential analysis showed that we did not have enough information to confirm or reject that lopinavir–ritonavir compared with standard care reduces nonserious adverse events with a relative risk reduction of 20%.

### Interferon β-1a versus control

We identified two trials randomizing 4,219 participants to interferon β-1a versus standard care [36, 110]. In one of the trials, the first 1,200 participants received interferon β-1a and lopinavir-ritonavir or lopinavir-ritonavir alone, while the remaining 2,927 participants received interferon β-1a or standard care [110]. All trials were assessed at high risk of bias (**S3 Table**).

#### Meta-analysis of all-cause mortality

Random-effects meta-analysis showed no evidence of a difference between interferon β-1a versus standard care on all-cause mortality (RR 0.75; 95% CI 0.30 to 1.88; *p* = 0.54; I^2^ = 84.1%; two trials; very low certainty) (**S35 Fig, S8 Table)**. Visual inspection of the forest plot and measures to quantify heterogeneity (I^2^ = 84.1%) indicated substantial heterogeneity. The time-point of assessment was 28 days after randomization in both trials [36, 110]. The trial sequential analysis showed that we did not have enough information to confirm or reject that interferon β-1a versus standard care reduces the risk of all-cause mortality with a relative risk reduction of 20%.

#### Meta-analysis of serious adverse events

Random-effects meta-analysis showed no evidence of a difference between interferon β-1a versus standard care on serious adverse events (RR 0.75; 95% CI 0.30 to 1.88; *p* = 0.54; I^2^ = 84.1%; two trials; very low certainty) (**S36 Fig, S8 Table)**. However, the data was solely based on all-cause mortality data, since no other serious adverse events were reported [15]. Visual inspection of the forest plot and measures to quantify heterogeneity (I^2^ = 84.1%) indicated substantial heterogeneity. The time-point of assessment was 28 days after randomization in both trials [36, 110]. The trial sequential analysis showed that we did not have enough information to confirm or reject that interferon β-1a versus standard care reduces the risk of serious adverse events with a relative risk reduction of 20%.

### Convalescent plasma versus control

We identified four trials randomizing 734 participants to convalescent plasma versus standard care [39,51,78,91]. All trials were assessed as at high risk of bias (**S3 Table**).

#### Meta-analysis of all-cause mortality

Random-effects meta-analysis showed no evidence of difference between convalescent plasma versus standard care on all-cause mortality (RR 0.77; 95% CI 0.47 to 1.24; *p* = 0.28; I^2^ = 27.5%; four trials; very low certainty) (**S37 Fig, S9 Table**). Visual inspection of the forest plot and measures to quantify heterogeneity (I^2^ = 27.5%) indicated some heterogeneity. The outcome was assessed 28 days after randomization in two trials [39, 91], at 29 days after randomization in one trial [78] and up to hospital discharge or 60 days in one trial [51]. The trial sequential analysis showed that we did not have enough information to confirm or reject that convalescent plasma versus standard care reduces the risk of all-cause mortality with a relative risk reduction of 20% (**S38 Fig**).

#### Meta-analysis of serious adverse events

Fixed-effect meta-analysis showed no evidence of a difference between convalescent plasma versus standard care on serious adverse events (RR 0.93; 95% CI 0.64 to 1.35; *p* = 0.70; I^2^ = 0.0%; three trials; very low certainty) (**S39 Fig, S9 Table**). Visual inspection of the forest plot and measures to quantify heterogeneity (I^2^ = 0.0%) indicated no substantial heterogeneity. The time-point of assessment was 28 days after randomization in two trials [39, 91], 29 days after randomization in one trial [78], and up to hospital discharge or 60 days in the last trial [51]. The trial sequential analysis showed that we did not have enough information to confirm or reject that convalescent plasma versus standard care reduces the risk of serious adverse events with a relative risk reduction of 20% (**S40 Fig**)

### Azithromycin versus control

We identified three trials randomizing 996 participants to azithromycin versus standard care [83] or versus co-interventions with standard care [54], or without standard care [82]. All trials were assessed at high risk of bias (**S3 Table**). One trial assessed the effects of azithromycin versus standard care [54], one trial assessed the effects of azithromycin plus lopinavir-ritonavir and hydroxychloroquine versus lopinavir-ritonavir and hydroxychloroquine alone [82], and one trial assessed the effects of azithromycin plus hydroxychloroquine and standard care versus hydroxychloroquine and standard care alone [54].

#### Meta-analysis of all-cause mortality

Fixed-effect meta-analysis showed no evidence of a difference between azithromycin versus control on all-cause mortality (RR 0.99; 95% CI 0.79 to 1.25; *p* = 0.95; I^2^ = 7.4%; three trials; very low certainty) (**S41 Fig, S10 Table**).Visual inspection of the forest plot and measures to quantify heterogeneity (I^2^ = 7.4%) indicated no substantial heterogeneity. The time-point of assessment was 15 days after randomization in the first trial [54], 29 days after randomization in the second trial [83], and unclear in the third trial [82]. We have contacted the trial authors and requested information on the assessment time-point, but we have not received a response yet. The trial sequential analysis showed that we did not have enough information to confirm or reject that azithromycin versus control reduces the risk of all-cause mortality with a relative risk reduction of 20% (**S42 Fig**). The subgroup analysis assessing the effects of azithromycin versus different control interventions showed no significant subgroup differences (*p* = 0.35) (**S41 Fig**).

#### Meta-analysis of serious adverse events

Random-effects meta-analysis showed no evidence of a difference between azithromycin versus control on serious adverse events (RR 0.95; 95% CI 0.55 to 1.63; *p* = 0.84; I^2^ = 16%; three trials; very low certainty) (**S43 Fig, S10 Table**). Visual inspection of the forest plot and measures to quantify heterogeneity (I^2^ = 16%) indicated no substantial heterogeneity. The time-point of assessment was 15 days after randomization in the first trial [54], and 29 days after randomization in the second trial [83], and unclear in the third trial [82]. We have contacted the trial authors and requested information on the assessment time-point, but we have not received a response yet. The trial sequential analysis showed that we did not have enough information to confirm or reject that azithromycin versus control reduces the risk of serious adverse events with a relative risk reduction of 20%. The subgroup analysis assessing the effects of azithromycin versus different control interventions showed no significant subgroup differences (*p* = 0.30) (**S43 Fig**).

#### Meta-analysis of mechanical ventilation

Fixed-effect meta-analysis showed no evidence of a difference between azithromycin versus control on mechanical ventilation (RR 1.07; 95% CI 0.59 to 1.94; p = 0.83; I^2^ = 53%; two trials; very low certainty) (**S44 Fig, S10 Table**). Visual inspection of the forest plot and measures to quantify heterogeneity (I^2^ = 53%) indicated moderate heterogeneity. The time-point of assessment was 15 days after randomization in the first trial [54], and unclear in the second trial [82]. We have contacted the trial authors and requested information on the assessment time-point, but we have not received a response yet. The trial sequential analysis showed that we did not have enough information to confirm or reject that azithromycin versus control reduces the risk of receiving mechanical ventilation with a relative risk reduction of 20%. The subgroup analysis assessing the effects of azithromycin versus different control interventions showed no significant subgroup differences (*p* = 0.15) (**S44 Fig**).

#### Meta-analysis of non-serious adverse events

Fixed-effect meta-analysis showed no evidence of a difference between azithromycin versus control on non-serious adverse events (RR 1.09; 95% CI 0.89 to 1.34; p = 0.38; I^2^ = 0%; two trials; very low certainty) (**S45 Fig, S10 Table**). Visual inspection of the forest plot and measures to quantify heterogeneity (I^2^ = 0%) indicated no heterogeneity. The time-point of assessment was 15 days after randomization in the first trial [54], and 29 days after randomization in the second trial [83]. The trial sequential analysis showed that we did not have enough information to confirm or reject that azithromycin versus control reduces the risk of non-serious adverse events with a relative risk reduction of 20% (**S46 Fig**). The subgroup analysis assessing the effects of azithromycin versus different control interventions showed no significant subgroup differences (*p* = 0.71) (**S45 Fig**)

### Colchicine versus control

We identified three trials randomizing 248 participants to colchicine versus standard care [49], placebo plus standard care [92], or placebo plus hydroxychloroquine [107]. In the latter trial, the colchicine group also received hydroxychloroquine as a co-intervention [107]. All trials were assessed as at high risk of bias (**S3 Table**).

#### Meta-analysis of all-cause mortality

Fixed-effect meta-analysis showed no evidence of a difference between colchicine versus control on all-cause mortality (RR 1.03; 95% CI 0.07 to 16.01; p = 0.98; I^2^ = 0%; two trials; very low certainty) (**S47 Fig, S11 Table**). Visual inspection of the forest plot and measures to quantify heterogeneity (I^2^ = 0%) indicated no heterogeneity. The time-point of assessment was unclear in both trials [92, 107]. We have contacted the trial authors and requested information on the assessment time-points, but we have not received a response yet. The trial sequential analysis showed that we did not have enough information to confirm or reject that colchicine versus control reduces the risk of all-cause mortality with a relative risk reduction of 20%. The subgroup analysis assessing the effects of colchicine versus different control interventions showed no evidence of a significant subgroup difference (*p* = 0.98) (**S47 Fig**).

#### Meta-analysis of non-serious adverse events

Random-effects meta-analysis showed no evidence of a difference between colchicine versus control on non-serious adverse events (RR 0.88; 95% CI 0.18 to 4.39; *p* = 0.87; I^2^ = 79.1%; three trials; very low certainty) (**S48 Fig, S11 Table**). Visual inspection of the forest plot and measures to quantify heterogeneity (I^2^ = 79.1%) indicated substantial heterogeneity. The time-point of assessment was 21 days after randomization in one trial [49], but unclear in the other two trials [92, 107]. We have contacted the trial authors and requested information on the assessment time-points, but we have not received a response yet. The trial sequential analysis showed that we did not have enough information to confirm or reject that colchicine versus control reduces the risk of non-serious adverse events with a relative risk reduction of 20%. The subgroup analysis assessing the effects of colchicine versus different control interventions showed evidence of significant subgroup differences (*p* = 0.01) (**S48 Fig**)

### Intravenous immunoglobin versus control

We identified two trials randomizing 93 participants to intravenous immunoglobulin versus standard care [57] or placebo [95]. Both trials included immunoglobulin from healthy donors [57, 95]. Both trials were assessed at high risk of bias (**S3 Table**).

#### Meta-analysis of all-cause mortality

Fixed-effect meta-analysis showed evidence of a beneficial effect of intravenous immunoglobulin versus control on all-cause mortality (RR 0.40; 95% CI 0.19 to 0.87; *p* = 0.02; I^2^ = 0%; two trials; very low certainty) (**S49 Fig, S12 Table)**. Visual inspection of the forest plot and measures to quantify heterogeneity (I^2^ = 0%) indicated no heterogeneity. The outcome was assessed only in hospital in the first trial [95] and up to 30 days in the second trial [57]. The trial sequential analysis showed that we did not have enough information to confirm or reject that intravenous immunoglobulin versus control reduces the risk of all-cause mortality with a relative risk reduction of 20% (**S50 Fig**). The subgroup analysis assessing the effects of different control interventions showed no evidence of a significant subgroup difference between placebo and standard care (*p* = 0.89) (**S49 Fig**)

#### Meta-analysis of serious adverse events

Fixed-effect meta-analysis showed evidence of a beneficial effect of intravenous immunoglobulin versus control on serious adverse events (RR 0.40; 95% CI 0.19 to 0.87; *p* = 0.02; I^2^ = 0%; two trials; very low certainty) (**S51 Fig, S12 Table**). This data is solely based on all-cause mortality data according to the ICH-GCP guidelines [15], since no other serious adverse events were reported. Visual inspection of the forest plot and measures to quantify heterogeneity (I^2^ = 0%) indicated no heterogeneity. The outcome was assessed only in hospital in the first trial [95] and up to 30 days in the second trial [57]. The trial sequential analysis showed that we did not have enough information to confirm or reject that intravenous immunoglobulin versus control reduces the risk of all-cause mortality with a relative risk reduction of 20% (**S52 Fig**). The subgroup analysis assessing the effects of different control interventions showed no evidence of a significant subgroup difference between placebo and standard care (*p* = 0.89) (**S51 Fig**)

### Tocilizumab versus control

We identified six trials randomizing 1038 patients to tocilizumab versus standard care [93,111–113], placebo with standard care [90] or favipiravir alone as co-intervention [114]. All trials were assessed as at high risk of bias (**S3 Table**).

#### Meta-analysis of all-cause mortality

Random-effects meta-analysis showed no evidence of a difference between tocilizumab and control interventions on all-cause mortality (RR 1.03; 95% CI 0.72 to 1.46; *p* = 0.89; I^2^ = 0.0%; four trials; very low certainty) (**S53 Fig, S13 Table)**. Visual inspection of the forest plot and measures to quantify heterogeneity (I^2^ = 0.0%) indicated no heterogeneity. The time-points of assessment were 28 days [90,111,113] and 30 days [112] after randomization. The trial sequential analysis showed that we did not have enough information to confirm or reject that tocilizumab versus control reduces the risk of all-cause mortality with a relative risk reduction of 20% (**S54 Fig)**. The subgroup analysis assessing the effects of different control interventions showed no significant subgroup differences (p = 0.87) (**S53 Fig**)

#### Meta-analysis of serious adverse events

Random-effects meta-analysis showed no evidence of a difference between tocilizumab and control interventions on serious adverse events (RR 0.63; 95% CI 0.35 to 1.14; *p* = 0.12; I^2^ = 77.4%; five trials; very low certainty) (**S55 Fig, S13 Table**). Fixed-effect meta-analysis showed evidence of a beneficial effect of tocilizumab versus control on serious adverse events (RR 0.68; 95% CI 0.57 to 0.81; *p* = 0.00; I^2^ = 77.5%, five trials) (**S56 Fig**). Visual inspection of the forest plot and measures to quantify heterogeneity (I^2^ = 77.4%) indicated heterogeneity. The time-point of assessment was either unclear [90, 114], 28 days [111, 113], or 30 days [112] after randomization. The trial sequential analysis showed that we did not have enough information to confirm or reject that tocilizumab versus control reduces the risk of serious adverse events with a relative risk reduction of 20%. The subgroup analysis assessing the effects of different control interventions showed no significant subgroup differences (p = 0.13) (**S55 Fig**).

#### Meta-analysis of admission to intensive care

Random-effects meta-analysis showed no evidence of a difference between tocilizumab and control interventions on admission to intensive care (RR 0.71; 95% CI 0.37 to 1.38; *p* = 0.32; I^2^ = 36%; two trials; very low certainty) (**S57 Fig, S13 Table**). Visual inspection of the forest plot and measures to quantify heterogeneity (I^2^ = 36%) indicated no substantial heterogeneity. The time-point of assessment was either unclear [90] or 30 days [112] after randomization. The trial sequential analysis showed that we did not have enough information to confirm or reject that tocilizumab versus control reduces the risk of admission to intensive care with a relative risk reduction of 20%. The subgroup analysis assessing the effects of control interventions showed no significant subgroup differences (p = 0.21) (**S57 Fig**).

#### Meta-analysis of mechanical ventilation

Random-effects meta-analysis showed evidence of a beneficial effect of tocilizumab versus control on mechanical ventilation (RR 0.70; 95% CI 0.51 to 0.96; *p* = 0.02; I^2^ = 0%; three trials; very low certainty) (**S58 Fig, S13 Table**). Visual inspection of the forest plot and measures to quantify heterogeneity (I^2^ = 0%) indicated no heterogeneity. The time-point of assessment was either unclear [90] or 28 days [111, 113] after randomization. The trial sequential analysis showed that we did not have enough information to confirm or reject that tocilizumab reduce the risk of mechanical ventilation with a relative risk reduction of 20% (**S59 Fig**). The subgroup analysis assessing the effects of control interventions showed no significant subgroup differences (p = 0.34) (**S58 Fig**).

#### Meta-analysis of non-serious adverse events

Fixed-effect meta-analysis showed no evidence of a difference between tocilizumab versus control on non-serious adverse events (RR 1.03; 95% CI 0.92 to 1.14; *p* = 0.63; I^2^ = 57.9%; five trials; very low certainty) (**S60 Fig, S13 Table**). Visual inspection of the forest plot and measures to quantify heterogeneity (I^2^ = 57.9%) indicated moderate heterogeneity. The time-point of assessment was either unclear [90,93,114], 28 days [111], or 30 days [112] after randomization. The trial sequential analysis showed that we did not have enough information to confirm or reject that tocilizumab versus control reduces the risk of non-serious adverse events with a relative risk reduction of 20%. The subgroup analysis assessing the effects of different control interventions showed no significant subgroup differences (p = 0.27) (**S60 Fig**).

### Bromhexidine versus control

We identified two trials randomizing 96 participants to bromhexidine versus standard care [94, 104]. Both trials were assessed at high risk of bias (**S3 Table**).

#### Meta-analysis of all-cause mortality

Random-effects meta-analysis showed no evidence of a difference between bromhexidine versus standard care on all-cause mortality (RR 0.17; 95% CI 0.02 to 1.70; *p* = 0.13; I^2^ = 0%; two trials; very low certainty certainty) (**S61 Fig, S14 Table)**. Visual inspection of the forest plot and measures to quantify heterogeneity (I^2^ = 0%) indicated no heterogeneity. The time-point of assessment was 28 days for the first trial [104] and unclear for the second trial [94]. The trial sequential analysis showed that we did not have enough information to confirm or reject that bromhexidine versus standard care reduces the risk of all-cause mortality with a relative risk reduction of 20%.

#### Meta-analysis of non-serious adverse events

Random-effects meta-analysis showed evidence of a beneficial effect of bromhexidine versus standard care on non-serious adverse events (RR 0.32; 95% CI 0.15 to 0.69; *p* < 0.00; I^2^ = 0%; two trials; very low certainty) (**S62 Fig, S14 Table)**. Visual inspection of the forest plot and measures to quantify heterogeneity (I^2^ = 0%) indicated no heterogeneity. The time-point of assessment was 28 days for the first trial [104] and unclear for the second trial [94]. The trial sequential analysis showed that we did not have enough information to confirm or reject that bromhexidine versus standard care reduces the risk of non-serious adverse events with a relative risk reduction of 20%.

### Remaining trial data

Because of lack of relevant data, it was not possible to conduct other meta-analyses, individual patient data meta-analyses, or network meta-analysis. Nine single trials showed statistically significant results but were all underpowered to confirm or reject realistic intervention effects.

One trial randomizing 402 participants compared five versus ten days of remdesivir and showed evidence of a beneficial effect of five days of remdesivir on serious adverse events (*p* = 0.003 (Fisher’s exact test)) [37]. One trial randomizing 92 participants compared the immunomodulator interferon β-1a added to standard care versus standard care alone and showed evidence of a beneficial effect of interferon β-1a on all-cause mortality (*p* = 0.029) [36]. This trial also showed evidence of a harmful effect of interferon β-1a on non-serious adverse events (*p* = 0.006) [36]. One single trial randomizing 81 participants compared high-dosage versus low-dosage chloroquine diphosphate and showed evidence of a beneficial effect of low-dosage chloroquine on all-cause mortality (*p* = 0.024) [50]. One single three group trial randomizing 667 participants to hydroxychloroquine with or without azithromycin versus standard care and showed evidence of a harmful effect of hydroxychloroquine with azithromycin on adverse events not considered serious (*p* = 0.015) [54]. One single trial randomizing 76 participants compared calcifediol versus standard care and showed evidence of a beneficial effect of calcifediol on admittance to intensive care (*p* = 0.0001) [84]. One single trial randomizing 200 participants compared recombinant human granulocyte colony–stimulating factor (rhG-CSF) versus standard care and showed evidence of a beneficial effect of rhG-CSF on all-cause mortality (*p* = 0.017), and receipt of mechanical ventilation (*p* = 0.003) [85]. This trial also showed evidence of a harmful effect of rhG-CSF on non-serious adverse events (*p* = 0.0001) [85]. One single trial randomizing 84 participants compared electrolyzed saline versus standard care and showed evidence of a beneficial effect of electrolyzed saline on all-cause mortality (*p* = 0.019) [100]. One single trial randomizing 78 participants compared bromhexidine hydrochloride versus standard care and showed evidence of a beneficial effect of bromhexidine hydrochloride on admittance to intensive care (*p* = 0.013) and receipt of mechanical ventilation (*p* = 0.014) [104]. One single trial randomizing 100 participants compared hydroxychloroquine combined with arbidol versus hydroxychloroquine combined with lopinavir-ritonavir and showed evidence of a beneficial effect of hydroxychloroquine combined with arbidol on admission to intensive care (*p =* 0.0001) [109].

None of the remaining single trial results showed evidence of a difference on our predefined review outcomes. Two trials did not report the results in a usable way; one trial reported results of the experimental group with a proportion of participants being non-randomized [56], and the second trial reported the results as per-protocol, and there was participant crossover [42]. Seven trials did not report on our review outcomes [44,62,63,67,71,96,101]. We have contacted all corresponding authors, but we have not been able to obtain outcomes for our analyses from the trialists yet. Most trials were assessed at high risk of bias (**S3 Table**). Characteristics of the trials and their results on the review outcomes can be found in **S2 Table**. Certainty of the evidence was assessed as ‘low’ or ‘very low’ for all outcomes (**S15-S66 Table**).

### Possible future contributions of ongoing trials

On November 2, 2020, a search on the Cochrane COVID-19 Study Register revealed 2527 registered randomized clinical trials [13]. From these, 106 different interventions for treatment of COVID-19 patients were identified [13]. The ten most investigated experimental interventions were hydroxychloroquine (162 trials), convalescent plasma (55 trials), azithromycin (52 trials), lopinavir and ritonavir (40 trials), tocilizumab (33 trials), chloroquine (30 trials), favipiravir (24 trials), remdesivir (15 trials), sarilumab (15 trials), and dexamethasone (13 trials). Eligible trials will continuously be included in the present living systematic review once results become available.

## Discussion

We conducted the second edition of our living systematic review assessing the beneficial and harmful effects of any intervention for COVID-19. We searched relevant databases and websites for published and unpublished trials until November 2, 2020. We included a total of 82 trials randomizing 40,249 participants. Our study showed that no evidence-based treatment for COVID-19 currently exists.

Very low certainty evidence indicated that corticosteroids might reduce the risk of death, serious adverse events, and mechanical ventilation.

Moderate certainty evidence showed that we could reject that remdesivir reduces the risk of death by 20%. Very low certainty evidence indicated that remdesivir might reduce the risk of serious adverse events.

Very low certainty evidence indicated that intraveneous immunoglobin might reduce the risk of death and serious adverse events, that tocilizumab might reduce the risk of serious adverse events and mechanical ventilation, and that bromhexidine might reduce the risk of non-serious adverse events.

Moderate certainty evidence showed that we could reject that hydroxychloroquine reduces the risk of death and serious adverse events by 20%, and that we could reject that lopinavir-ritonavir reduces the risk of death, serious adverse events, and risk of mechanical ventilation by 20%.

Otherwise, we could neither confirm nor reject the effects of other interventions for COVID-19. More trials with low risks of bias and random errors are urgently needed. For several interventions we found a large number of currently ongoing trials.

The present review concludes that no evidence-based treatment currently exists for COVID-19. Previous studies [118–120] including our first edition of the present review [120] have concluded that both corticosteroids and remdesivir showed promising results. Since our last edition, we have included more trials, and we therefore have more information, causing this difference, but there are other reasons for these contrary conclusions. One previous systematic review published in JAMA assessed the association between corticosteroids and 28-day all-cause mortality and concluded that corticosteroids are effective for treating critically ill patients with COVID-19 in reducing all-cause mortality [118]. This review assessed the certainty of evidence for all-cause mortality to be moderate, while we assessed the certainty of evidence to be very low.

Our present results showed a discrepancy between the random-effects meta-analysis result and the fixed-effect meta-analysis result (due to heterogeneity) of corticosteroids versus control interventions when assessing all-cause mortality, i.e., the fixed-effect meta-analysis indicated a more beneficial effect of corticosteroids. Due to the discrepancy between the random-effects and the fixed-effect model we believe that these results should be interpreted with great caution considering the uncertainty of the evidence. Furthermore, the meta-analytic effect estimate was 0.89 which may be considered relatively small.

The U.S. Food and Drug Administration (FDA) recently approved remdesivir for use in adult and pediatric patients 12 years of age and older for the treatment of COVID-19 requiring hospitalization [121]. Based on the current evidence, we conclude that remdesivir is not effective in reducing all-cause mortality, neither for patients not requiring oxygen nor for patients requiring oxygen or respiratory support at baseline. There was a discrepancy between the random-effects and the fixed-effect meta-analysis (due to heterogeneity) of remdesivir versus control on serious adverse events, i.e., the fixed-effect meta-analysis indicated a more beneficial effect of remdesivir. Due to the discrepancy between the random-effects and the fixed-effect model we believe that these results should be interpreted with great caution considering the uncertainty of the evidence. On all other outcomes when assessing the effects of remdesivir, we conclude that more information is needed to confirm or reject the effects of remdesivir. Hence, the clinical effects of remdesivir are unclear based on current evidence.

Our results are similar to the results of a preprint of an international collaborative meta-analysis of randomized clinical trials assessing mortality outcomes with hydroxychloroquine and chloroquine for participants with COVID-19 [122]. This review included some unpublished data [122]. We have contacted the trialists of the trials that provided unpublished data for this review, but we have not received any data yet. Nevertheless, our conclusions that hydroxychloroquine does not reduce mortality in COVID-19 patients are the same [122].

Although we could exclude an intervention effect at 20% or above for most of our interventions with our trial sequential analyses, we did not assess smaller and still worthwhile intervention effects. If patients and investigators feel that such smaller intervention effects are worth pursuing, then we recommend the conduct of trials with much larger sample sizes than the ones we have identified in the present systematic review. That will require more national and international collaboration [123].

Our living systematic review has a number of strengths. The predefined methodology was based on the Cochrane Handbook for Systematic Reviews of Interventions [11], the eight-step assessment suggested by Jakobsen et al. [17], and trial sequential analysis [22]. Hence, this review considers both risks of systematic errors and risk of random errors. Another strength is the living systematic review design, which allows us to continuously surveil and update the evidence-base of existing interventions for treatment of COVID-19 resulting in a decreased timespan from publication of our results to optimization of clinical practice. This is particularly important in this international health-care crisis, where a large number of new randomized clinical trials are continuously registered and published.

Our living systematic review also has limitations. First, the primary limitation is the paucity of trials currently available, and the results from most current meta-analyses are of low or very low certainty. This must be considered when interpreting our meta-analysis results. Second, the trials that we included were all at risks of systematic errors so our results presumably overestimate the beneficial effects and underestimate the harmful effects of the included interventions [124–131]. Third, it was not relevant to perform individual patient data meta-analyses, network-meta-analysis, or several of the planned subgroup analyses due to lack of relevant data. We contacted all trial authors requesting individual patient data, but until now we only received five datasets [38,69,79,80,107]. We did not perform network meta-analysis because the ranking of the interventions is not unclear, i.e., no evidence-based intervention currently exits for COVID-19. Fourth, we included ’time to clinical improvement’ as an outcome post hoc. We did not initially plan to assess ‘time to clinical improvement’ [8] because this outcome is poorly defined and if outcome assessors are not adequately blinded, assessments of ‘improvement’ may be biased. Furthermore, time to clinical improvement is not one of the most patient-important outcomes. As an example, most patients would rather survive without complications than recover a few days sooner. Fifth, the included trials assessed the outcomes at different time points, which might contribute to increased heterogeneity. Sixth, some data are included from preprints, and these might be subject to change following peer-review. Therefore, some results, bias risk assessments, and GRADE summaries might change in later editions of this living systematic review following inclusion of the published peer-reviewed manuscripts.

We have identified two important reviews that are comparable to our present project [119, 132]. The first is a network meta-analysis published in BMJ [119]. However, this review only includes drug treatments for COVID-19, does not include individual patient data meta-analyses, and does not use trial sequential analysis or similar methods to handle problems with multiplicity (repeating updating of meta-analysis, multiple comparisons due to inclusion of multiple interventions, assessing multiple outcomes).

The second project is a living mapping of ongoing randomized clinical trials with network meta-analysis on all interventions for COVID-19 [132]. The authors are producing and disseminating preliminary results through an open platform [132]. This review includes both prevention and treatment and does not use trial sequential analysis or similar methods to handle problems with multiplicity (repeating updating of meta-analysis, multiple comparisons due to inclusion of multiple interventions, assessing multiple outcomes) [8].

## Conclusions

No evidence-based treatment for COVID-19 currently exists. Very low certainty evidence indicates that corticosteroids might reduce the risk of death, serious adverse events, and mechanical ventilation; that remdesivir might reduce the risk of serious adverse events; that intraveneous immunoglobin might reduce the risk of death and serious adverse events; that tocilizumab might reduce the risk of serious adverse events and mechanical ventilation, and that bromhexidine might reduce the risk of non-serious adverse events. More trials with low risks of bias and random errors are urgently needed. This review will continuously inform best practice in treatment and clinical research of COVID-19.

## Differences between the protocol and the review

We erroneously reported the adjusted TSA alpha as 2% in our published protocol [8]. This has now been corrected to 3.3% according to two primary outcomes [17]. Further, we included ‘time to clinical improvement’ as an outcome post hoc.

## Data Availability

The data underlying the results presented in the study are available from the secretary at Copenhagen Trial Unit, Mette Hansen. She is not an author of this paper. If we receive individual patient data from a third party, there may apply some restrictions.

## Acknowledgements

Not applicable

## Supporting information captions

### Text

S1 Text: PRISMA 2009 Checklist.

S2 Text: Search strategies.

### Tables

S1 Table. Excluded trials.

S2 Table. Characteristics of included studies.

S3 Table. Risk of bias assessments.

S4 Table. Summary of Findings table of corticosteroids versus control interventions (standard care or placebo)

S5 Table. Summary of Findings table of remdesivir versus control interventions (standard care or placebo)

S6 Table. Summary of Findings table of hydroxychloroquine versus control interventions (standard care or placebo)

S7 Table. Summary of Findings table of lopinavir-ritonavir versus control interventions (standard care or placebo)

S8 Table. Summary of Findings table of interferon beta-1a versus standard care

S9 Table. Summary of Findings table of convalescent plasma versus control interventions (standard care or placebo)

S10 Table. Summary of Findings table of azithromycin versus control interventions (standard care or placebo)

S11 Table. Summary of Findings table of colchicine versus control interventions (standard care or placebo)

S12 Table. Summary of Findings table of intraveneous immunoglobin versus control interventions (standard care or placebo)

S13 Table. Summary of Findings table of tocilizumab versus control interventions (standard care or placebo)

S14 Table. Summary of Findings table of bromhexine versus control interventions (standard care or placebo)

S15 Table. Summary of Findings table of favipiravir versus control interventions (standard care or placebo)

S16 Table. Summary of Findings table of favipiravir versus umifenovir

S17 Table. Summary of Findings table of umifenovir versus lopinavir-ritonavir

S18 Table. Summary of Findings table of umifenovir versus standard care

S19 Table. Summary of Findings table of novaferon versus novaferon + lopinavir-ritonavir

S20 Table. Summary of Findings table of novaferon + lopinavir-ritonavir versus lopinavir-ritonavir

S21 Table. Summary of Findings table of novaferon versus lopinavir-ritonavir

S22 Table. Summary of Findings table of alpha lipotic acid versus placebo

S23 Table. Summary of Findings table of baloxavir marboxil versus favipavir

S24 Table. Summary of Findings table of baloxavir marboxil versus standard care

S25 Table. Summary of Findings table of triple combination of interferon beta-1b + lopinavir-ritonavir + ribavirin versus lopinavir-ritonavir

S26 Table. Summary of Findings table of remdesivir for 10 days versus remdesivir for 5 days

S27 Table. Summary of Findings table of high-flow nasal oxygenation versus standard bag-valve oxygenation

S28 Table. Summary of Findings table of hydroxychloroquine versus chloroquine

S29 Table. Summary of Findings table of chloroquine versus standard care

S30 Table. Summary of Findings table of high dosage chloroquine diphosphate versus low dosage chloroquine diphosphate

S31 Table. Summary of Findings table of hydroxychloroquine + azithromycin versus standard care

S32 Table. Summary of Findings table of triple combination of darunavir + cobicistat + interferon alpha-2b versus interferon alpha-2b

S33 Table. Summary of Findings table of lopinavir-ritonavir + interferon alpha versus ribavirin + interferon alpha

S34 Table. Summary of Findings table of ribavirin + lopinavir-ritonavir + interferon alpha versus ribavirin + interferon alpha

S35 Table. Summary of Findings table of ribavirin + lopinavir-ritonavir + interferon alpha versus lopinavir-ritonavir + interferone alpha

S36 Table. Summary of Findings table of Lincocin® versus Azitro®

S37 Table. Summary of Findings table of ^99m^Tc-MDP injection versus standard care

S38 Table. Summary of Findings table of interferon alpha-2b + interferon gamma versus interferone alpha-2b

S39 Table. Summary of Findings table of telmisartan versus standard care

S40 Table. Summary of Findings table of avifavir 1600/600 versus avifavir 1800/800

S41 Table. Summary of Findings table of dexamethasone + aprepitant versus dexamethasone

S42 Table. Summary of Findings table of anti-C5a antibody versus standard care

S43 Table. Summary of Findings table of azvudine versus standard care

S44 Table. Summary of Findings table of human plasma-derived C1 esterase/kallikrein inhibitor versus standard care

S45 Table. Summary of Findings table of icatibant acetate versus standard care

S46 Table. Summary of Findings table of icatibant acetate versus human plasma-derived C1 esterase/kallikrein inhibitor

S47 Table. Summary of Findings table of pulmonary rehabilitation program versus isolation at home

S48 Table. Summary of Findings table of auxora (calcium release-activated calcium channel inhibitors) versus standard care

S49 Table. Summary of Findings table of umbilical cord stem cell infusion versus standard care

S50 Table. Summary of Findings table of vitamin C versus placebo

S51 Table. Summary of Findings table of sofosbuvir + daclatasvir versus standard care

S52 Table. Summary of Findings table of sofosbuvir + daclatasvir + ribavirin versus hydroxychloroquine + lopinavir-ritonavir with or without ribavirin

S53 Table. Summary of Findings table of interferon beta-1b versus standard care S54 Table. Summary of Findings table of calcifediol versus standard care

S55 Table. Summary of Findings table of rhG-CSF versus standard care

S56 Table. Summary of Findings table of intravenous and/or nebulized electrolyzed saline with dose escalation versus standard care

S57 Table. Summary of Findings table of nasal irrigation with hypertonic saline + surfactant versus no intervention

S58 Table. Summary of Findings table of nasal irrigation with hypertonic saline versus nasal irrigation with hypertonic saline + surfactant

S59 Table. Summary of Findings table of nasal irrigation with hypertonic saline versus no intervention

S60 Table. Summary of Findings table of triazavirin versus placebo

S61 Table. Summary of Findings table of N-acetylcysteine versus placebo

S62 Table. Summary of Findings table of hydroxychloroquine + arbidiol versus hydroxychloroquine + lopinavir-ritonavir

S63 Table. Summary of Findings table of tocilizumab versus favipiravir

S64 Table. Summary of Findings table of avifavir 1600/600 versus standard care

S65 Table. Summary of Findings table of avifavir 1800/800 versus standard care

S66 Table. Summary of Findings table of tocilizumab + favipiravir versus tocilizumab

### Figures

S1 Fig. Fixed-effect meta-analysis of corticosteroids versus control interventions (standard care or placebo) on all-cause mortality.

S2 Fig. Subgroup analysis of disease severity for corticosteroids versus control interventions (standard care or placebo) on all-cause mortality

S3 Fig. Random-effects meta-analysis of corticosteroids versus control interventions (standard care or placebo) on serious adverse events.

S4 Fig. Fixed-effect meta-analysis of corticosteroids versus control interventions (standard care or placebo) on serious adverse events.

S5 Fig. Trial Sequential Analysis of corticosteroids versus control interventions (standard care or placebo) on serious adverse events.

S6 Fig. Random-effects meta-analysis of corticosteroids versus control interventions (standard care or placebo) on receipt of mechanical ventilation.

S7 Fig. Fixed-effect meta-analysis of corticosteroids versus control interventions (standard care or placebo) on receipt of mechanical ventilation.

S8 Fig. Trial Sequential Analysis of corticosteroids versus control interventions (standard care or placebo) on receipt of mechanical ventilation.

S9 Fig. Subgroup analysis of early versus late intervention for remdesivir versus control interventions (standard care or placebo) on all-cause mortality.

S10 Fig. Random-effects meta-analysis of remdesivir versus control interventions (standard care or placebo) on serious adverse events.

S11 Fig. Fixed-effect meta-analysis of remdesivir versus control interventions (standard care or placebo) on serious adverse events.

S12 Fig. Trial Sequential Analysis of remdesivir versus control interventions (standard care or placebo) on serious adverse events.

S13 Fig. Random-effects meta-analysis of remdesivir versus control interventions (standard care or placebo) on receipt of mechanical ventilation.

S14 Fig. Trial Sequential Analysis of remdesivir versus control interventions (standard care or placebo) on receipt of mechanical ventilation

S15 Fig. Fixed-effect meta-analysis of remdesivir versus control interventions (standard care or placebo) on non-serious adverse events.

S16 Fig. Trial Sequential Analysis of remdesivir versus control interventions (standard care or placebo) on non-serious adverse events.

S17 Fig. Fixed-effect meta-analysis of hydroxychloroquine versus control interventions (standard care or placebo) on all-cause mortality.

S18 Fig. Trial Sequential Analysis of hydroxychloroquine versus control interventions (standard care or placebo) on all-cause mortality.

S19 Fig. Fixed-effect meta-analysis of hydroxychloroquine versus control interventions (standard care or placebo) on serious adverse events.

S20 Fig. Trial Sequential Analysis of hydroxychloroquine versus control interventions (standard care or placebo) on serious adverse events.

S21 Fig. Fixed-effect meta-analysis of hydroxychloroquine versus control interventions (standard care or placebo) on admission to intensive care.

S22 Fig. Trial Sequential Analysis of hydroxychloroquine versus control interventions (standard care or placebo) on admission to intensive care.

S23 Fig. Fixed-effect meta-analysis of hydroxychloroquine versus control interventions (standard care or placebo) on receipt of mechanical ventilation.

S24 Fig. Trial Sequential Analysis of hydroxychloroquine versus control interventions (standard care or placebo) on receipt of mechanical ventilation.

S25 Fig. Random-effects meta-analysis of hydroxychloroquine versus control interventions (standard care or placebo) on non-serious adverse events.

S26 Fig. Fixed-effect meta-analysis of lopinavir-ritonavir versus control interventions (standard care or placebo) on all-cause mortality.

S27 Fig. Trial Sequential Analysis of lopinavir-ritonavir versus control interventions (standard care or placebo) on all-cause mortality.

S28 Fig. Random-effects meta-analysis of lopinavir-ritonavir versus control interventions (standard care or placebo) on serious adverse events.

S29 Fig. Trial Sequential Analysis of lopinavir-ritonavir versus control interventions (standard care or placebo) on serious adverse events.

S30 Fig. Random-effects meta-analysis of lopinavir-ritonavir versus control interventions (standard care or placebo) on receipt of mechanical ventilation

S31 Fig. Trial Sequential Analysis of lopinavir-ritonavir versus control interventions (standard care or placebo) on receipt of mechanical ventilation

S32 Fig. Random-effects meta-analysis of lopinavir-ritonavir versus control interventions (standard care or placebo) on receipt of renal replacement therapy

S33 Fig. Trial Sequential Analysis of lopinavir-ritonavir versus control interventions on receipt of renal replacement therapy

S34 Fig. Fixed-effect meta-analysis of lopinavir-ritonavir versus control interventions (standard care or placebo) on non-serious adverse events

S35 Fig. Random-effects meta-analysis of interferon β-1a versus control interventions (standard care or placebo) on all-cause mortality

S36 Fig. Random-effects meta-analysis of interferon β-1a versus control interventions (standard care or placebo) on serious adverse events

S37 Fig. Random-effects meta-analysis of convalescent plasma versus control interventions (standard care or placebo) on all-cause mortality

S38 Table. Trial Sequential Analysis of convalescent plasma versus control interventions (standard care or placebo) on all-cause mortality.

S39 Fig. Fixed-effect meta-analysis of convalescent plasma versus control interventions (standard care or placebo) on serious adverse events.

S40 Fig. Trial Sequential Analysis of convalescent plasma versus control interventions (standard care or placebo) on serious adverse events.

S41 Fig. Fixed-effect meta-analysis of azithromycin versus control interventions (standard care or placebo) on all-cause mortality

S42 Fig. Trial Sequential Analysis of azithromycin versus control interventions (standard care or placebo) on all-cause mortality.

S43 Fig. Random-effects meta-analysis of azithromycin versus control interventions (standard care or placebo) on serious adverse events.

S44 Fig. Fixed-effect meta-analysis of azithromycin versus control interventions (standard care or placebo) on receipt of mechanical ventilation.

S45 Fig. Fixed-effect meta-analysis of azithromycin versus control interventions (standard care or placebo) on non-serious adverse events

S46 Fig. Trial Sequential Analysis of azithromycin versus control interventions (standard care or placebo) on non-serious adverse events.

S47 Fig. Fixed-effect meta-analysis of colchicine versus control interventions (standard care or placebo) on all-cause mortality.

S48 Fig. Random-effects meta-analysis of colchicine versus control interventions (standard care or placebo) on non-serious adverse events.

S49 Fig. Fixed-effect meta-analysis of intraveneous immunoglobin versus control interventions (standard care or placebo) on all-cause mortality.

S50 Fig. Trial Sequential Analysis of intraveneous immunoglobin versus control interventions (standard care or placebo) on all-cause mortality

S51 Fig. Fixed-effect meta-analysis of intraveneous immunoglobin versus control interventions on serious adverse events.

S52 Fig. Trial Sequential Analysis of intraveneous immunoglobin versus control interventions (standard care or placebo) on all-cause mortality.

S53 Fig. Random-effects meta-analysis of tocilizumab versus control interventions (standard care or placebo) on all-cause mortality.

S54 Fig. Trial Sequential Analysis of tocilizumab versus control interventions (standard care or placebo) on all-cause mortality.

S55 Fig. Random-effects meta-analysis of tocilizumab versus control interventions (standard care, placebo, or a co-intervention alone) on serious adverse events.

S56 Fig. Fixed-effect meta-analysis of tocilizumab versus control interventions (standard care, placebo, or a co-intervention alone) on serious adverse events.

S57 Fig. Random-effects meta-analysis of tocilizumab versus control interventions (standard care or placebo) on admission to intensive care.

S58 Fig. Random-effects meta-analysis of tocilizumab versus control interventions (standard care or placebo) on mechanical ventilation.

S59 Fig. Trial Sequential Analysis of tocilizumab versus control interventions (standard care or placebo) on mechanical ventilation.

S60 Fig. Fixed-effect meta-analysis of tocilizumab versus control interventions (standard care, placebo, or a co-intervention alone) on non-serious adverse events.

S61 Fig. Random-effects meta-analysis of bromhexidine versus control interventions (standard care) on all-cause mortality.

S62 Fig. Random-effects meta-analysis of bromhexidine versus control interventions (standard care) on non-serious adverse events.

