## Supplementary material for "Interventions for treatment of COVID-19: second edition of a living systematic review with meta-analyses and trial sequential analyses (The LIVING Project)": S19 Fig.pdf

| Study | Hydroxychloroquine Events | Hydroxychloroquine No Events | Control Events | Control No Events | Risk Ratio with 95% CI | Weight (%) |
|---|---|---|---|---|---|---|
| Hydroxychloroquine vs. standard care | | | | | | |
| Abd-Elsalam 2020 | 6 | 91 | 5 | 92 | 1.20 [ 0.38, 3.80] | 0.78 |
| Cavalcanti 2020a | 7 | 214 | 6 | 221 | 1.20 [ 0.41, 3.51] | 0.92 |
| Chen CP 2020 | 0 | 19 | 0 | 11 | 0.60 [ 0.01, 28.31] | 0.10 |
| Chen J 2020 | 0 | 15 | 0 | 15 | 1.00 [ 0.02, 47.38] | 0.08 |
| Chen L 2020b | 0 | 18 | 0 | 12 | 0.68 [ 0.01, 32.34] | 0.09 |
| Lyngbakken 2020 | 5 | 21 | 6 | 19 | 0.80 [ 0.28, 2.29] | 0.95 |
| Mitja 2020 | 8 | 161 | 12 | 172 | 0.73 [ 0.30, 1.73] | 1.78 |
| RECOVERY (hydroxychloroquine) 2020 | 418 | 1,143 | 788 | 2,367 | 1.07 [ 0.97, 1.19] | 80.91 |
| SOLIDARITY (hydroxychloroquine) 2020 | 104 | 843 | 84 | 822 | 1.18 [ 0.90, 1.56] | 13.32 |
| Heterogeneity: $I^2 = 0.00\%$, $H^2 = 1.00$ | | | | | 1.08 [ 0.98, 1.19] | |
| Test of $\theta_i = \theta_j$: Q(8) = 1.78, p = 0.99 | | | | | | |
| Hydroxychloroquine vs. placebo | | | | | | |
| Skipper 2020 | 1 | 230 | 1 | 233 | 1.01 [ 0.06, 16.10] | 0.15 |
| Ulrich 2020 | 7 | 43 | 6 | 44 | 1.17 [ 0.42, 3.23] | 0.93 |
| Heterogeneity: $I^2 = 0.00\%$, $H^2 = 1.00$ | | | | | 1.14 [ 0.44, 2.98] | |
| Test of $\theta_i = \theta_j$: Q(1) = 0.01, p = 0.93 | | | | | | |
| **Overall** | | | | | 1.08 [ 0.98, 1.19] | |
| Heterogeneity: $I^2 = 0.00\%$, $H^2 = 1.00$ | | | | | | |
| Test of $\theta_i = \theta_j$: Q(10) = 1.81, p = 1.00 | | | | | | |
| Test of group differences: $Q_b(1) = 0.01$, p = 0.90 | | | | | | |

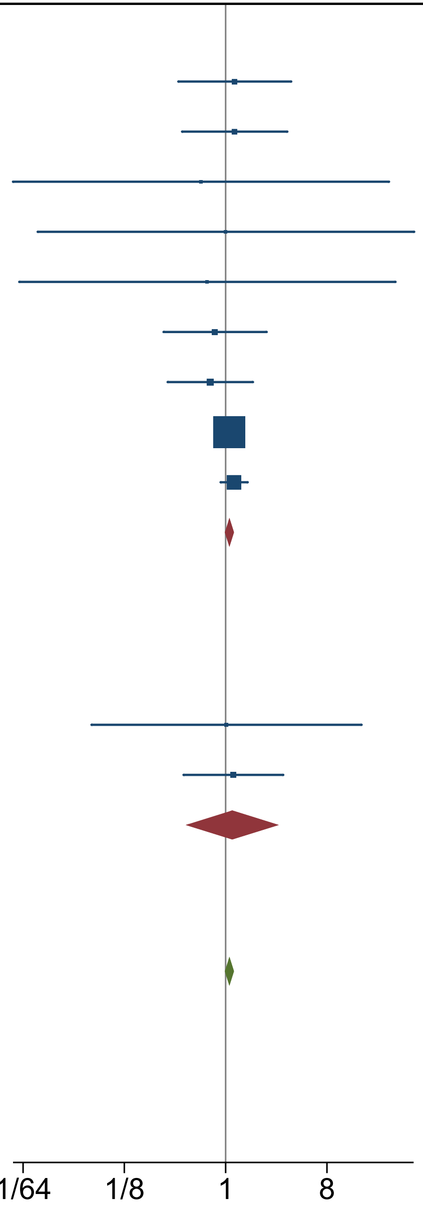


Fixed-effects Mantel-Haenszel model
