## Supplementary material for "Interventions for treatment of COVID-19: second edition of a living systematic review with meta-analyses and trial sequential analyses (The LIVING Project)": S21 Fig.pdf

| Study | Hydroxychloroquine Events | Hydroxychloroquine No Events | Control Events | Control No Events | Risk Ratio with 95% CI | Weight (%) |
|---|---|---|---|---|---|---|
| Hydroxychloroquine vs. standard care | | | | | | |
| Abd-Elsalam 2020 | 11 | 86 | 13 | 84 | 0.85 [ 0.40, 1.79] | 48.15 |
| Heterogeneity: $I^2$ = 0.00%, $H^2$ = 1.00 | | | | | 0.85 [ 0.40, 1.79] | |
| Test of $\theta_i = \theta_j$: Q(0) = 0.00, p = . | | | | | | |
| Hydroxychloroquine vs. placebo | | | | | | |
| Ulrich 2020 | 9 | 41 | 14 | 36 | 0.64 [ 0.31, 1.35] | 51.85 |
| Heterogeneity: $I^2$ = 0.00%, $H^2$ = 1.00 | | | | | 0.64 [ 0.31, 1.35] | |
| Test of $\theta_i = \theta_j$: Q(0) = 0.00, p = . | | | | | | |
| **Overall** | | | | | 0.74 [ 0.44, 1.25] | |
| Heterogeneity: $I^2$ = 0.00%, $H^2$ = 1.00 | | | | | | |
| Test of $\theta_i = \theta_j$: Q(1) = 0.26, p = 0.61 | | | | | | |
| Test of group differences: $Q_b(1)$ = 0.26, p = 0.61 | | | | | | |

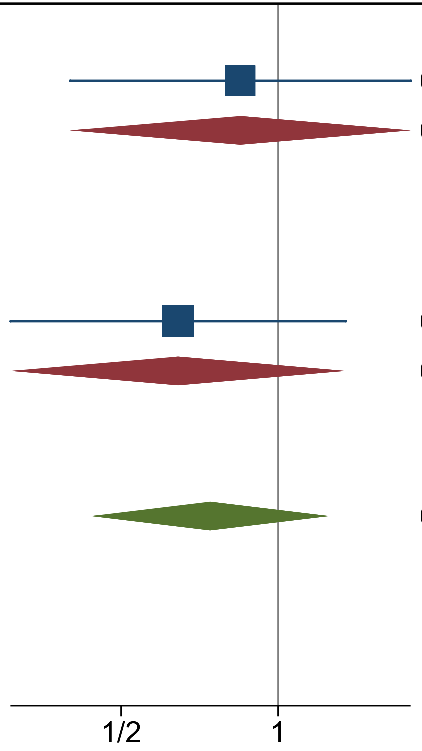


Fixed-effects Mantel-Haenszel model
