## Supplementary material for "Interventions for treatment of COVID-19: second edition of a living systematic review with meta-analyses and trial sequential analyses (The LIVING Project)": S23 Fig.pdf

| Study | Hydroxychloroquine Events | Hydroxychloroquine No Events | Control Events | Control No Events | Risk Ratio with 95% CI | Weight (%) |
|---|---|---|---|---|---|---|
| Hydroxychloroquine vs. standard care | | | | | | |
| Abd-Elsalam 2020 | 4 | 93 | 5 | 92 | 0.80 [ 0.22, 2.89] | 5.60 |
| Cavalcanti 2020a | 16 | 205 | 13 | 214 | 1.26 [ 0.62, 2.57] | 14.36 |
| SOLIDARITY (hydroxychloroquine) 2020 | 75 | 787 | 66 | 758 | 1.09 [ 0.79, 1.49] | 75.56 |
| Heterogeneity: $I^2 = 0.00\%$, $H^2 = 1.00$ | | | | | 1.10 [ 0.83, 1.45] | |
| Test of $\theta_i = \theta_j$: Q(2) = 0.39, p = 0.82 | | | | | | |
| Hydroxychloroquine vs. placebo | | | | | | |
| Ulrich 2020 | 5 | 45 | 4 | 46 | 1.25 [ 0.36, 4.38] | 4.48 |
| Heterogeneity: $I^2 = 0.00\%$, $H^2 = 1.00$ | | | | | 1.25 [ 0.36, 4.38] | |
| Test of $\theta_i = \theta_j$: Q(0) = 0.00, p = . | | | | | | |
| **Overall** | | | | | 1.10 [ 0.84, 1.45] | |
| Heterogeneity: $I^2 = 0.00\%$, $H^2 = 1.00$ | | | | | | |
| Test of $\theta_i = \theta_j$: Q(3) = 0.43, p = 0.93 | | | | | | |
| Test of group differences: $Q_b(1) = 0.04$, p = 0.84 | | | | | | |

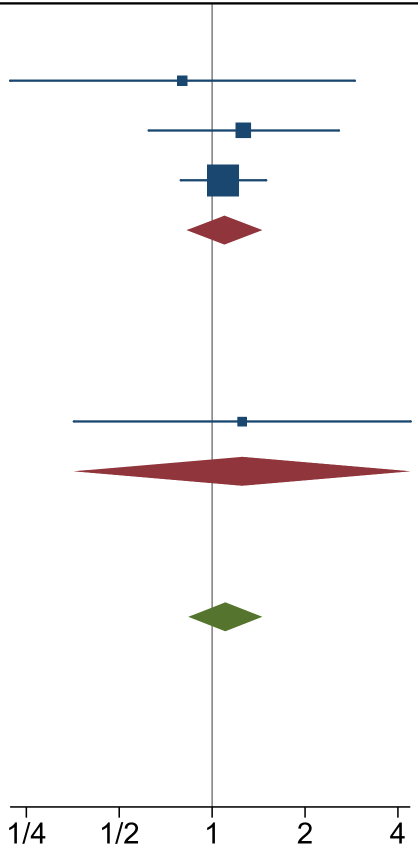


Fixed-effects Mantel-Haenszel model
