## Supplementary material for "Interventions for treatment of COVID-19: second edition of a living systematic review with meta-analyses and trial sequential analyses (The LIVING Project)": S25 Fig.pdf

| Study | Hydroxychloroquine Events | Hydroxychloroquine No Events | Control Events | Control No Events | Risk Ratio with 95% CI | Weight (%) |
|---|---|---|---|---|---|---|
| Hydroxychloroquine vs. standard care | | | | | | |
| Cavalcanti 2020a | 74 | 147 | 61 | 166 | 1.25 [ 0.94, 1.66] | 20.16 |
| Chen J 2020 | 4 | 11 | 3 | 13 | 1.42 [ 0.38, 5.33] | 10.44 |
| Chen L 2020b | 9 | 9 | 2 | 10 | 3.00 [ 0.78, 11.54] | 10.23 |
| Mitja 2020 | 121 | 48 | 16 | 168 | 8.23 [ 5.11, 13.28] | 18.62 |
| Heterogeneity: $\tau^2 = 1.33$, $I^2 = 93.33\%$, $H^2 = 14.98$ | | | | | 2.63 [ 0.77, 8.91] | |
| Test of $\theta_i = \theta_j$: Q(3) = 44.95, p = 0.00 | | | | | | |
| Hydroxychloroquine vs. placebo | | | | | | |
| Skipper 2020 | 92 | 120 | 46 | 165 | 1.99 [ 1.48, 2.68] | 20.07 |
| Ulrich 2020 | 38 | 12 | 36 | 14 | 1.06 [ 0.84, 1.33] | 20.47 |
| Heterogeneity: $\tau^2 = 0.18$, $I^2 = 90.74\%$, $H^2 = 10.80$ | | | | | 1.44 [ 0.77, 2.68] | |
| Test of $\theta_i = \theta_j$: Q(1) = 10.80, p = 0.00 | | | | | | |
| **Overall** | | | | | 2.09 [ 1.14, 3.80] | |
| Heterogeneity: $\tau^2 = 0.44$, $I^2 = 92.10\%$, $H^2 = 12.66$ | | | | | | |
| Test of $\theta_i = \theta_j$: Q(5) = 63.29, p = 0.00 | | | | | | |
| Test of group differences: $Q_b(1) = 0.74$, p = 0.39 | | | | | | |

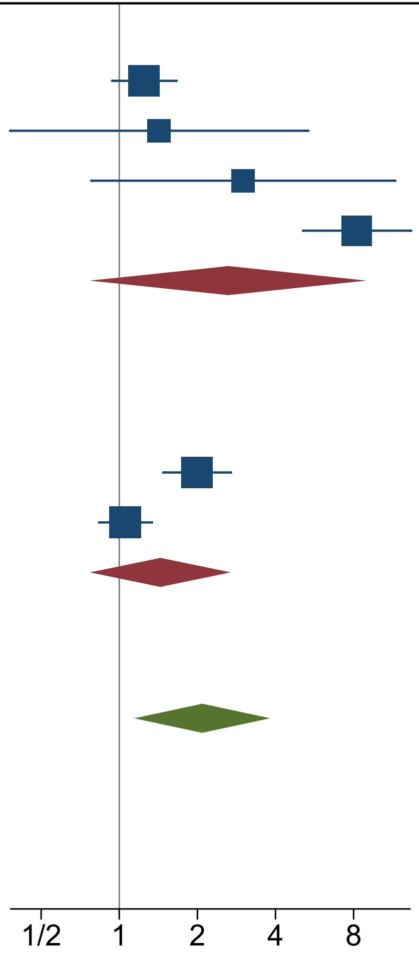


Random-effects DerSimonian-Laird model
