## Supplementary material for "Interventions for treatment of COVID-19: second edition of a living systematic review with meta-analyses and trial sequential analyses (The LIVING Project)": S37 Fig.pdf

| Study | Convalescent plasma Events | Convalescent plasma No Events | Standard care Events | Standard care No Events | Risk Ratio with 95% CI | Weight (%) |
|---|---|---|---|---|---|---|
| Agarwal 2020 | 34 | 201 | 31 | 198 | 1.07 [ 0.68, 1.68] | 49.75 |
| Avendaño-Solà 2020 | 0 | 37 | 4 | 35 | 0.12 [ 0.01, 2.10] | 2.74 |
| Gharbharan 2020 | 6 | 37 | 11 | 32 | 0.55 [ 0.22, 1.34] | 21.82 |
| Li 2020 | 8 | 43 | 12 | 38 | 0.65 [ 0.29, 1.46] | 25.69 |
| **Overall** | | | | | 0.77 [ 0.47, 1.24] | |

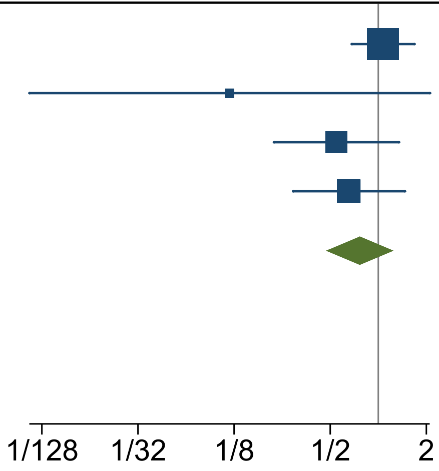


Heterogeneity: $\tau^2 = 0.07$, $I^2 = 27.54\%$, $H^2 = 1.38$

Test of $\theta_i = \theta_j$: $Q(3) = 4.14$, $p = 0.25$

Test of $\theta = 0$: $z = -1.08$, $p = 0.28$

Random-effects DerSimonian-Laird model
