## Supplementary material for "Interventions for treatment of COVID-19: second edition of a living systematic review with meta-analyses and trial sequential analyses (The LIVING Project)": S39 Fig.pdf

| Study | Convalescent plasma Events | Convalescent plasma No Events | Standard care Events | Standard care No Events | Risk Ratio with 95% CI | Weight (%) |
|---|---|---|---|---|---|---|
| Agarwal 2020 | 34 | 201 | 31 | 198 | 1.07 [ 0.68, 1.68] | 63.80 |
| Avendaño-Solà 2020 | 6 | 31 | 7 | 32 | 0.90 [ 0.33, 2.44] | 13.85 |
| Gharbharan 2020 | 6 | 37 | 11 | 32 | 0.55 [ 0.22, 1.34] | 22.35 |
| **Overall** | | | | | 0.93 [ 0.64, 1.35] | |

Heterogeneity: $I^2 = -16.50\%$, $H^2 = 0.86$

Test of $\theta_i = \theta_j$: $Q(2) = 1.72$, $p = 0.42$

Test of $\theta = 0$: $z = -0.39$, $p = 0.70$

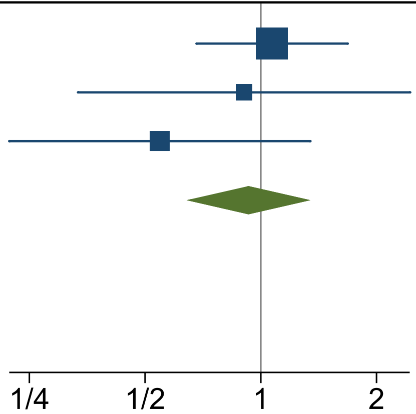


Fixed-effects Mantel-Haenszel model
