## Supplementary material for "Interventions for treatment of COVID-19: second edition of a living systematic review with meta-analyses and trial sequential analyses (The LIVING Project)": S44 Fig.pdf

| Study | Azithromycin Events | Azithromycin No Events | Control Events | Control No Events | Risk Ratio with 95% CI | Weight (%) |
|---|---|---|---|---|---|---|
| Azithromycin vs. azithromycin + LPV/r + HCQ | | | | | | |
| Sekhavati 2020 | 0 | 56 | 3 | 52 | 0.14 [ 0.01, 2.66] | 18.22 |
| Heterogeneity: $I^2 = 0.00\%$, $H^2 = 1.00$ | | | | | 0.14 [ 0.01, 2.66] | |
| Test of $\theta_i = \theta_j$: Q(0) = 0.00, p = . | | | | | | |
| Azithromycin + HCQ + standard care vs. HCQ + standard care | | | | | | |
| Cavalcanti 2020c | 20 | 197 | 16 | 205 | 1.27 [ 0.68, 2.39] | 81.78 |
| Heterogeneity: $I^2 = 0.00\%$, $H^2 = 1.00$ | | | | | 1.27 [ 0.68, 2.39] | |
| Test of $\theta_i = \theta_j$: Q(0) = 0.00, p = . | | | | | | |
| **Overall** | | | | | 1.07 [ 0.59, 1.94] | |
| Heterogeneity: $I^2 = 53.06\%$, $H^2 = 2.13$ | | | | | | |
| Test of $\theta_i = \theta_j$: Q(1) = 2.13, p = 0.14 | | | | | | |
| Test of group differences: $Q_b(1) = 2.07$, p = 0.15 | | | | | | |

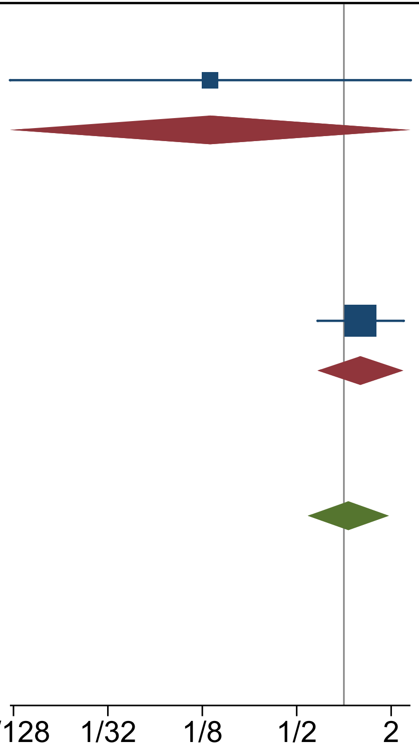


Fixed-effects Mantel-Haenszel model
