## Supplementary material for "Interventions for treatment of COVID-19: second edition of a living systematic review with meta-analyses and trial sequential analyses (The LIVING Project)": S47 Fig.pdf

| Study | Colchicine Events | Colchicine No Events | Control Events | Control No Events | Risk Ratio with 95% CI | Weight (%) |
|---|---|---|---|---|---|---|
| Colchicine + standard care vs. placebo + standard care | | | | | | |
| Lopes 2020 | 0 | 17 | 0 | 18 | 1.06 [ 0.02, 50.43] | 49.32 |
| Heterogeneity: $I^2 = 0.00\%$, $H^2 = 1.00$ | | | | | 1.06 [ 0.02, 50.43] | |
| Test of $\theta_i = \theta_j$: Q(0) = 0.00, p = . | | | | | | |
| Colchicine + HCQ vs. Placebo + HCQ | | | | | | |
| Salehzadeh 2020 | 0 | 50 | 0 | 50 | 1.00 [ 0.02, 49.44] | 50.68 |
| Heterogeneity: $I^2 = 0.00\%$, $H^2 = 1.00$ | | | | | 1.00 [ 0.02, 49.44] | |
| Test of $\theta_i = \theta_j$: Q(0) = 0.00, p = . | | | | | | |
| **Overall** | | | | | 1.03 [ 0.07, 16.01] | |
| Heterogeneity: $I^2 = 0.00\%$, $H^2 = 1.00$ | | | | | | |
| Test of $\theta_i = \theta_j$: Q(1) = 0.00, p = 0.98 | | | | | | |

Test of group differences: $Q_b(1) = 0.00$, $p = 0.98$

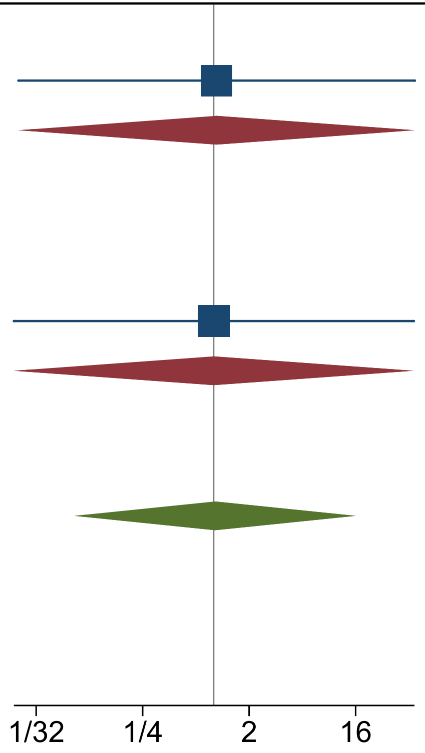


Fixed-effects Mantel-Haenszel model
