## Supplementary material for "Interventions for treatment of COVID-19: second edition of a living systematic review with meta-analyses and trial sequential analyses (The LIVING Project)": S49 Fig.pdf

| Study | IVIG Event | IVIG No event | Control Event | Control No event | Risk Ratio with 95% CI | Weight (%) |
|---|---|---|---|---|---|---|
| **Placebo + standard care** | | | | | | |
| Gharebaghi 2020 | 6 | 24 | 14 | 15 | 0.41 [ 0.18, 0.93] | 83.03 |
| Heterogeneity: $I^2$ = 0.00%, $H^2$ = 1.00 | | | | | 0.41 [ 0.18, 0.93] | |
| Test of $\theta_i = \theta_j$: Q(0) = 0.00, p = . | | | | | | |
| **Standard care** | | | | | | |
| Sakoulas 2020 | 1 | 15 | 3 | 14 | 0.35 [ 0.04, 3.06] | 16.97 |
| Heterogeneity: $I^2$ = 100.00%, $H^2$ = 1.00 | | | | | 0.35 [ 0.04, 3.06] | |
| Test of $\theta_i = \theta_j$: Q(0) = 0.00, p = . | | | | | | |
| **Overall** | | | | | 0.40 [ 0.19, 0.87] | |
| Heterogeneity: $I^2$ = 0.00%, $H^2$ = 1.00 | | | | | | |
| Test of $\theta_i = \theta_j$: Q(1) = 0.02, p = 0.89 | | | | | | |
| Test of group differences: $Q_b(1)$ = 0.02, p = 0.89 | | | | | | |

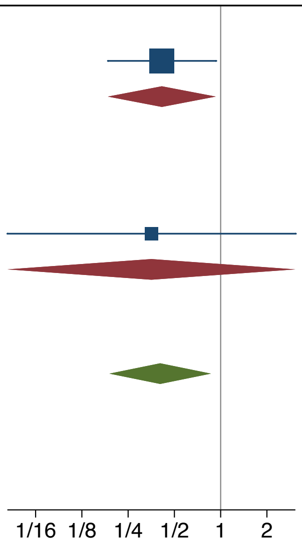


Fixed-effects Mantel-Haenszel model
