## Supplementary material for "Interventions for treatment of COVID-19: second edition of a living systematic review with meta-analyses and trial sequential analyses (The LIVING Project)": S61 Fig.pdf

| Study | Bromhexidine Event | Bromhexidine No event | Control Event | Control No event | Risk Ratio with 95% CI | Weight (%) |
|---|---|---|---|---|---|---|
| Ansarin 2020 | 0 | 39 | 5 | 34 | 0.09 [ 0.01, 1.59] | 63.94 |
| Li T 2020 | 0 | 12 | 0 | 6 | 0.54 [ 0.01, 24.33] | 36.06 |
| **Overall** | | | | | 0.17 [ 0.02, 1.70] | |

Heterogeneity: $\tau^2 = 0.00$, $I^2 = 0.00\%$, $H^2 = 1.00$

Test of $\theta_i = \theta_j$: Q(1) = 0.54, p = 0.46

Test of $\theta = 0$: z = -1.50, p = 0.13

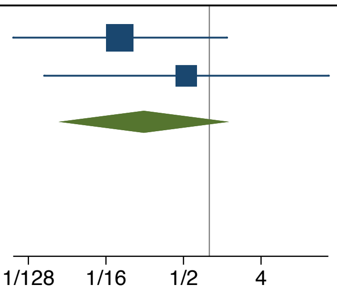


Random-effects DerSimonian-Laird model
