## Supplementary material for "Interventions for treatment of COVID-19: second edition of a living systematic review with meta-analyses and trial sequential analyses (The LIVING Project)": S62 Fig.pdf

| Study | Bromhexidine Event | Bromhexidine No event | Control Event | Control No event | Risk Ratio with 95% CI | Weight (%) |
|---|---|---|---|---|---|---|
| Ansarin 2020 | 4 | 35 | 14 | 25 | 0.29 [ 0.10, 0.79] | 55.24 |
| Li T 2020 | 3 | 9 | 4 | 2 | 0.38 [ 0.12, 1.16] | 44.76 |
| **Overall** | | | | | 0.32 [ 0.15, 0.69] | |

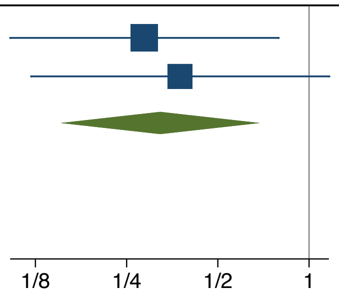


Heterogeneity: $\tau^2 = 0.00$, $I^2 = 0.00\%$, $H^2 = 1.00$

Test of $\theta_i = \theta_j$: Q(1) = 0.12, p = 0.73

Test of $\theta = 0$: z = -2.93, p = 0.00

Random-effects DerSimonian-Laird model
