## Supplementary material for "Interventions for treatment of COVID-19: second edition of a living systematic review with meta-analyses and trial sequential analyses (The LIVING Project)": S1 Table.docx

### Excluded trials:

| Trial ID | Reason for exclusion |
| --- | --- |
| Abella 2020 [1] | Wrong population (preexposure prophylaxis) |
| Boulware 2020 [2] | Wrong population (postexposure prophylaxis) |
| Carlucci 2020 [3] | Not randomised |
| Casadevall 2020 [4] | Not randomised |
| Cattaneo 2020 [5] | Not randomised |
| ChiCTR2000029954 2020 [6] | Wrong intervention (Chinese medicine) |
| Christensen 2020 [7] | Wrong intervention, and wrong population (teaching) |
| Clariot 2020 [8] | Wrong population (Simulation study, not on patients) |
| Davoodi [9] | Wrong population (suspected COVID-19) |
| Demidowich 2020 [10] | Wrong population (Non covid-19) |
| Deng 2020 [11] | Not randomised |
| Duan 2020 [12] | Not randomised |
| El-Lababidi 2020 [13] | Not randomised |
| Gautret 2020 [14] | Not randomised |
| Gong 2020 [15] | Not randomised |
| Goyal 2020 [16] | Not randomised |
| Grein 2020 [17] | Not randomised |
| Hu K (1) 2020 [18] | Wrong intervention (Chinese medicine) |
| Hu K (2) 2020 [19] | Wrong intervention (Chinese medicine) |
| Liu K 2020 [20] | Not randomised |
| Liu ST 2020 [21] | Not randomised |
| Magagnoli 2020 [22] | Not randomised |
| Mahevas 2020 [23] | Not randomised |
| Meng 2020 [24] | Not randomised |
| Menzella 2020 [25] | Not randomised |
| Milne 2020 [26] | Wrong population (Non covid-19) |
| Pielacinski 2020 [27] | Wrong population (Non covid-19) |
| Ramiro 2020 [28] | Not randomised |
| Salazar 2020 [29] | Not randomised |
| Somers 2020 [30] | Not randomised |
| Wang 2020 [31] | Wrong intervention (Chinese medicine) |
| Xiao 2020 [32] | Wrong intervention (Chinese medicine) |
| Xiong 2020 [33] | Wrong intervention (Chinese medicine) |
| Xu 2020 [34] | Not randomised |

6.	ChiCTR2000029954. Efficacy and safety of honeysuckle oral liquid in the treatment of novel coronavirus pneumonia (COVID-19): a multicenter, randomized, controlled, open clinical trial. ICTRP. 2020. PubMed PMID: 13841926.

15.	Gong Y, Guan L, Jin Z, Chen S, Xiang G, Gao B. Effects of methylprednisolone use on viral genomic nucleic acid negative conversion and CT imaging lesion absorption in COVID-19 patients under 50 years old. Journal of medical virology. 2020. PubMed PMID: 13842697; PubMed Central PMCID: PMC631853763.

16.	Goyal DK, Mansab F, Iqbal A, Bhatti S. Early intervention likely improves mortality in COVID-19 infection. Clinical medicine (London, England). 2020. PubMed PMID: 13842719; PubMed Central PMCID: PMC631801518.

17.	Grein J, Ohmagari N, Shin D, Diaz G, Asperges E, Castagna A, et al. Compassionate Use of Remdesivir for Patients with Severe Covid-19. New england journal of medicine 2020 apr 10. 2020. doi: 10.1056/NEJMoa2007016. PubMed PMID: 13380846.

18.	Hu K, Guan WJ, Bi Y, Zhang W, Li L, Zhang B, et al. Efficacy and safety of Lianhuaqingwen capsules, a repurposed Chinese herb, in patients with coronavirus disease 2019: a multicenter, prospective, randomized controlled trial. Phytomedicine. 2020;(no pagination). PubMed PMID: 13842694; PubMed Central PMCID: PMC2005976431.

19.	Hu K, Wang MM, Zhao Y, Zhang YT, Wang T, Zheng ZS, et al. A Small-Scale Medication of Leflunomide as a Treatment of COVID-19 in an Open-Label Blank-Controlled Clinical Trial. Virologica Sinica. doi: 10.1007/s12250-020-00258-7. PubMed PMID: 14477295.
