## Supplementary material for "Interventions for treatment of COVID-19: second edition of a living systematic review with meta-analyses and trial sequential analyses (The LIVING Project)": S4 Table.docx

S4 Summary of Findings

| **Glucocorticosteroids compared with standard care for COVID-19** | | | | | | |
| --- | --- | --- | --- | --- | --- | --- |
| **Patients or population:** Anyone with a diagnosis of COVID-19  **Setting:** Any setting  **Intervention:** Glucocorticosteroids  **Comparison:** Standard care | | | | | | |
| **Outcomes** | **Anticipated absolute effects* (95% CI)** | | **Relative effect (95% CI)** | **No of participants (studies)** | **Certainty of the evidence (GRADE)** | **Comments** |
|  | **Risk with standard care** | **Risk with**  **glucocorticosteroids** |  |  |  |  |
| **All-cause mortality**  *Follow-up mean: 24,5 days* | 277 per 1,000 | **247 per 1,000**  (227 to 277) | **RR 0.89** (0.79 to 1.00) | 7,795  (9 RCTs) | ⨁◯◯◯ VERY LOW ^a,b,c^ | - |
| **Serious adverse events** *Follow-up mean: 24,5 days* | 281 per 1,000 | **250 per 1,000**  (225 to 278) | **RR 0.89** (0.80 to 0.99) | 7,795  (9 RCTs) | ⨁◯◯◯ VERY LOW ^a,b^ | - |
| **Admission to intensive care** | - | - | - | - | - | Outcome not yet measured or reported |
| **Mechanical ventilation** *Follow-up mean: 17.5 days* | 81 per 1,000 | **70 per 1,000**  (44 to 108) | **RR 0.86** (0.55 to 1.33) | 5,614  (2 RCTs) | ⨁◯◯◯ VERY LOW ^a,b^ | - |
| **Renal replacement therapy** | - | - | - | - | - | Outcome not yet measured or reported |
| **Quality of Life** | - | - | - | - | - | Outcome not yet measured or reported |
| **Non-serious adverse events** | - | - | - | - | - | Outcome not yet measured or reported |
| *The risk in the intervention group (and its 95% confidence interval) is based on the assumed risk in the comparison group and the relative effect of the intervention (and its 95% CI).  **RR:** Risk ratio; **CI:** Confidence interval; **GRADE:** GRADE Working Group grades of evidence | | | | | | |
| **GRADE Working Group grades of evidence**  **High certainty:** We are very confident that the true effect lies close to that of the estimate of the effect **Moderate certainty:** We are moderately confident in the effect estimate: The true effect is likely to be close to the estimate of the effect, but there is a possibility that it is substantially different **Low certainty:** Our confidence in the effect estimate is limited: The true effect may be substantially different from the estimate of the effect **Very low certainty:** We have very little confidence in the effect estimate: The true effect is likely to be substantially different from the estimate of effect | | | | | | |

**Explanations**

a. Downgraded 2 for risk of bias

b. Downgraded 1 for imprecision due to Trial Sequential Analysis showing that there was not enough information to confirm or reject a relative risk reduction (RRR) of 20%.
c. Downgraded 1 for indirectness due to inconsistency between fixed-effect and random-effect meta-analysis.
