## Supplementary material for "Interventions for treatment of COVID-19: second edition of a living systematic review with meta-analyses and trial sequential analyses (The LIVING Project)": S5 Table.docx

S5 Summary of Findings

| **Remdesivir compared with placebo for COVID-19** | | | | | | |
| --- | --- | --- | --- | --- | --- | --- |
| **Patients or population:** Anyone with a diagnosis of COVID-19  **Setting:** Any setting  **Intervention:** Remdesivir  **Comparison:** Placebo | | | | | | |
| **Outcomes** | **Anticipated absolute effects* (95% CI)** | | **Relative effect (95% CI)** | **No of participants (studies)** | **Certainty of the evidence (GRADE)** | **Comments** |
|  | **Risk with placebo** | **Risk with**  **remdesivir** |  |  |  |  |
| **All-cause mortality**  *Follow-up: mean 21 days* | 113 per 1,000 | **105 per 1,000**  (92 to 121) | **RR 0.93** (0.82 to 1.07) | 7,310  (5 RCTs) | ⨁⨁⨁⨁ HIGH | - |
| **Serious adverse events**  *Follow-up: mean 21 days* | 144 per 1,000 | **118 per 1,000**  (98 to 144) | **RR 0.82** (0.68 to 1.00) | 7,313  (5 RCTs) | ⨁◯◯◯ VERY LOW ^a,b,c^ | - |
| **Admission to intensive care** | - | - | - | - | - | Outcome not yet measured or reported |
| **Mechanical ventilation** *Follow-up: mean 21 days* | 127 per 1,000 | **92 per 1,000**  (53 to 161) | **RR 0.73** (0.42 to 1.27) | 5,957  (3 RCTs) | ⨁◯◯◯ VERY LOW ^a,b,d^ | Outcome not yet measured or reported |
| **Renal replacement therapy** | - | - | - | - | - | Outcome not yet measured or reported |
| **Quality of Life** | - | - | - | - | - | Outcome not yet measured or reported |
| **Non-serious adverse events**  *Follow-up: mean 21 days* | 552 per 1,000 | **546 per 1,000**  (502 to 596) | **RR 0.99** (0.91 to 1.08) | 1,865  (4 RCTs) | ⨁◯◯◯ VERY LOW ^a,d^ | - |
| *The risk in the intervention group (and its 95% confidence interval) is based on the assumed risk in the comparison group and the relative effect of the intervention (and its 95% CI).  **RR:** Risk ratio; **CI:** Confidence interval; **GRADE:** GRADE Working Group grades of evidence | | | | | | |
| **GRADE Working Group grades of evidence**  **High certainty:** We are very confident that the true effect lies close to that of the estimate of the effect **Moderate certainty:** We are moderately confident in the effect estimate: The true effect is likely to be close to the estimate of the effect, but there is a possibility that it is substantially different **Low certainty:** Our confidence in the effect estimate is limited: The true effect may be substantially different from the estimate of the effect **Very low certainty:** We have very little confidence in the effect estimate: The true effect is likely to be substantially different from the estimate of effect | | | | | | |

**Explanations**

a. Downgraded 2 for risk of bias

b. Downgraded 2 for imprecision due to Trial Sequential Analysis showing that there was not enough information to confirm or reject a relative risk reduction (RRR) of 20%.
c. Downgraded 1 for indirectness due to inconsistency between fixed-effect and random-effect meta-analysis.

d. Downgraded 1 for inconsistency due to heterogenity.
