## Supplementary material for "Interventions for treatment of COVID-19: second edition of a living systematic review with meta-analyses and trial sequential analyses (The LIVING Project)": S6 Table.docx

S6 Summary of Findings

| **Hydroxychloroquine compared with standard care for COVID-19** | | | | | | |
| --- | --- | --- | --- | --- | --- | --- |
| **Patients or population:** Anyone with a diagnosis of COVID-19  **Setting:** Any setting  **Intervention:** Hydroxychloroquine  **Comparison:** Standard care | | | | | | |
| **Outcomes** | **Anticipated absolute effects* (95% CI)** | | **Relative effect (95% CI)** | **No of participants (studies)** | **Certainty of the evidence (GRADE)** | **Comments** |
|  | **Risk with standard care** | **Risk with**  **hydroxychloroquine** |  |  |  |  |
| **All-cause mortality**  *Follow-up: mean 18.3 days* | 182 per 1,000 | **198 per 1,000**  (180 to 218) | **1.09**  (0.99 to 1.20) | 8219  (10 RCTs) | ⨁⨁⨁◯ MODERATE ^a^ | - |
| **Serious adverse events**  *Follow-up: mean 23.5 days* | 185 per 1,000 | **200 per 1,000**  (181 to 220) | **1.08**  (0.98 to 1.19) | 8240  (10 RCTs) | ⨁⨁⨁◯ MODERATE ^a^ | - |
| **Admission to intensive care** *Follow-up: mean 28 days* | 184 per 1,000 | **136 per 1,000**  (81 to 230) | **0.74**  (0.44 to 1.25) | 294  (2 RCTs) | ⨁◯◯◯ VERY LOW ^b,c^ | - |
| **Mechanical ventilation** *Follow-up: mean 23.7 days* | 73 per 1,000 | **81 per 1,000**  (62 to 107) | **1.10**  (0.84 to 1.45) | 2428  (4 RCTs) | ⨁◯◯◯ VERY LOW ^b,c,e^ | - |
| **Renal replacement therapy** | - | - | - | - | - | Outcome not yet measured or reported |
| **Quality of Life** | - | - | - | - | - | Outcome not yet measured or reported |
| **Non-serious adverse events**  *Follow-up: mean 16.2 days* | 234 per 1,000 | **490 per 1,000**  (267 to 890) | **RR 2.09** (1.14 to 3.80) | 1385  (6 RCTs) | ⨁◯◯◯ VERY LOW ^b,c,d,e^ | - |
| *The risk in the intervention group (and its 95% confidence interval) is based on the assumed risk in the comparison group and the relative effect of the intervention (and its 95% CI).  **RR:** Risk ratio; **CI:** Confidence interval; **GRADE:** GRADE Working Group grades of evidence | | | | | | |
| **GRADE Working Group grades of evidence**  **High certainty:** We are very confident that the true effect lies close to that of the estimate of the effect **Moderate certainty:** We are moderately confident in the effect estimate: The true effect is likely to be close to the estimate of the effect, but there is a possibility that it is substantially different **Low certainty:** Our confidence in the effect estimate is limited: The true effect may be substantially different from the estimate of the effect **Very low certainty:** We have very little confidence in the effect estimate: The true effect is likely to be substantially different from the estimate of effect | | | | | | |

**Explanations**

a. Downgraded 1 for risk of bias

b. Downgraded 2 for risk of bias

c. Downgraded 2 for imprecision due to Trial Sequential Analysis showing that there was not enough information to confirm or reject a relative risk reduction (RRR) of 20%. Moreover, the meta-analysis showed wide CI.

d. Downgraded 2 for inconsistency due to large heterogeneity (I^2^ > 90%).

e. Downgraded 1 for publication bias/for profit bias due to two trials being funded by a pharmaceutical company
