## Supplementary material for "Interventions for treatment of COVID-19: second edition of a living systematic review with meta-analyses and trial sequential analyses (The LIVING Project)": S7 Table.docx

S7 Summary of Findings

| **Lopinavir-ritonavir compared with standard care for COVID-19** | | | | | | |
| --- | --- | --- | --- | --- | --- | --- |
| **Patients or population:** Anyone with a diagnosis of COVID-19  **Setting:** Any setting  **Intervention:** Lopinavir-ritonavir  **Comparison:** Standard care | | | | | | |
| **Outcomes** | **Anticipated absolute effects* (95% CI)** | | **Relative effect (95% CI)** | **No of participants (studies)** | **Certainty of the evidence (GRADE)** | **Comments** |
|  | **Risk with standard care** | **Risk with**  **lopinavir-ritonavir** |  |  |  |  |
| **All-cause mortality**  *Follow-up: mean 21.5 days* | 190 per 1,000 | **192 per 1,000** (175 to 213) | **RR 1.01** (0.92 to 1.12) | 8121  (5 RCTs) | ⨁⨁⨁◯ MODERATE ^a^ | Two trials reported zero events |
| **Serious adverse events**  *Follow-up: mean 21.5 days* | 191 per 1,000 | **191 per 1,000**  (174 to 212) | **RR 1.00** (0.91 to 1.11) | 8116  (5 RCTs) | ⨁⨁⨁◯ MODERATE ^a^ | One trial reported zero events |
| **Admission to intensive care** | - | - | - | - | - | Outcome not yet measured or reported |
| **Mechanical ventilation** *Follow-up: mean 28 days* | 90 per 1,000 | **97 per 1,000** (84 to 112) | **RR 1.08** (0.94 to 1.25) | 7575 (3 RCTs) | ⨁⨁⨁◯ MODERATE ^a^ | - |
| **Renal replacement therapy** *Follow-up: mean 28 days* | 42 per 1,000 | **41 per 1,000** (31 to 54) | **RR 0.97** (0.73 to 1.28) | 5130 (2 RCTs) | ⨁◯◯◯ VERY LOW ^b,c^ | - |
| **Quality of Life** | - | - | - | - | - | Outcome not yet measured or reported |
| **Non-serious adverse events**  *Follow-up: mean 24.5 days* | 422 per 1,000 | **449 per 1,000**  (359 to 646) | **RR 1.14** (0.85 to 1.53) | 245  (2 RCTs) | ⨁◯◯◯ VERY LOW ^b,c,d^ | - |
| *The risk in the intervention group (and its 95% confidence interval) is based on the assumed risk in the comparison group and the relative effect of the intervention (and its 95% CI).  **RR:** Risk ratio; **CI:** Confidence interval; **GRADE:** GRADE Working Group grades of evidence | | | | | | |
| **GRADE Working Group grades of evidence**  **High certainty:** We are very confident that the true effect lies close to that of the estimate of the effect **Moderate certainty:** We are moderately confident in the effect estimate: The true effect is likely to be close to the estimate of the effect, but there is a possibility that it is substantially different **Low certainty:** Our confidence in the effect estimate is limited: The true effect may be substantially different from the estimate of the effect **Very low certainty:** We have very little confidence in the effect estimate: The true effect is likely to be substantially different from the estimate of effect | | | | | | |

**Explanations**

a. Downgraded 1 for risk of bias
b. Downgraded 2 for risk of bias

c. Downgraded 2 for imprecision due to Trial Sequential Analysis showing that there was not enough information to confirm or reject a relative risk reduction (RRR) of 20%. Moreover, the meta-analysis showed wide CI.

d. Downgraded 2 for inconsistency due to large heterogeneity
