## Supplementary material for "Interventions for treatment of COVID-19: second edition of a living systematic review with meta-analyses and trial sequential analyses (The LIVING Project)": S11 Table.docx

S11 Summary of Findings

| **Colchicine compared with standard care for COVID-19** | | | | | | |
| --- | --- | --- | --- | --- | --- | --- |
| **Patients or population:** Anyone with a diagnosis of COVID-19  **Setting:** Any setting  **Intervention:** Colchicine  **Comparison:** Standard care | | | | | | |
| **Outcomes** | **Anticipated absolute effects* (95% CI)** | | **Relative effect (95% CI)** | **No of participants (studies)** | **Certainty of the evidence (GRADE)** | **Comments** |
|  | **Risk with standard care** | **Risk with**  **colchicine** |  |  |  |  |
| **All-cause mortality** *Follow-up mean: unclear* | 0 per 1,000 | **0 per 1,000** | **RR 1.03** (0.07 to 16.01) | 135 (2 RCT) | ⨁◯◯◯ VERY LOW ^a,b^ | Both trials reported zero mortality in both groups. |
| **Serious adverse events**  *Follow-up: unclear* | 20 per 1,000 | **20 per 1,000-** | - | 100 (1 RCT) | ⨁◯◯◯ VERY LOW ^a,e,f^ | - |
| **Admission to intensive care** *Follow-up: unclear* | 56 per 1,000 | **59 per 1,000** | - | 35 (1 RCT) | ⨁◯◯◯ VERY LOW ^a,e,f^ | - |
| **Mechanical ventilation** | **-** | **-** | - | - | - | Outcome not yet measured or reported |
| **Renal replacement therapy** | - | - | - | - | - | Outcome not yet measured or reported |
| **Quality of Life** | - | - | - | - | - | Outcome not yet measured or reported |
| **Non-serious adverse events**  *Follow-up: 21 days* | 195 per 1,000 | **172 per 1,000** (35 to 856) | **RR 0.88** (0.18 to 4.39) | 248 (3 RCT) | ⨁◯◯◯ VERY LOW ^a,b,c,d^ | - |
| *The risk in the intervention group (and its 95% confidence interval) is based on the assumed risk in the comparison group and the relative effect of the intervention (and its 95% CI).  **RR:** Rate ratio for all-cause mortality, and risk ratio for mechanical ventilation; **CI:** Confidence interval; **GRADE:** GRADE Working Group grades of evidence | | | | | | |
| **GRADE Working Group grades of evidence**  **High certainty:** We are very confident that the true effect lies close to that of the estimate of the effect **Moderate certainty:** We are moderately confident in the effect estimate: The true effect is likely to be close to the estimate of the effect, but there is a possibility that it is substantially different **Low certainty:** Our confidence in the effect estimate is limited: The true effect may be substantially different from the estimate of the effect **Very low certainty:** We have very little confidence in the effect estimate: The true effect is likely to be substantially different from the estimate of effect | | | | | | |

**Explanations**

a. Downgraded 2 for risk of bias
b. Downgraded 2 for imprecision due to Trial Sequential Analysis showing that there was not enough information to confirm or reject a relative risk reduction (RRR) of 20%. Moreover, meta-analysis showed wide CI. There were few participants and few events.
c. Downgraded 2 due to visual inspection of forest plot showing heterogenity and I^2^=79.19%
d. Downgraded 1 for publication bias/for profit bias due to one study being funded by a pharmaceutical company
e. Downgraded 1 for indirectness due to a single study from a single country, therefore results in this population might not be generalizable to other settings
f. Downgraded 2 for imprecision due to low number of participants and events
