## Supplementary material for "Interventions for treatment of COVID-19: second edition of a living systematic review with meta-analyses and trial sequential analyses (The LIVING Project)": S13 Table.docx

S13 Summary of Findings

| **Tocilizumab compared with standard care for COVID-19** | | | | | | |
| --- | --- | --- | --- | --- | --- | --- |
| **Patients or population:** Anyone with a diagnosis of COVID-19  **Setting:** Any setting  **Intervention:** Tocilizumab  **Comparison:** Standard care | | | | | | |
| **Outcomes** | **Anticipated absolute effects* (95% CI)** | | **Relative effect (95% CI)** | **No of participants (studies)** | **Certainty of the evidence (GRADE)** | **Comments** |
|  | **Risk with standard care** | **Risk with**  **Tocilizumab** |  |  |  |  |
| **All-cause mortality** *Follow-up mean: 29 days* | 115 per 1,000 | **119 per 1,000** (83 to 169) | **RR 1.03** (0.72 to 1.46) | 933  (4 RCTs) | ⨁◯◯◯ VERY LOW ^a,b,c^ | - |
| **Serious adverse events** *Follow-up mean: 30 days* | 378 per 1,000 | **238 per 1,000** (132 to 431) | **RR 0.63** (0.35 to 1.14) | 955  (5 RCTs) | ⨁◯◯◯ VERY LOW ^a,b,c,d^ | - |
| **Admission to intensive care** *Follow-up mean: 30 days* | 244 per 1,000 | **173 per 1,000** (90 to 337) | **RR 0.71** (0.37 to 1.38) | 314  (2 RCTs) | ⨁◯◯◯ VERY LOW ^a,b,c^ | - |
| **Mechanical ventilation** *Follow-up mean: Unclear* | 230 per 1,000 | **161 per 1,000** (117 to 221) | **RR 0.70** (0.51 to 0.96) | 646  (3 RCTs) | ⨁◯◯◯ VERY LOW ^a,b,c^ | - |
| **Renal replacement therapy** | - | - | - | - | - | Outcome not yet measured or reported |
| **Quality of Life** | - | - | - | - | - | Outcome not yet measured or reported |
| **Non-serious adverse events** *Follow-up mean: Unclear* | 527 per 1,000 | **543 per 1,000**  (485 to 601) | **RR 1.03** (0.92 to 1.14) | 777  (5 RCTs) | ⨁◯◯◯ VERY LOW ^a,b,c,d,e^ | - |
| *The risk in the intervention group (and its 95% confidence interval) is based on the assumed risk in the comparison group and the relative effect of the intervention (and its 95% CI).  **RR:** Risk ratio; **CI:** Confidence interval; **GRADE:** GRADE Working Group grades of evidence | | | | | | |
| **GRADE Working Group grades of evidence**  **High certainty:** We are very confident that the true effect lies close to that of the estimate of the effect **Moderate certainty:** We are moderately confident in the effect estimate: The true effect is likely to be close to the estimate of the effect, but there is a possibility that it is substantially different **Low certainty:** Our confidence in the effect estimate is limited: The true effect may be substantially different from the estimate of the effect **Very low certainty:** We have very little confidence in the effect estimate: The true effect is likely to be substantially different from the estimate of effect | | | | | | |

**Explanations**

a. Downgraded 2 for risk of bias

b. Downgraded 2 for imprecision due to Trial Sequential Analysis showing that there was not enough information to confirm or reject a relative risk reduction (RRR) of 20%.

c. Downgraded 1 due to publication bias/for profit bias as 1 trial received funding from the pharmaceutical company supplying the intervention.

d. Downgraded 2 for inconsistency due to heterogeneity.

e. Downgraded 1 for indirectness due to differences in subgroup analysis.
