## Supplementary material for "Interventions for treatment of COVID-19: second edition of a living systematic review with meta-analyses and trial sequential analyses (The LIVING Project)": S15 Table.docx

S15 Summary of Findings

| **Favipiravir compared with standard care for COVID-19** | | | | | | |
| --- | --- | --- | --- | --- | --- | --- |
| **Patients or population:** Anyone with a diagnosis of COVID-19  **Setting:** Any setting  **Intervention:** Favipiravir  **Comparison:** Standard care | | | | | | |
| **Outcomes** | **Anticipated absolute effects* (95% CI)** | | **Relative effect (95% CI)** | **No of participants (studies)** | **Certainty of the evidence (GRADE)** | **Comments** |
|  | **Risk with**  **standard care** | **Risk with**  **favipiravir** |  |  |  |  |
| **All-cause mortality**  *Follow-up: 14 days* | 0 per 1.000 | **0 per 1.000** | - | 19  (1 RCT) | ⨁◯◯◯ VERY LOW ^a,b,c^ | Zero events in both groups. Zero numbers analyzed in two studies. |
| **Serious adverse events**  *Follow-up: 14 days* | 400 per 1.000 | **444 per 1.000** | - | 19  (1 RCT) | ⨁◯◯◯ VERY LOW ^a,b,c^ | Zero numbers analyzed in two studies. |
| **Admission to intensive care**  *Follow-up: 14 days* | 0 per 1.000 | **222 per 1.000** | - | 19  (1 RCT) | ⨁◯◯◯ VERY LOW ^a,b,c^ | Zero numbers analyzed in two studies. |
| **Mechanical ventilation**  *Follow-up: 14 days* | 0 per 1.000 | **0 per 1.000** | - | 19  (1 RCT) | ⨁◯◯◯ VERY LOW ^a,b,c^ | Zero events in both groups. Zero numbers analyzed in two studies. |
| **Renal replacement therapy** | - | - | - | - | - | Outcome not yet measured or reported |
| **Quality of Life** | - | - | - | - | - | Outcome not yet measured or reported |
| **Non-serious adverse events**  *Follow-up: 14 days* | 900 per 1.000 | **889 per 1.000** | - | 19  (1 RCT) | ⨁◯◯◯ VERY LOW ^a,b,c^ | Zero numbers analyzed in two studies. |
| **RR:** Risk ratio; **CI:** Confidence interval; **GRADE:** GRADE Working Group grades of evidence | | | | | | |
| **GRADE Working Group grades of evidence**  **High certainty:** We are very confident that the true effect lies close to that of the estimate of the effect **Moderate certainty:** We are moderately confident in the effect estimate: The true effect is likely to be close to the estimate of the effect, but there is a possibility that it is substantially different **Low certainty:** Our confidence in the effect estimate is limited: The true effect may be substantially different from the estimate of the effect **Very low certainty:** We have very little confidence in the effect estimate: The true effect is likely to be substantially different from the estimate of effect | | | | | | |

**Explanations**

a. Downgraded 2 for risk of bias

b. Downgraded 2 for imprecision due to low number of participants

c. Downgraded 2 for publication bias due to two trials having no published results.
