## Supplementary material for "Interventions for treatment of COVID-19: second edition of a living systematic review with meta-analyses and trial sequential analyses (The LIVING Project)": S26 Table.docx

S26 Summary of Findings

| **Remdesivir for five days compared with Remdesivir for ten days for COVID-19** | | | | | | |
| --- | --- | --- | --- | --- | --- | --- |
| **Patients or population:** Anyone with a diagnosis of COVID-19  **Setting:** Any setting  **Intervention:** Five days remdesivir  **Comparison:** Ten days remdesivir | | | | | | |
| **Outcomes** | **Anticipated absolute effects* (95% CI)** | | **Relative effect (95% CI)** | **No of participants (studies)** | **Certainty of the evidence (GRADE)** | **Comments** |
|  | **Risk with**  **ten days** | **Risk with**  **five days** |  |  |  |  |
| **All-cause mortality**  *Follow-up: 30 days* | 107 per 1,000 | **80 per 1,000** | - | 402  (1 RCT) | ⨁◯◯◯ VERY LOW ^a,b,c^ | - |
| **Serious adverse events**  *Follow-up: 30 days* | 345 per 1,000 | **210 per 1,000** | - | 402  (1 RCT) | ⨁◯◯◯ VERY LOW ^a,b,c^ | - |
| **Admission to intensive care** | - | - | - | - | - | Outcome not yet measured or reported |
| **Mechanical ventilation**  *Follow-up: 30 days* | - | **-** | - | - | - | Outcome not yet measured or reported |
| **Renal replacement therapy** | - | - | - | - | - | Outcome not yet measured or reported |
| **Quality of Life** | - | - | - | - | - | Outcome not yet measured or reported |
| **Non-serious adverse events** | - | **-** | - | - | - | Outcome not yet measured or reported |
| **RR:** Risk ratio; **CI:** Confidence interval; **GRADE:** GRADE Working Group grades of evidence | | | | | | |
| **GRADE Working Group grades of evidence**  **High certainty:** We are very confident that the true effect lies close to that of the estimate of the effect **Moderate certainty:** We are moderately confident in the effect estimate: The true effect is likely to be close to the estimate of the effect, but there is a possibility that it is substantially different **Low certainty:** Our confidence in the effect estimate is limited: The true effect may be substantially different from the estimate of the effect **Very low certainty:** We have very little confidence in the effect estimate: The true effect is likely to be substantially different from the estimate of effect | | | | | | |

**Explanations**

a. Downgraded 2 for risk of bias

b. Downgraded 1 for imprecision due to low number of participants
