## Supplementary material for "Interventions for treatment of COVID-19: second edition of a living systematic review with meta-analyses and trial sequential analyses (The LIVING Project)": S27 Table.docx

S27 Summary of Findings

| **High-flow nasal versus standard bag-valve oxygenation for fiberoptic intubation of patients with COVID-19** | | | | | | |
| --- | --- | --- | --- | --- | --- | --- |
| **Patients or population:** Anyone with a diagnosis of COVID-19  **Setting:** Any setting  **Intervention:** High-flow nasal oxygenation  **Comparison:** Standard bag-valve oxygenation | | | | | | |
| **Outcomes** | **Anticipated absolute effects* (95% CI)** | | **Relative effect (95% CI)** | **No of participants (studies)** | **Certainty of the evidence (GRADE)** | **Comments** |
|  | **Risk with**  **Standard bag-valve oxygenation** | **Risk with**  **High-flow nasal oxygenation** |  |  |  |  |
| **All-cause mortality** | - | - | - | - | - | Outcome not yet measured or reported |
| **Serious adverse events** | - | - | - | - | - | Outcome not yet measured or reported |
| **Admission to intensive care** | - | - | - | - | - | Outcome not yet measured or reported |
| **Mechanical ventilation** | - | - | - | - | - | Outcome not yet measured or reported |
| **Renal replacement therapy** | - | - | - | - | - | Outcome not yet measured or reported |
| **Quality of Life** | - | - | - | - | - | Outcome not yet measured or reported |
| **Non-serious adverse events** | - | - | - | - | - | Outcome not yet measured or reported |
| **RR:** Risk ratio; **CI:** Confidence interval; **GRADE:** GRADE Working Group grades of evidence | | | | | | |
| **GRADE Working Group grades of evidence**  **High certainty:** We are very confident that the true effect lies close to that of the estimate of the effect **Moderate certainty:** We are moderately confident in the effect estimate: The true effect is likely to be close to the estimate of the effect, but there is a possibility that it is substantially different **Low certainty:** Our confidence in the effect estimate is limited: The true effect may be substantially different from the estimate of the effect **Very low certainty:** We have very little confidence in the effect estimate: The true effect is likely to be substantially different from the estimate of effect | | | | | | |
