## Supplementary figures and images for "Interventions for treatment of COVID-19: second edition of a living systematic review with meta-analyses and trial sequential analyses (The LIVING Project)"

### S1 Fig.tiff

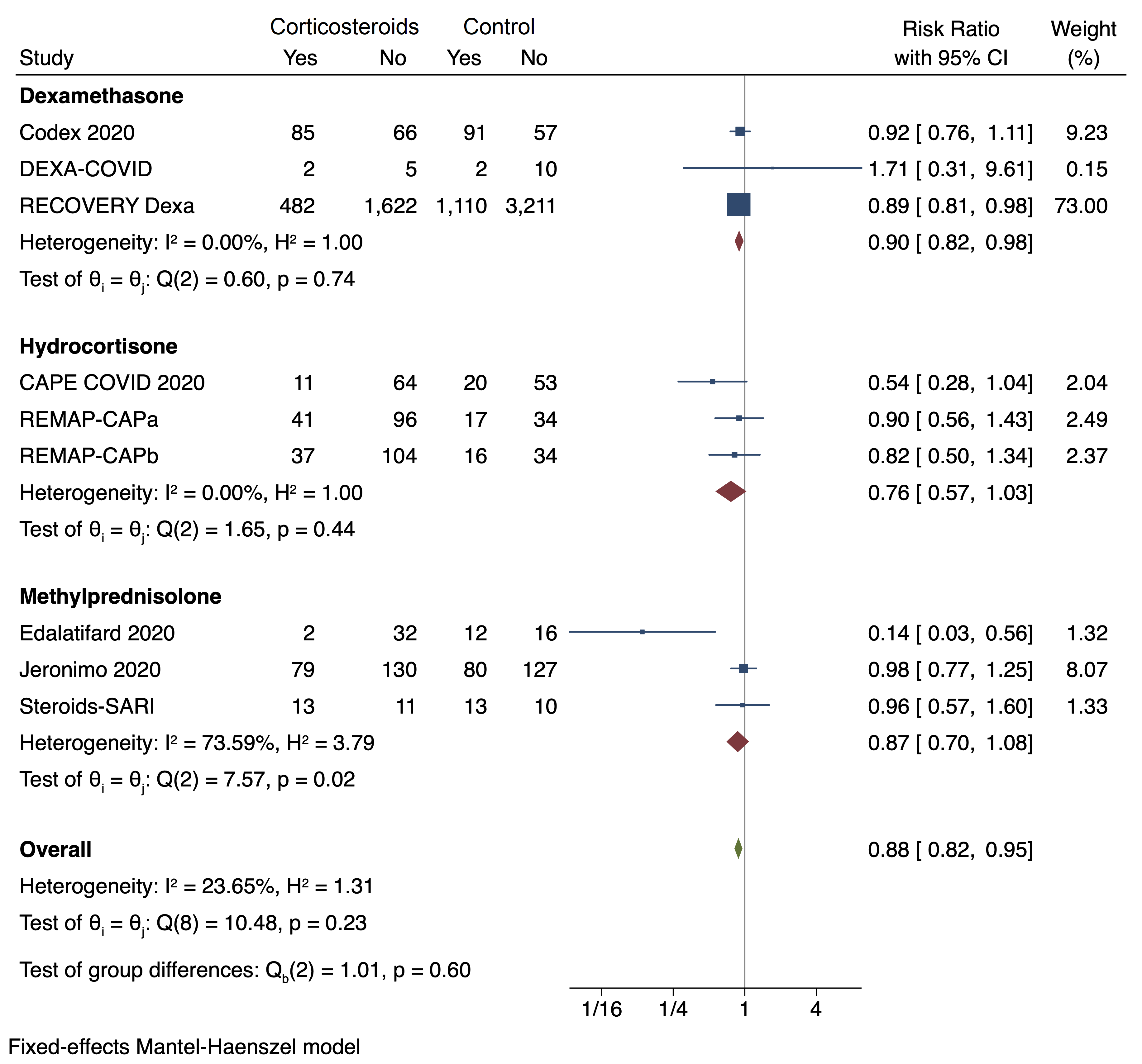

### S2 Fig.tiff

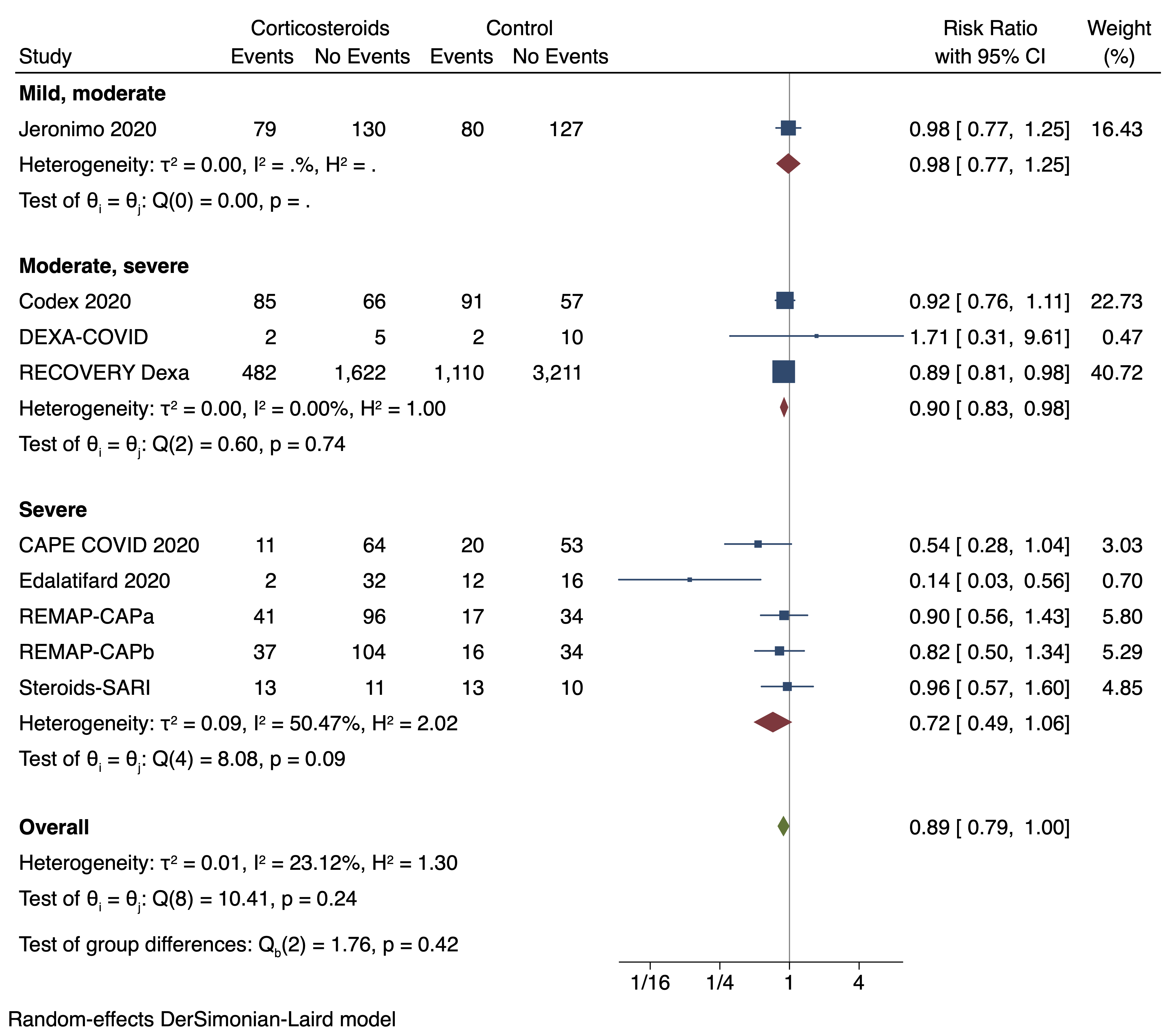

### S3 Fig.tiff

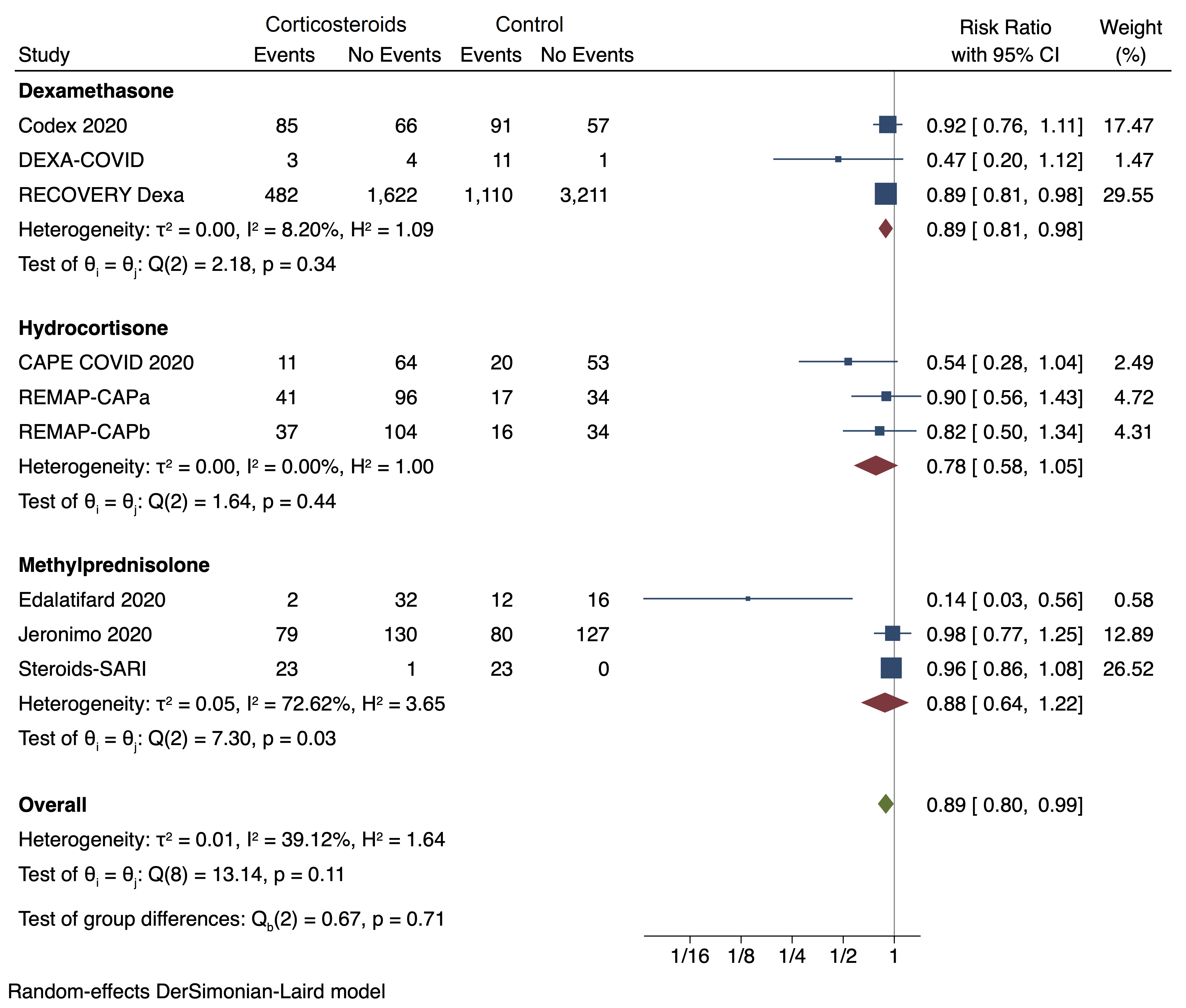

### S3 Table.tiff

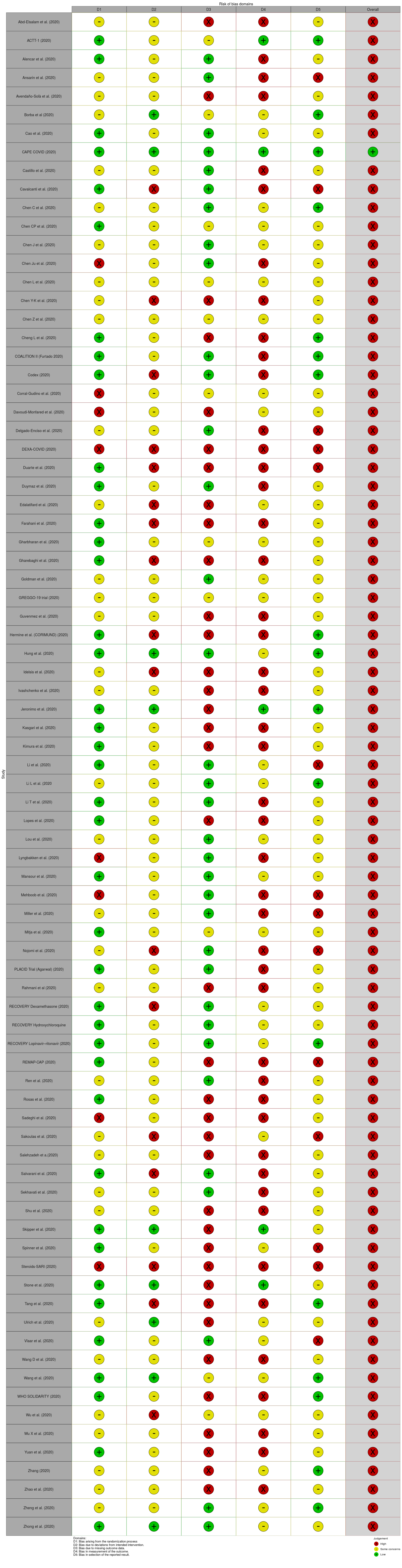

### S4 Fig.tiff

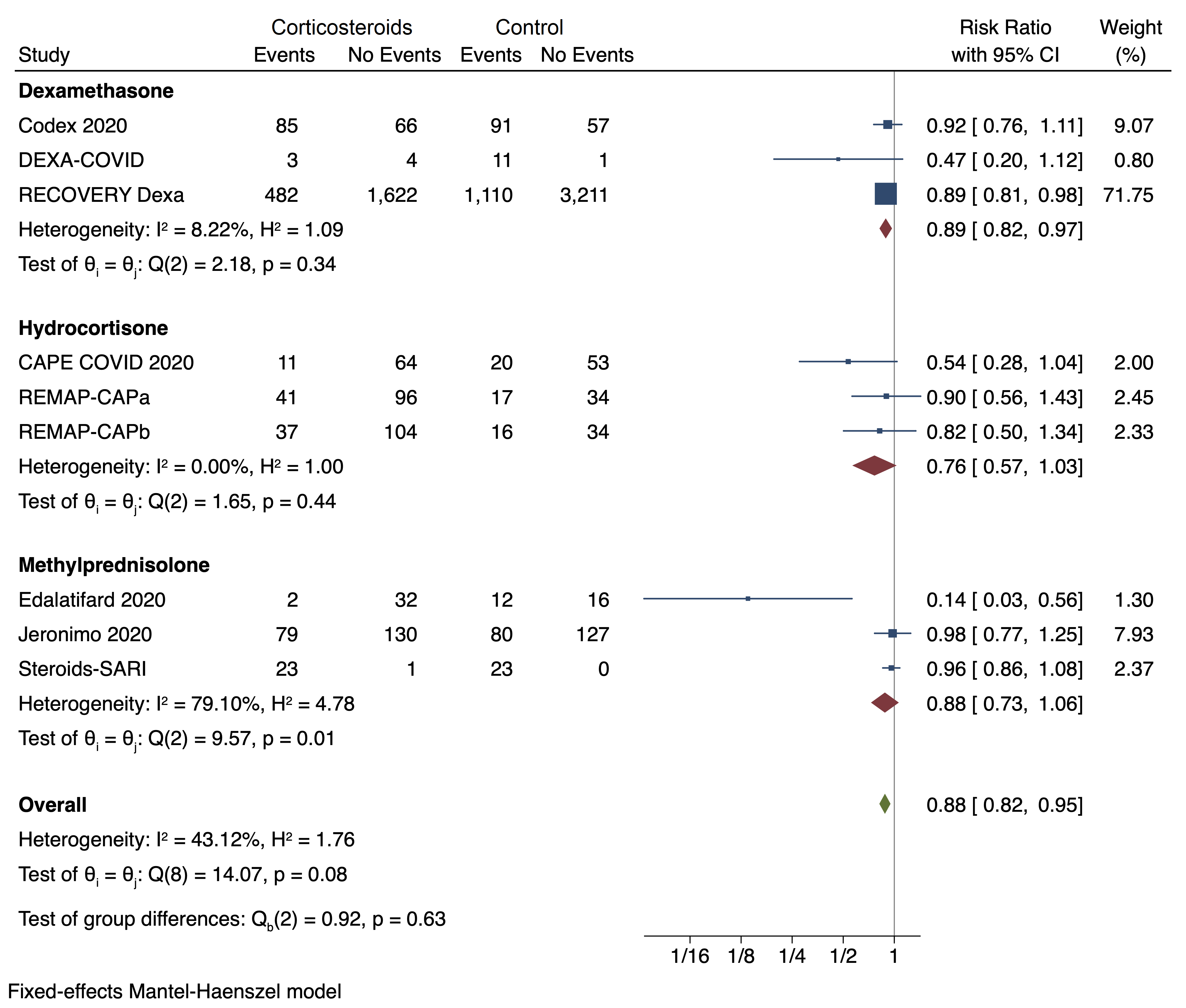

### S5 Fig.tiff

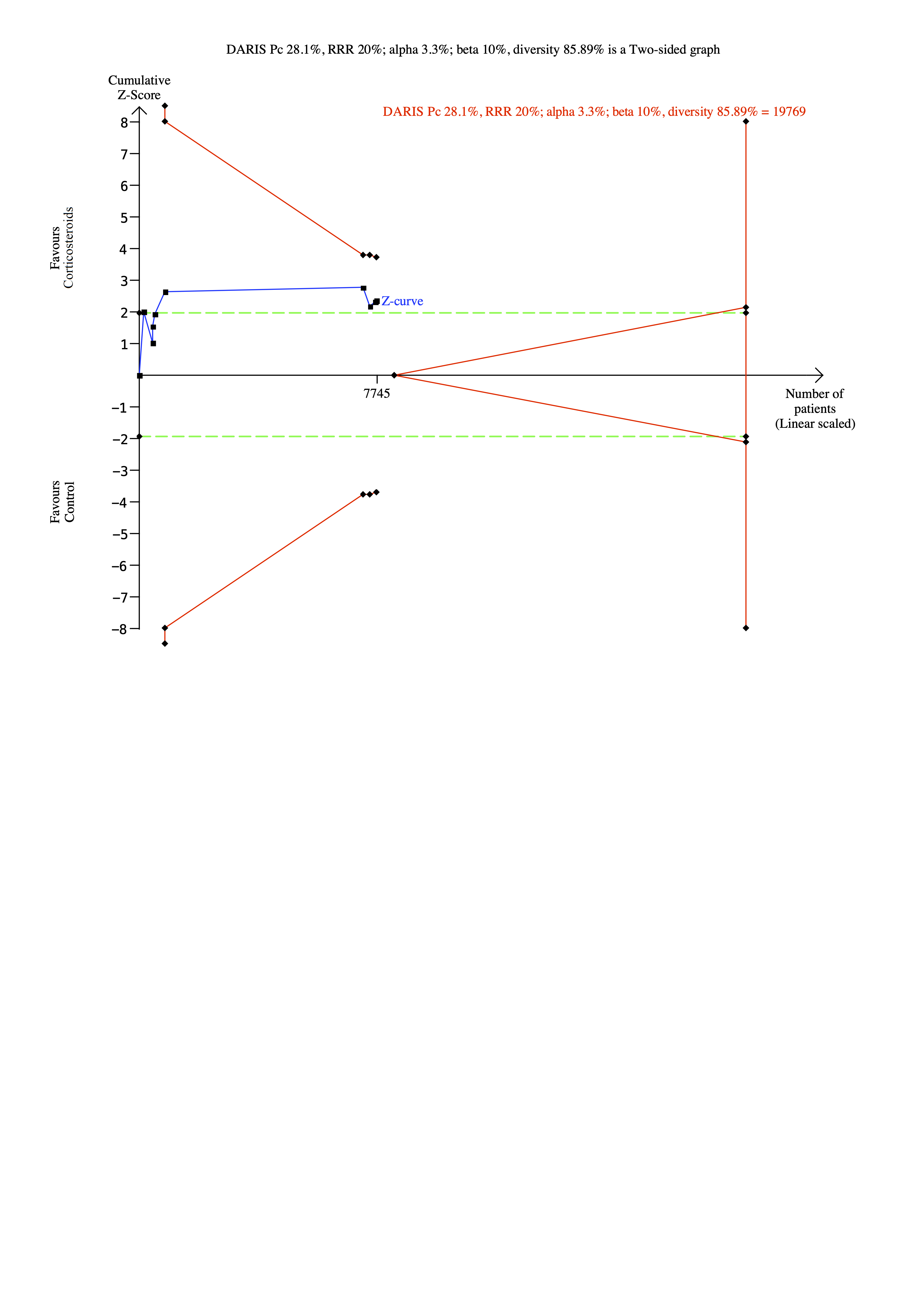

### S6 Fig.tiff

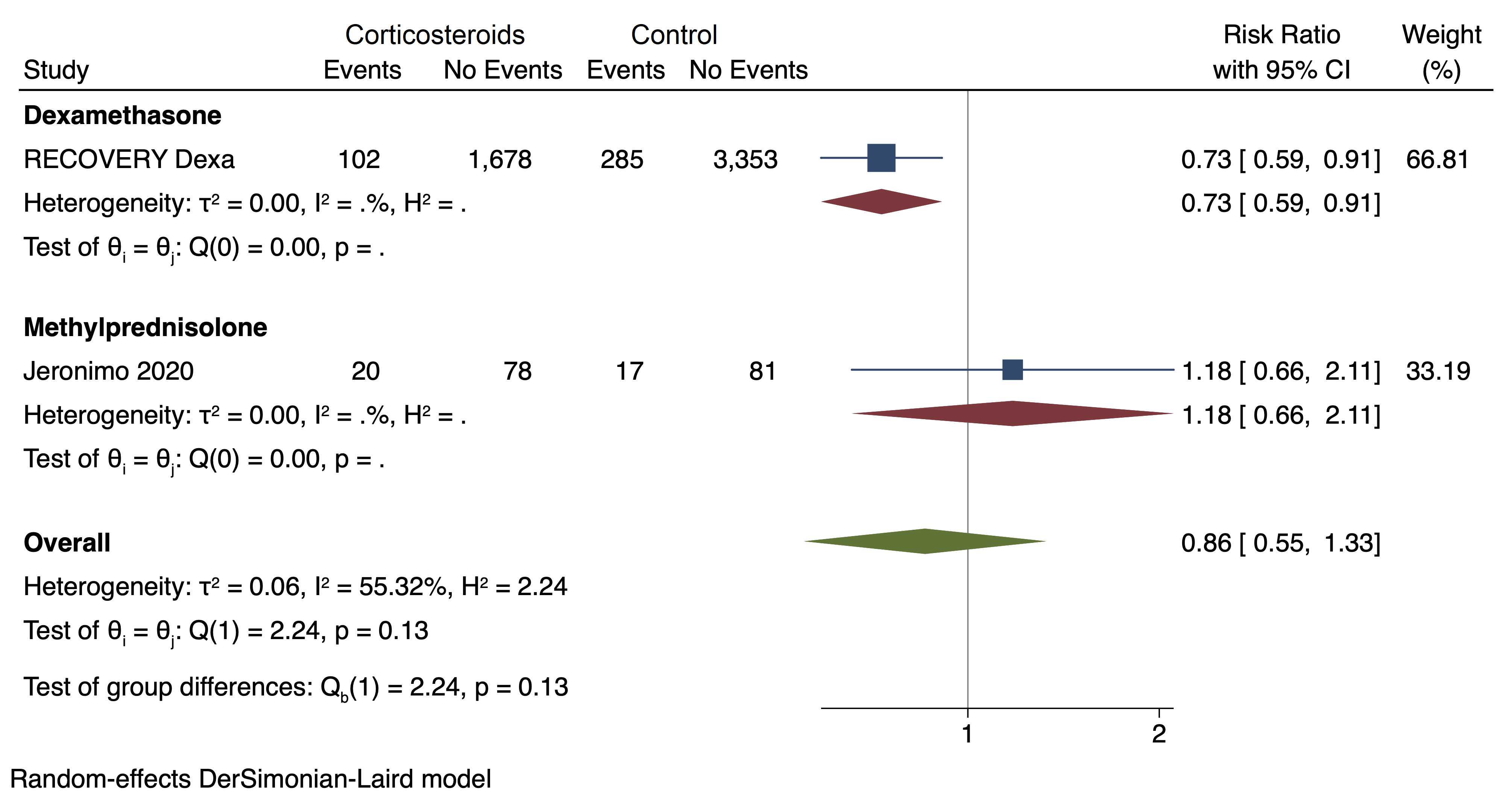

### S7 Fig.tiff

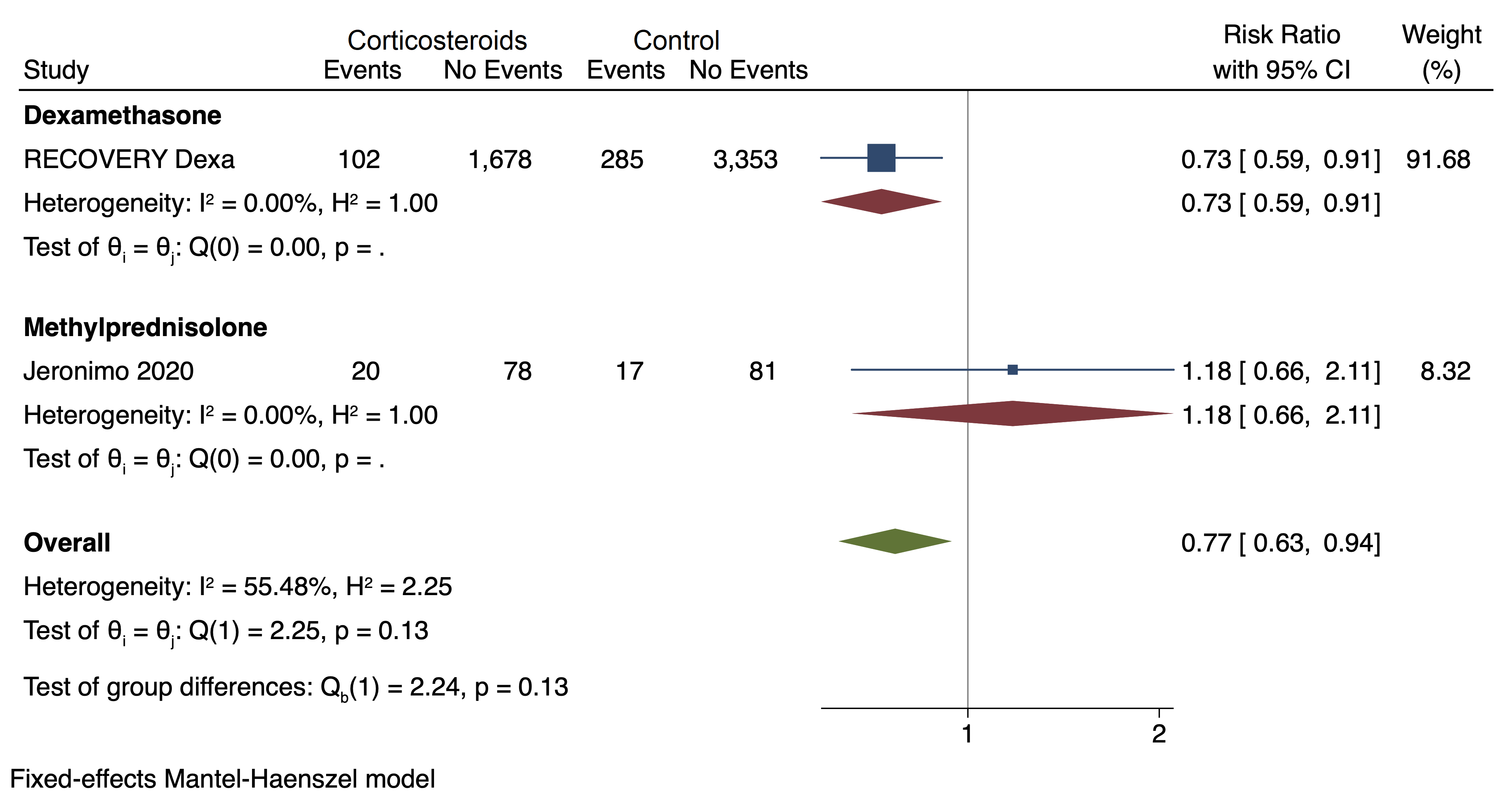

### S8 Fig.tiff

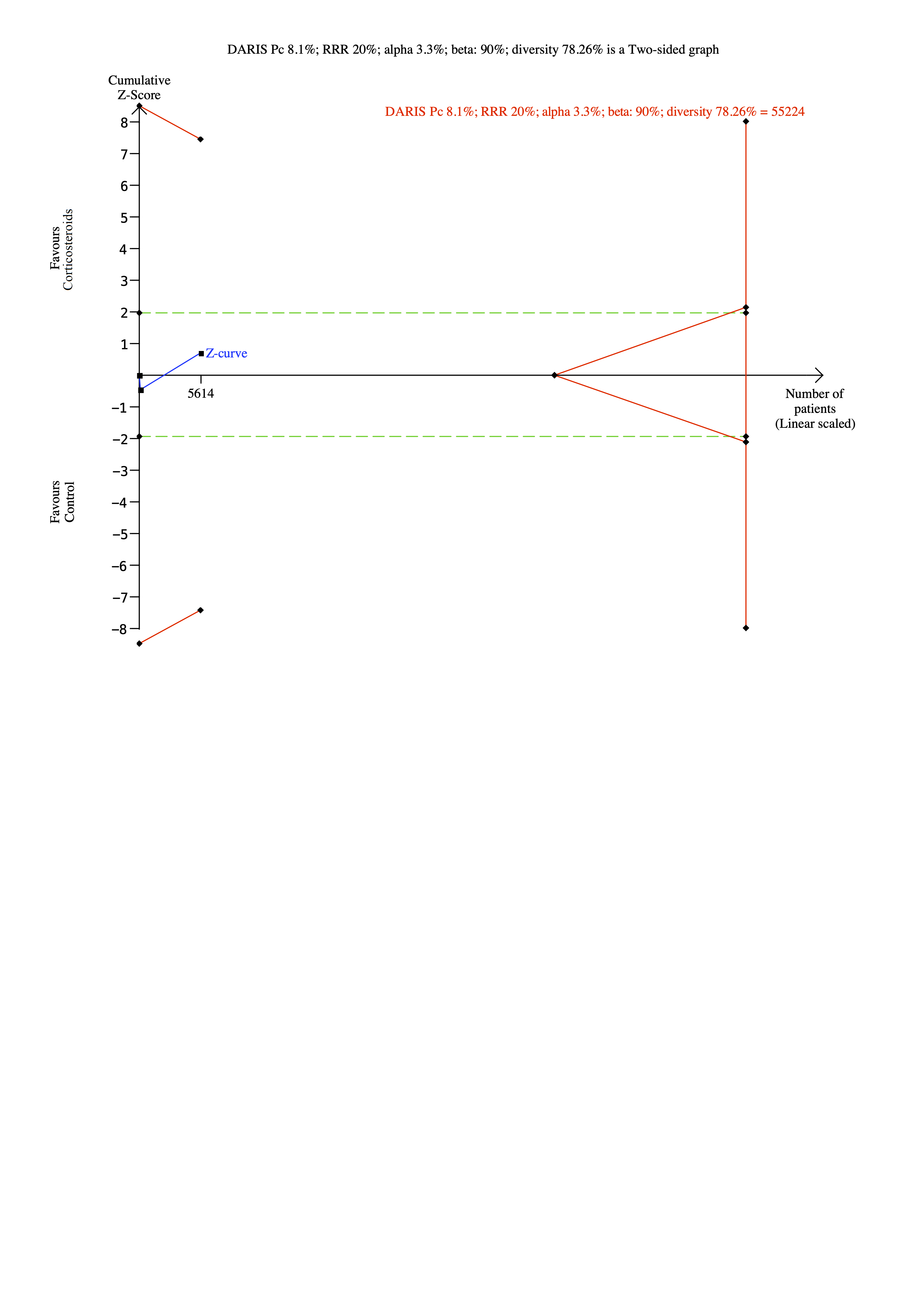

### S9 Fig.tiff

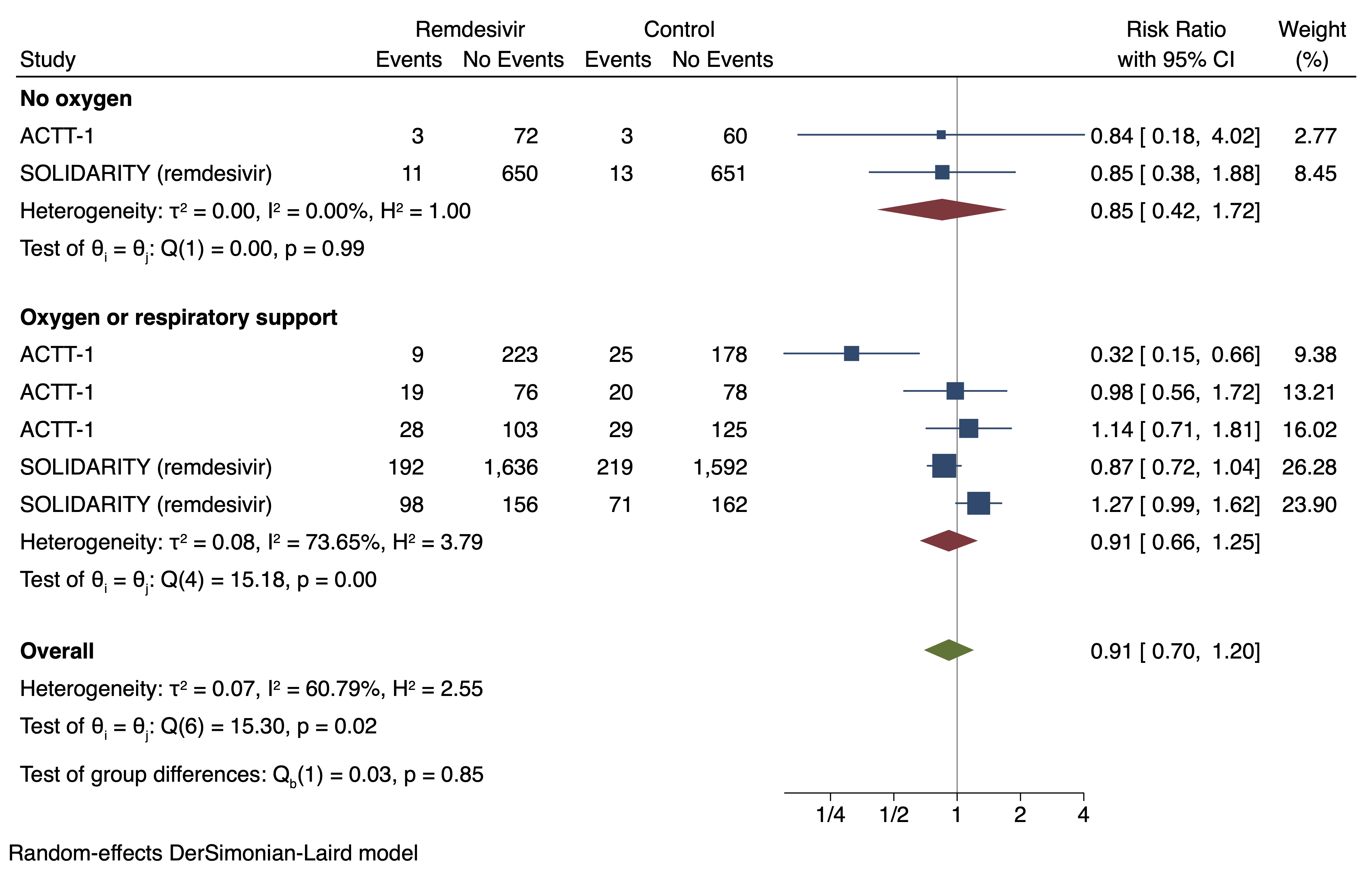

### S10 Fig.tiff

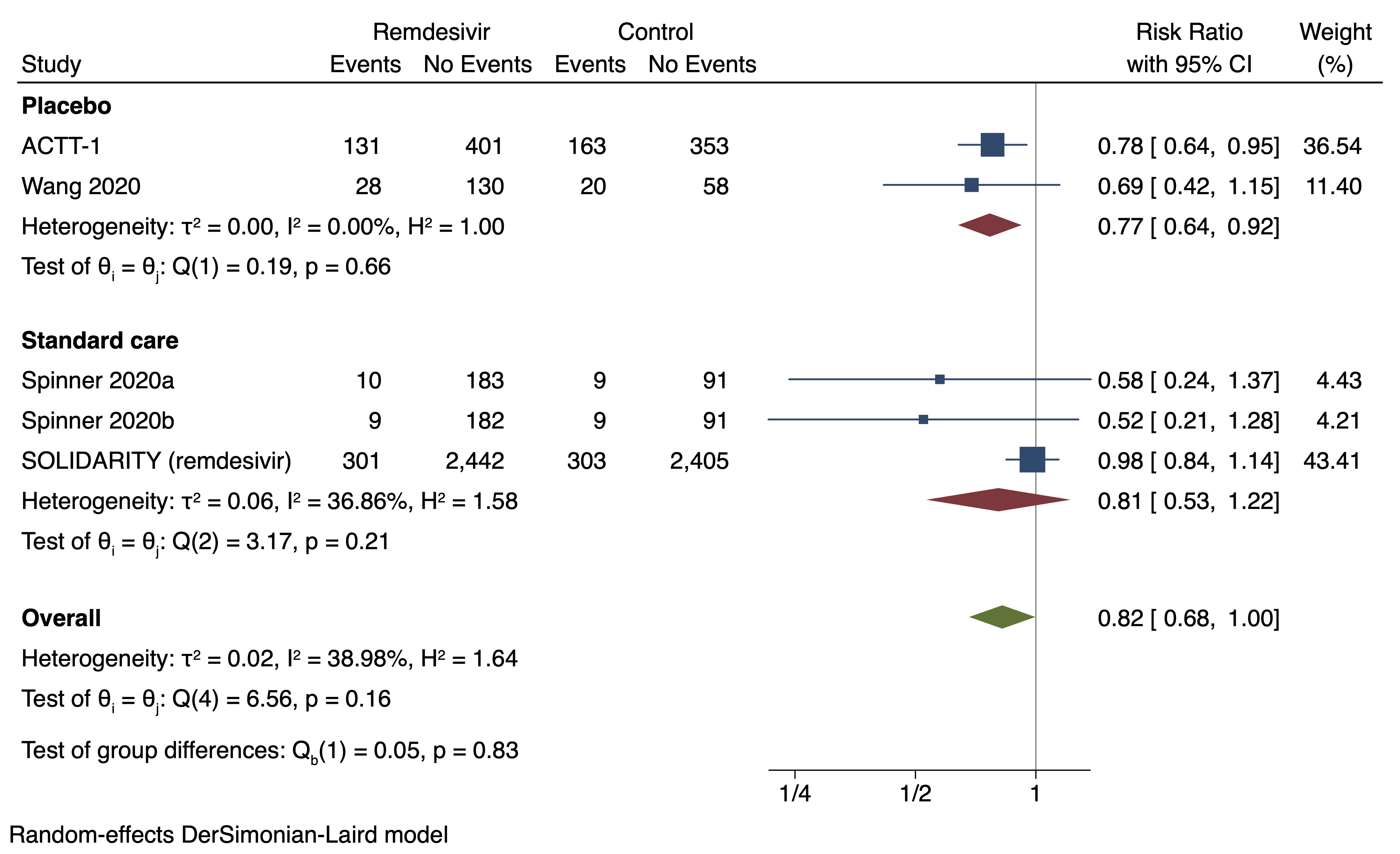

### S11 Fig.tiff

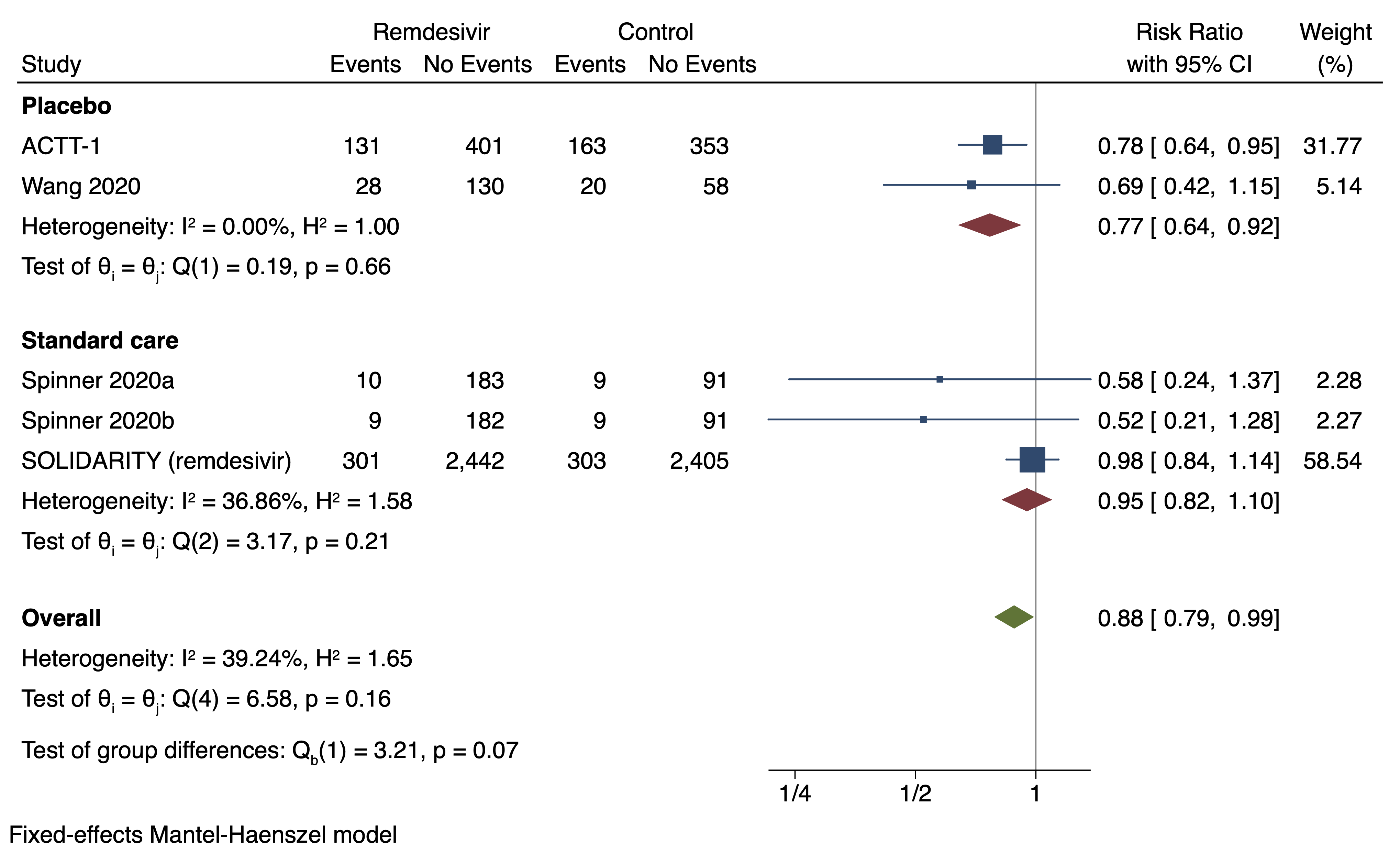

### S12 Fig.tiff

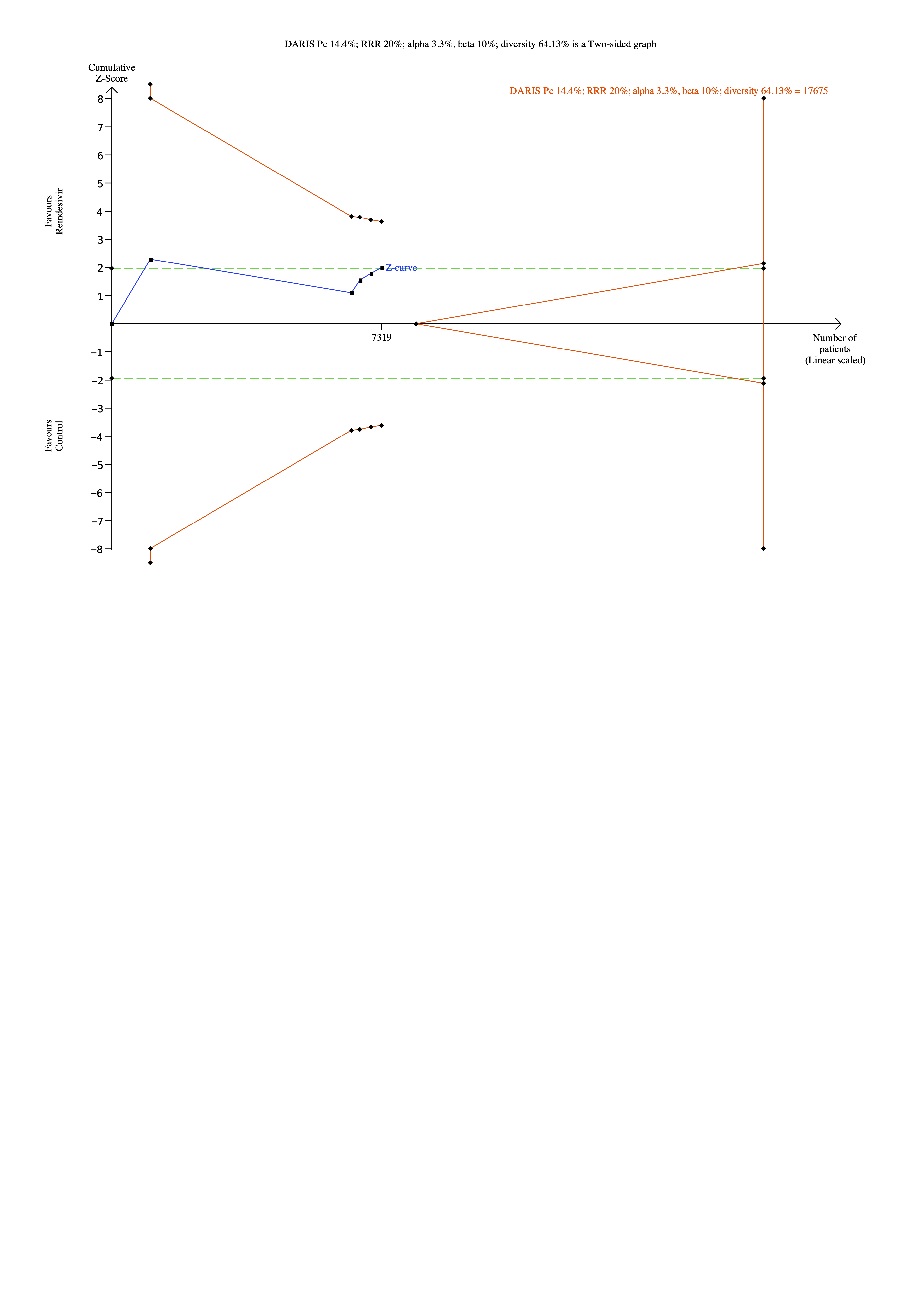

### S13 Fig .tiff

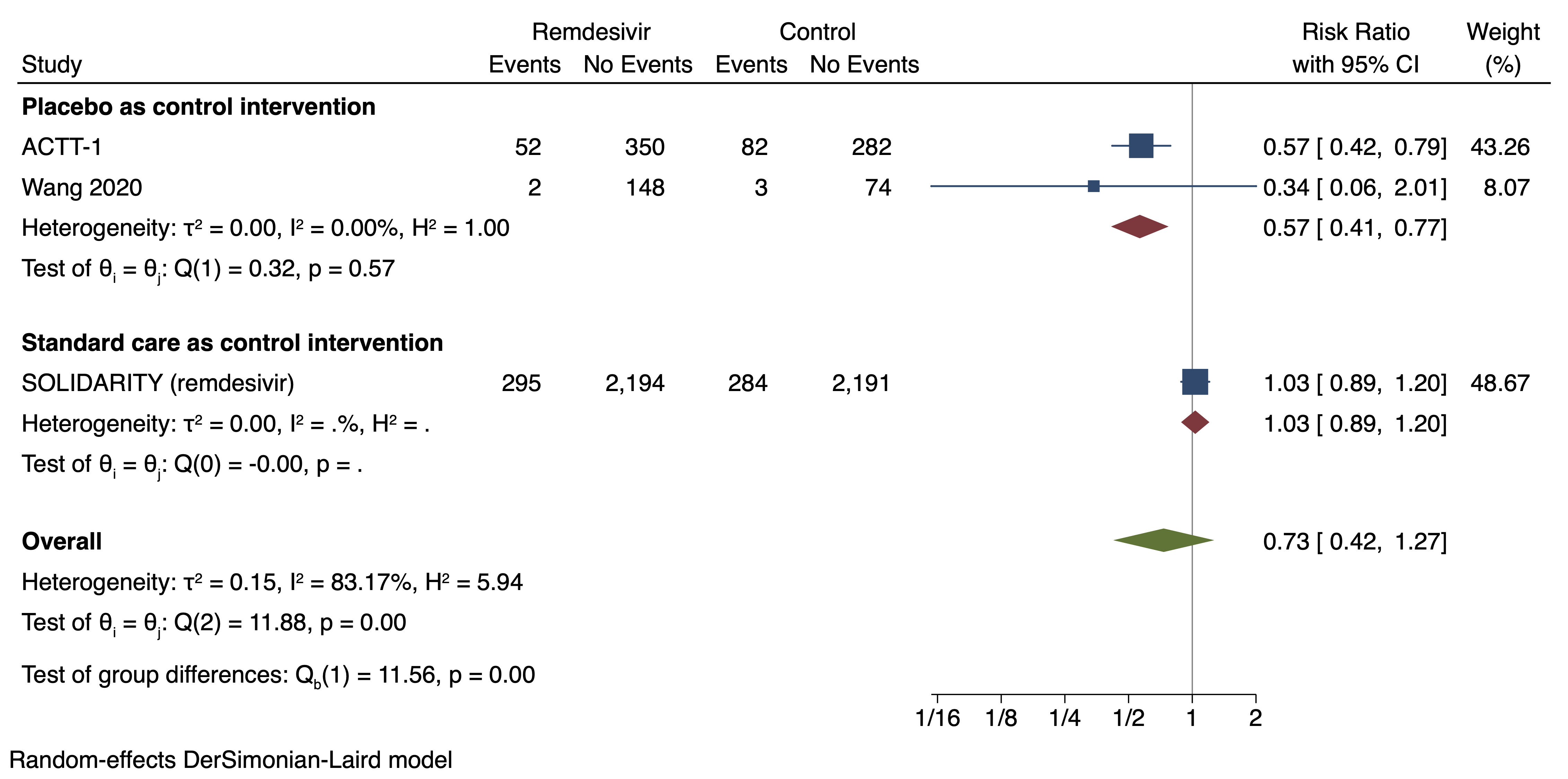

### S14 Fig.tiff

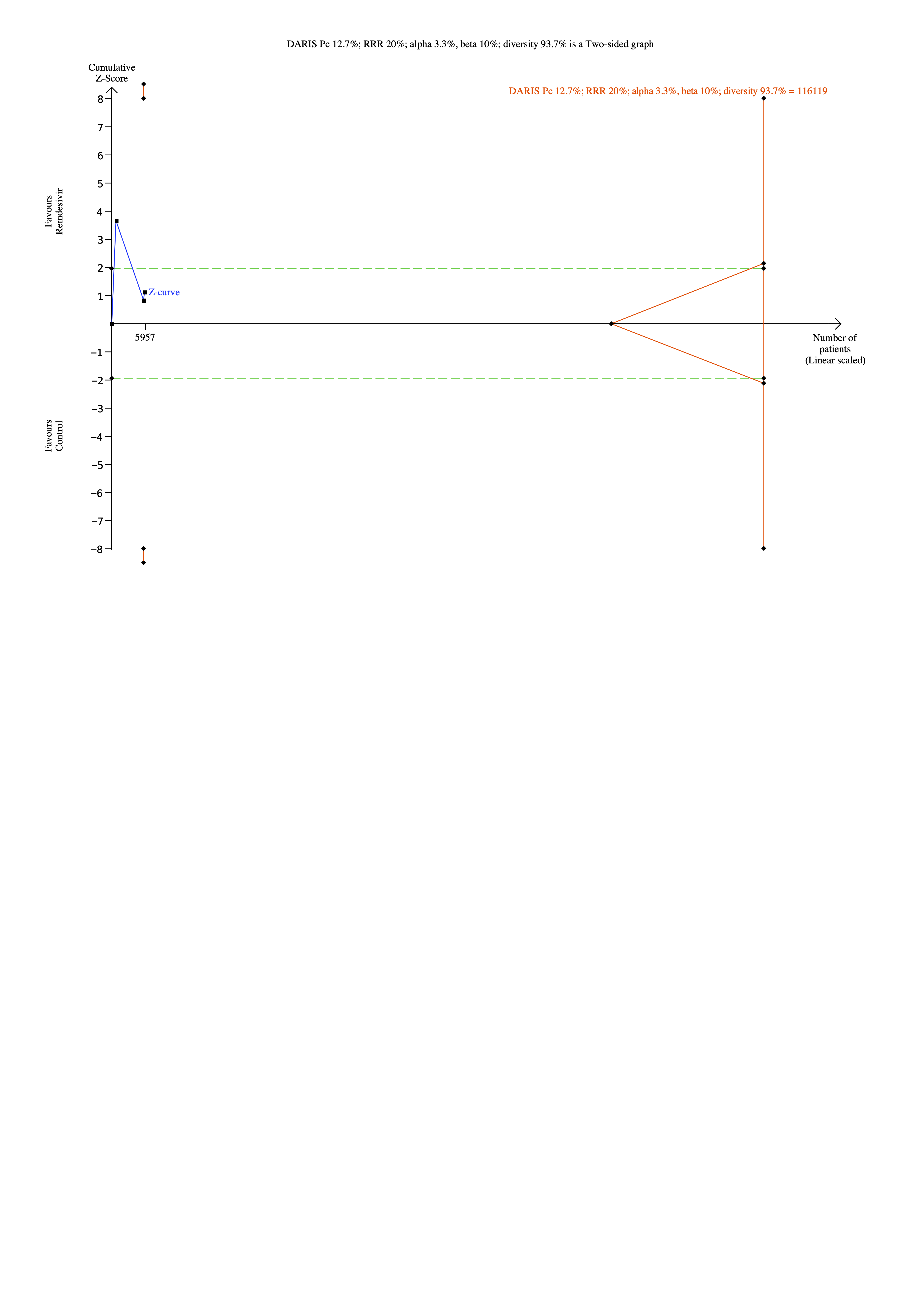

### S15 Fig.tiff

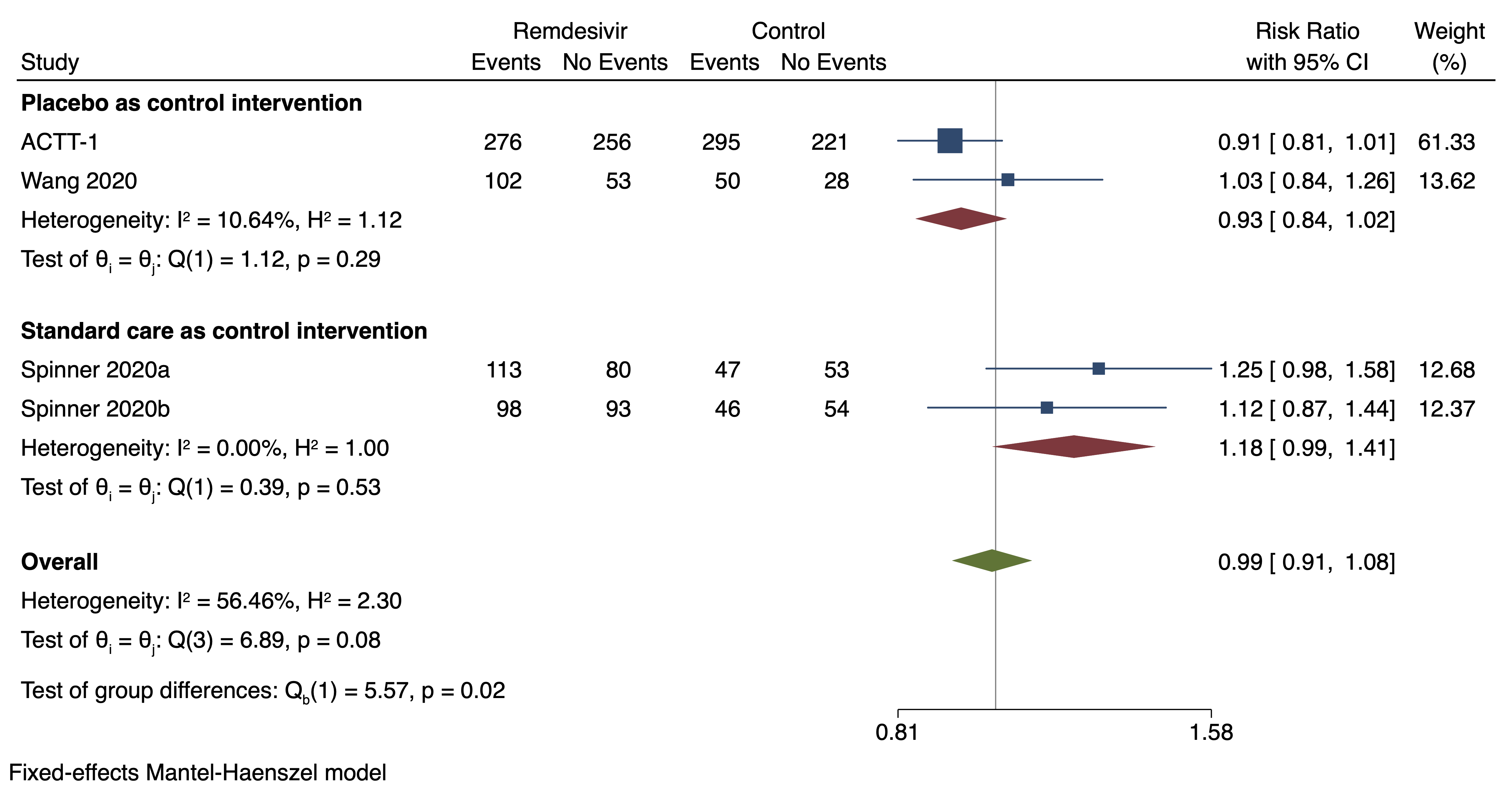

### S16 Fig.tiff

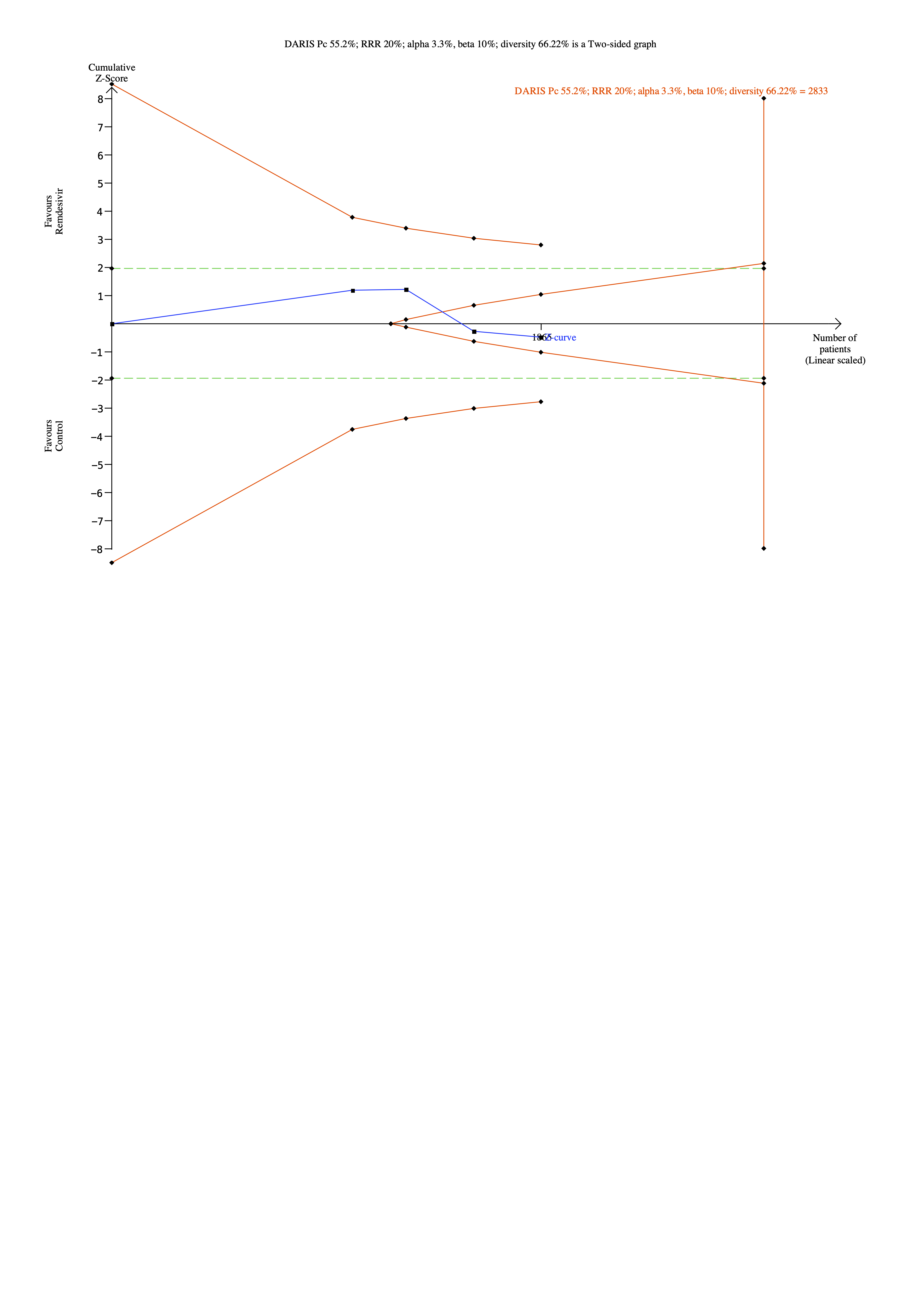

### S18 Fig.pdf

DARIS Pc 19.1%; RRR 20%; alpha 3.3%; beta 10%; diversity 0.00% is a Two-sided graph

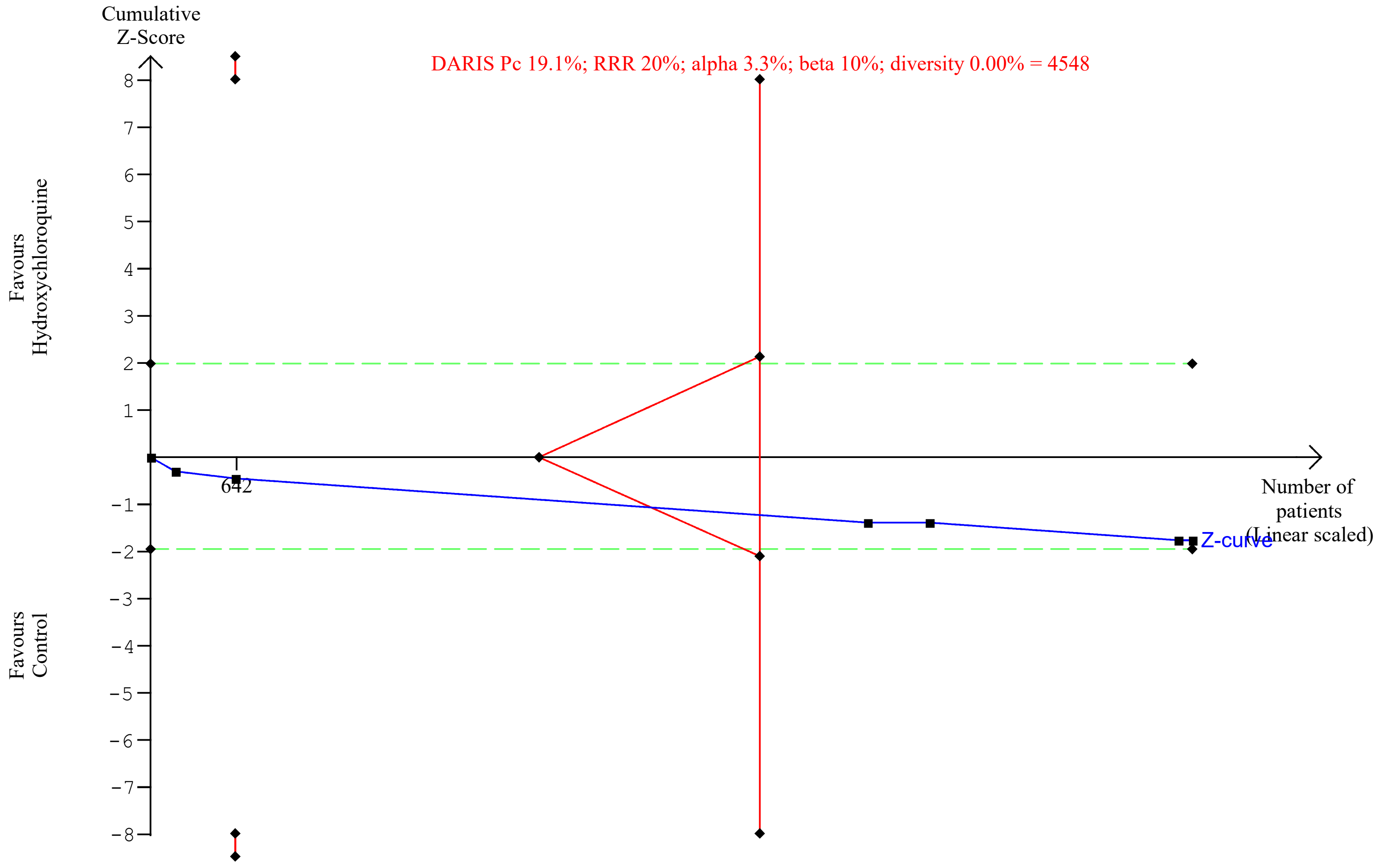

### S20 Fig.pdf

DARIS Pc 18.6%; RRR 20%; alpha 3.3%; beta 10%; diversity 0.00% is a Two-sided graph

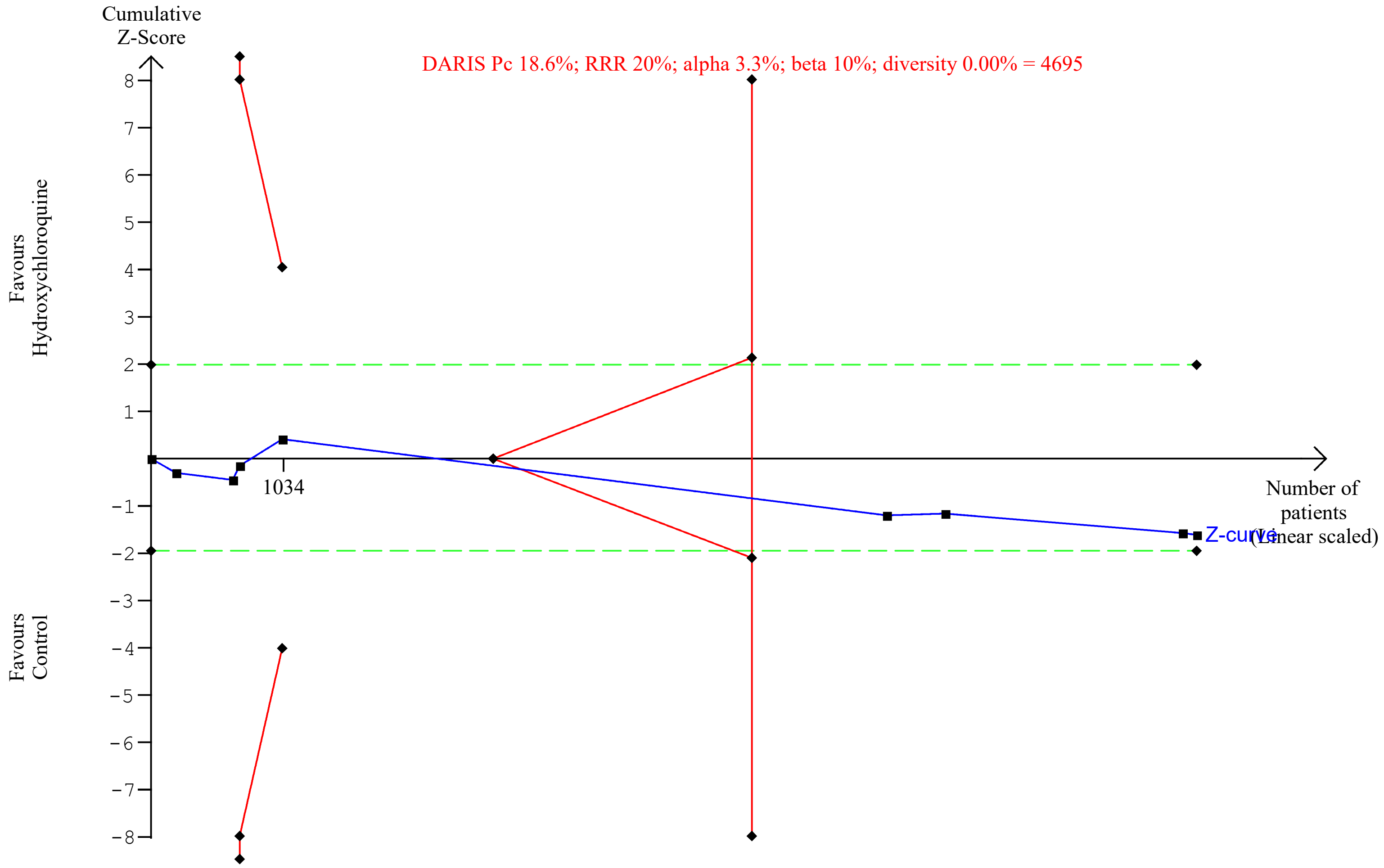

### S22 Fig.pdf

DARIS Pc 18.4%; RRR 20%; alpha 3.3%; beta 10%; diversity 0.00% is a Two-sided graph

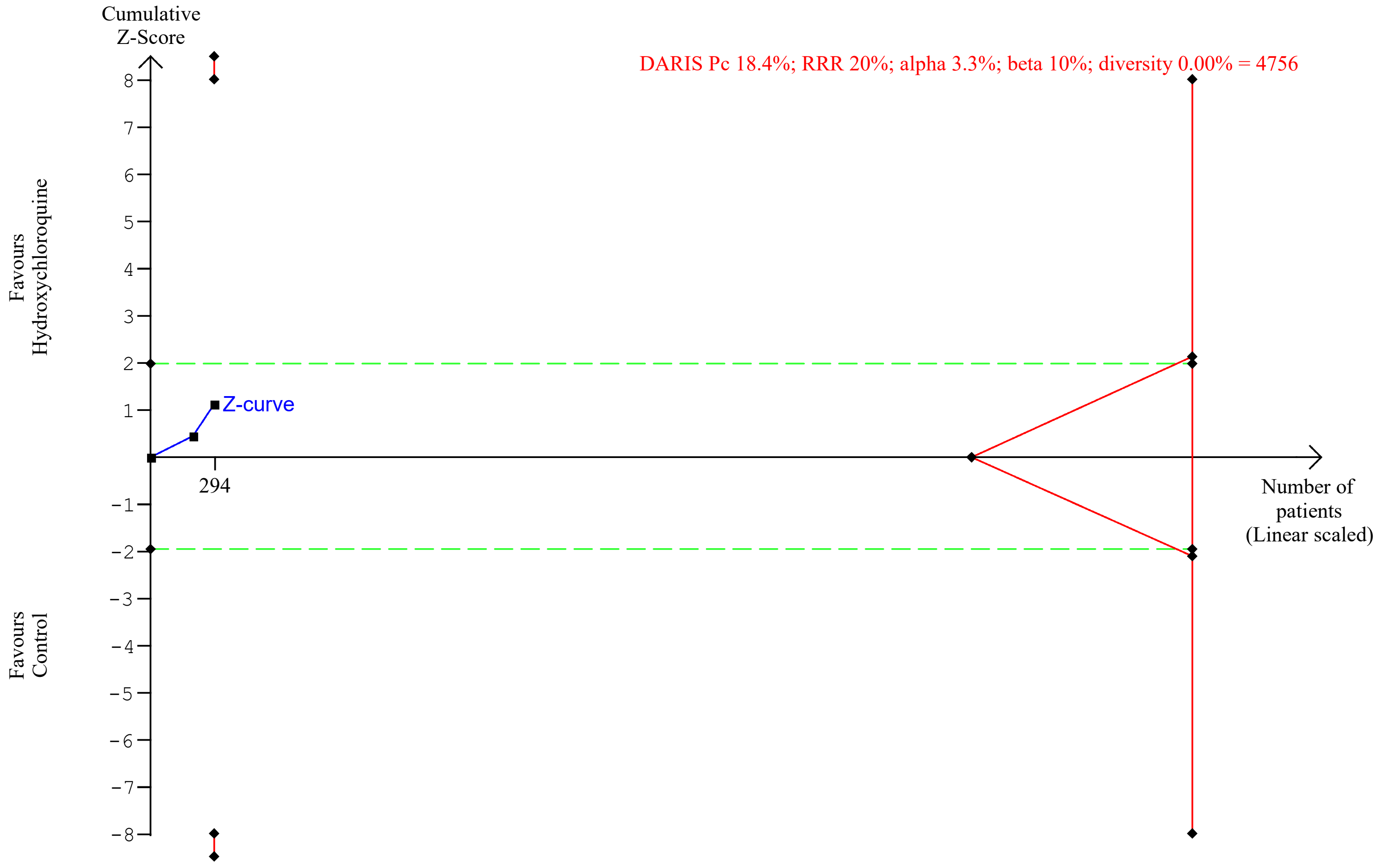

### S24 Fig.pdf

DARIS Pc 7.3%; RRR 20%; alpha 3.3%; beta 10%; diversity 0.00% is a Two-sided graph

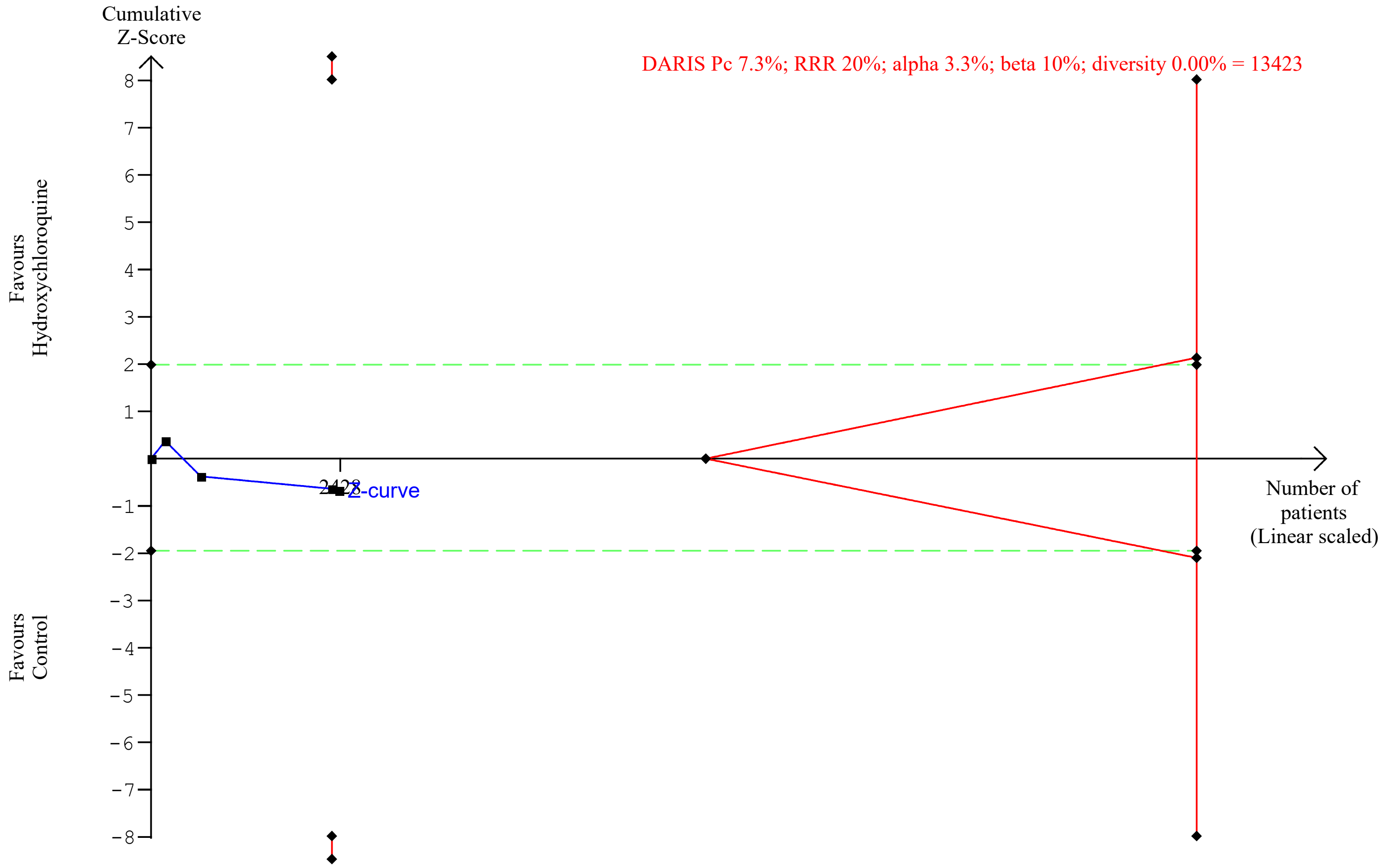

### S26 Fig.tiff

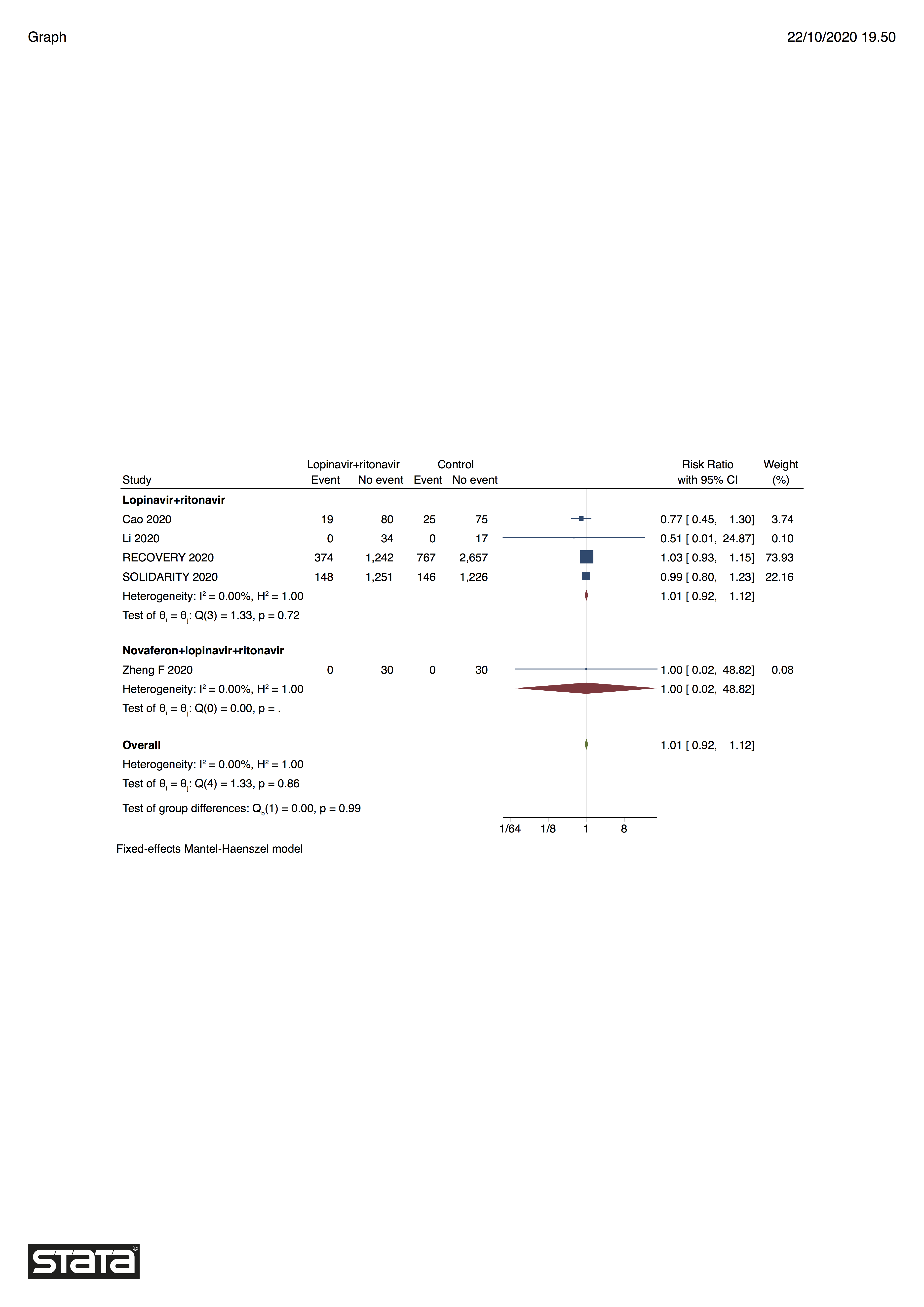

### S27 Fig.tiff

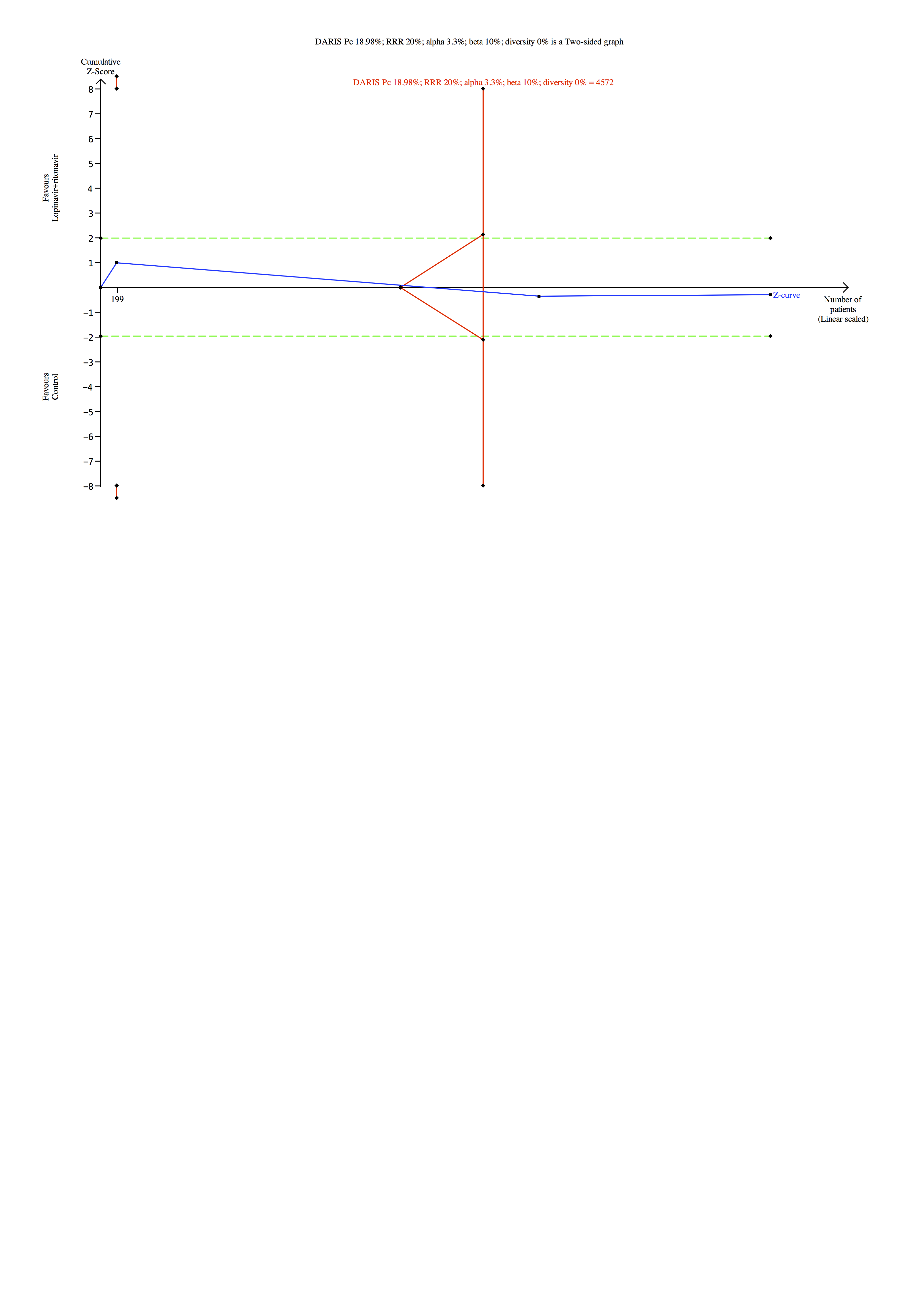

### S28 Fig.tiff

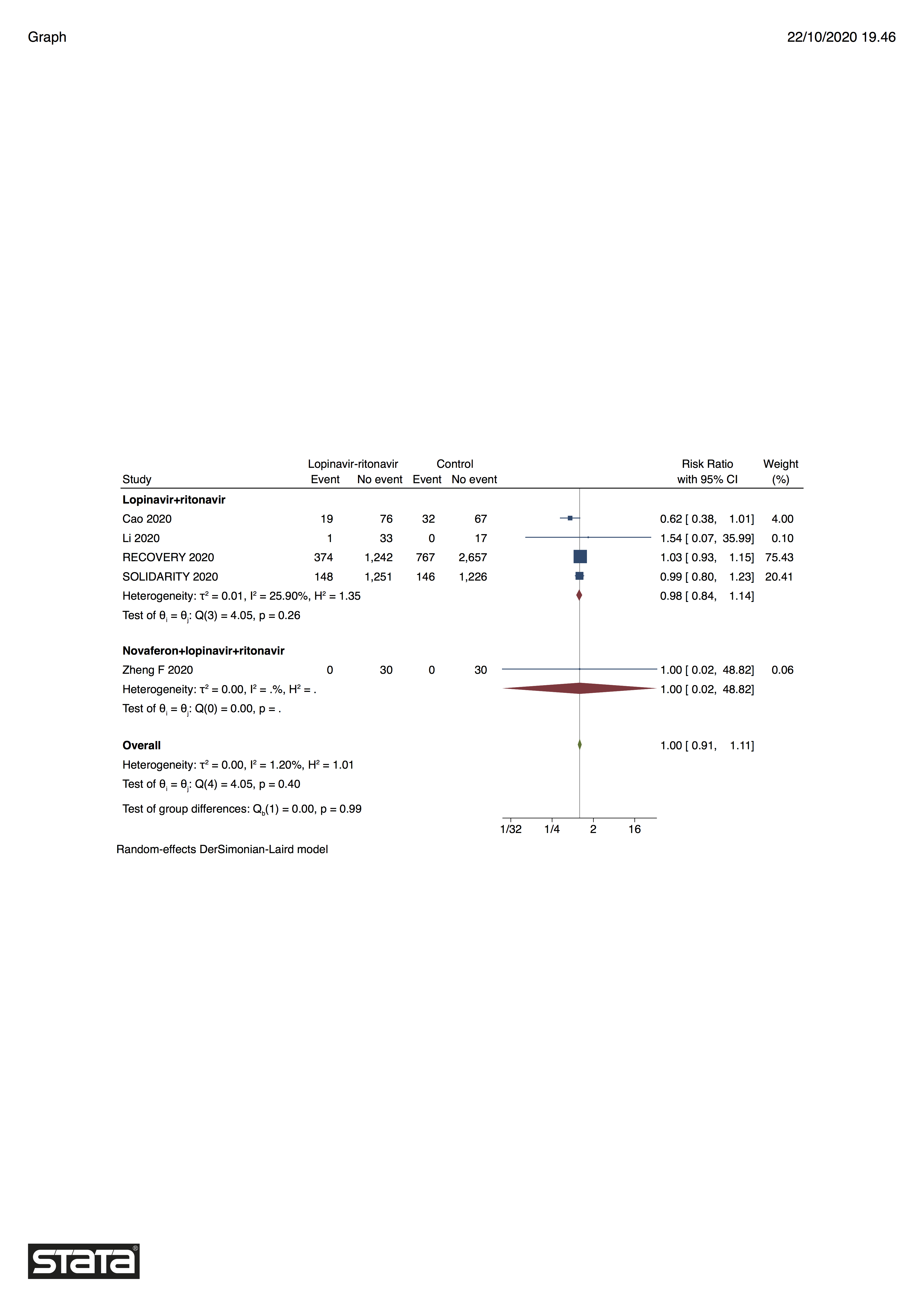

### S29 Fig.tiff

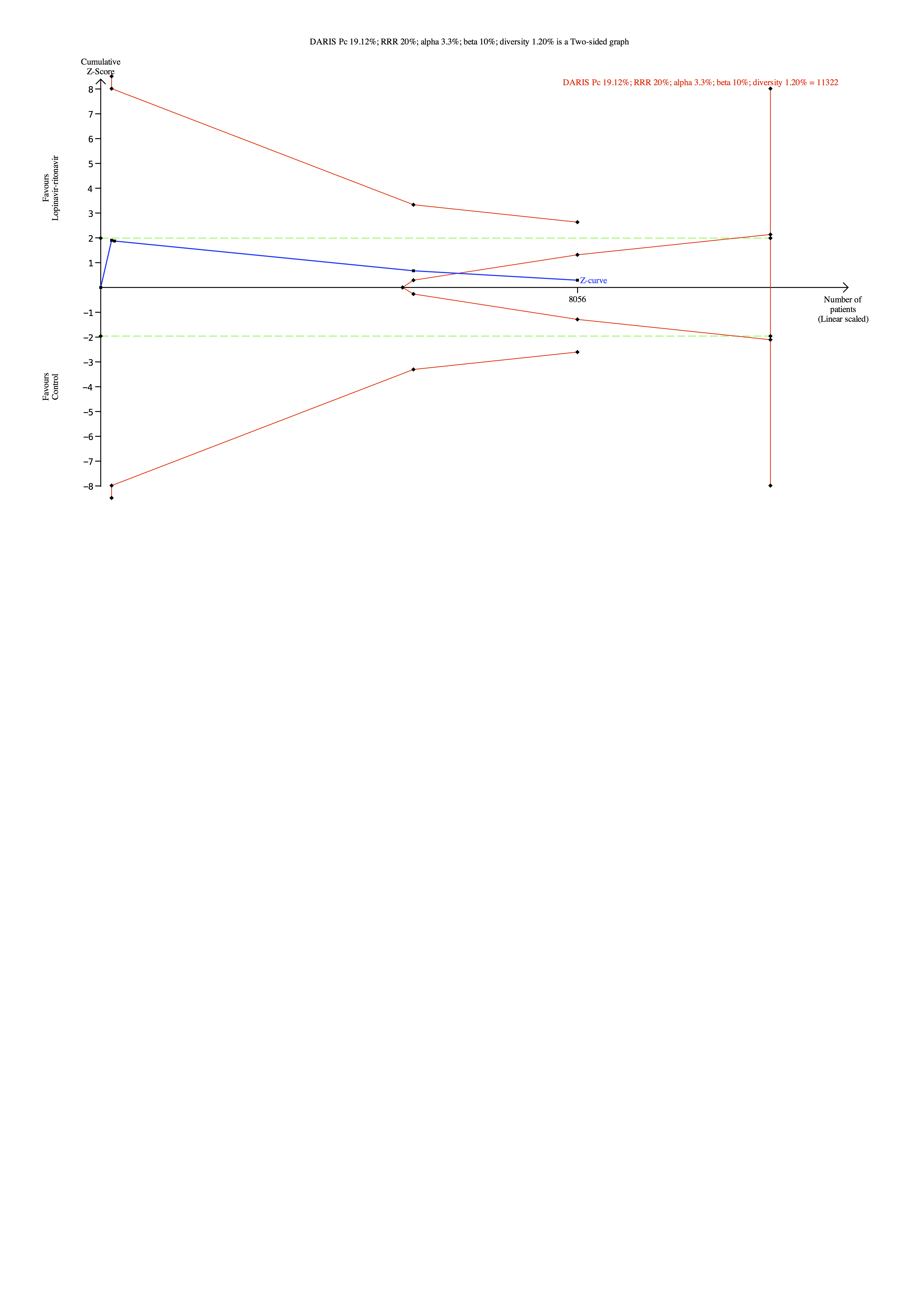

### S30 Fig.tiff

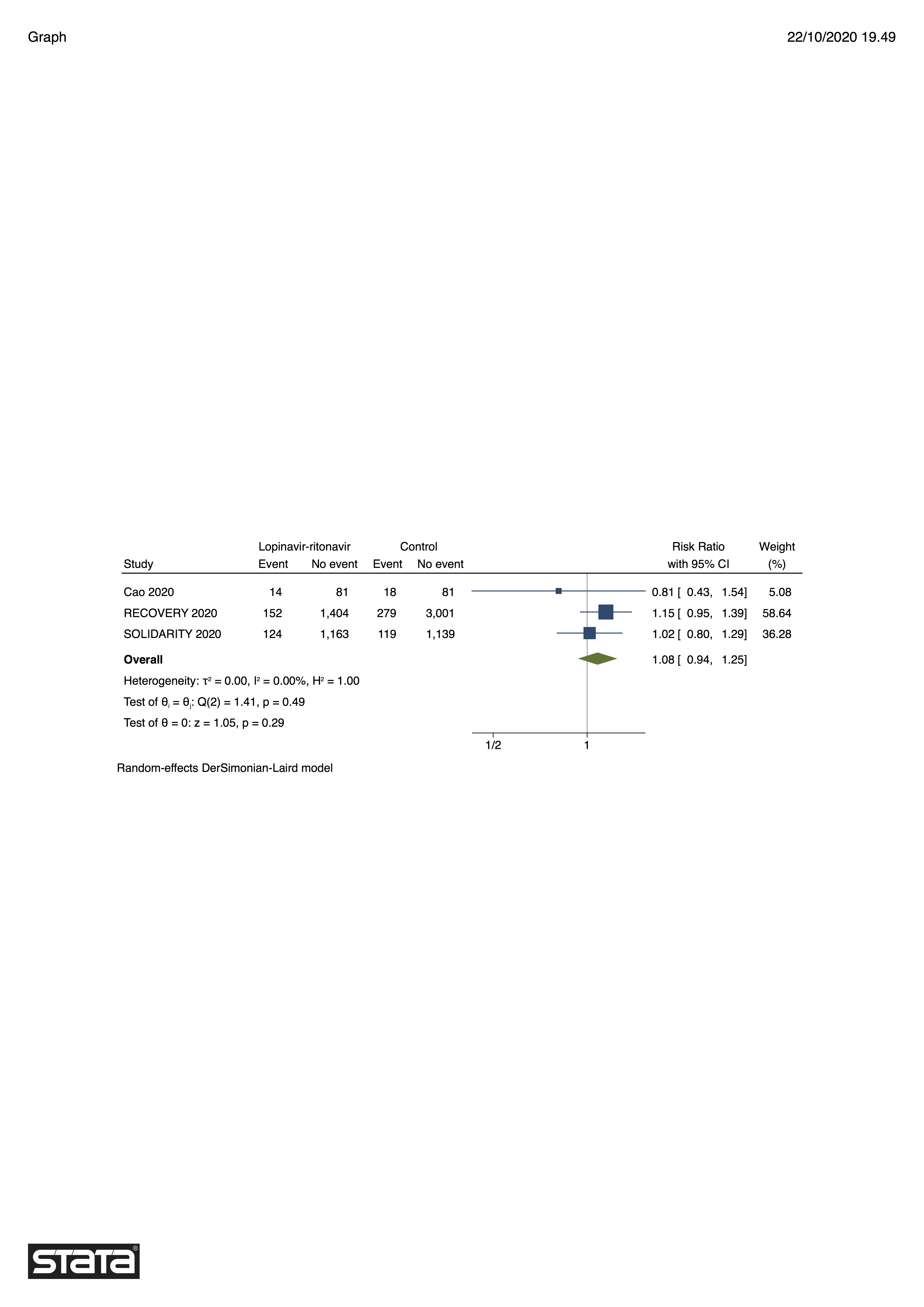

### S31 Fig.tiff

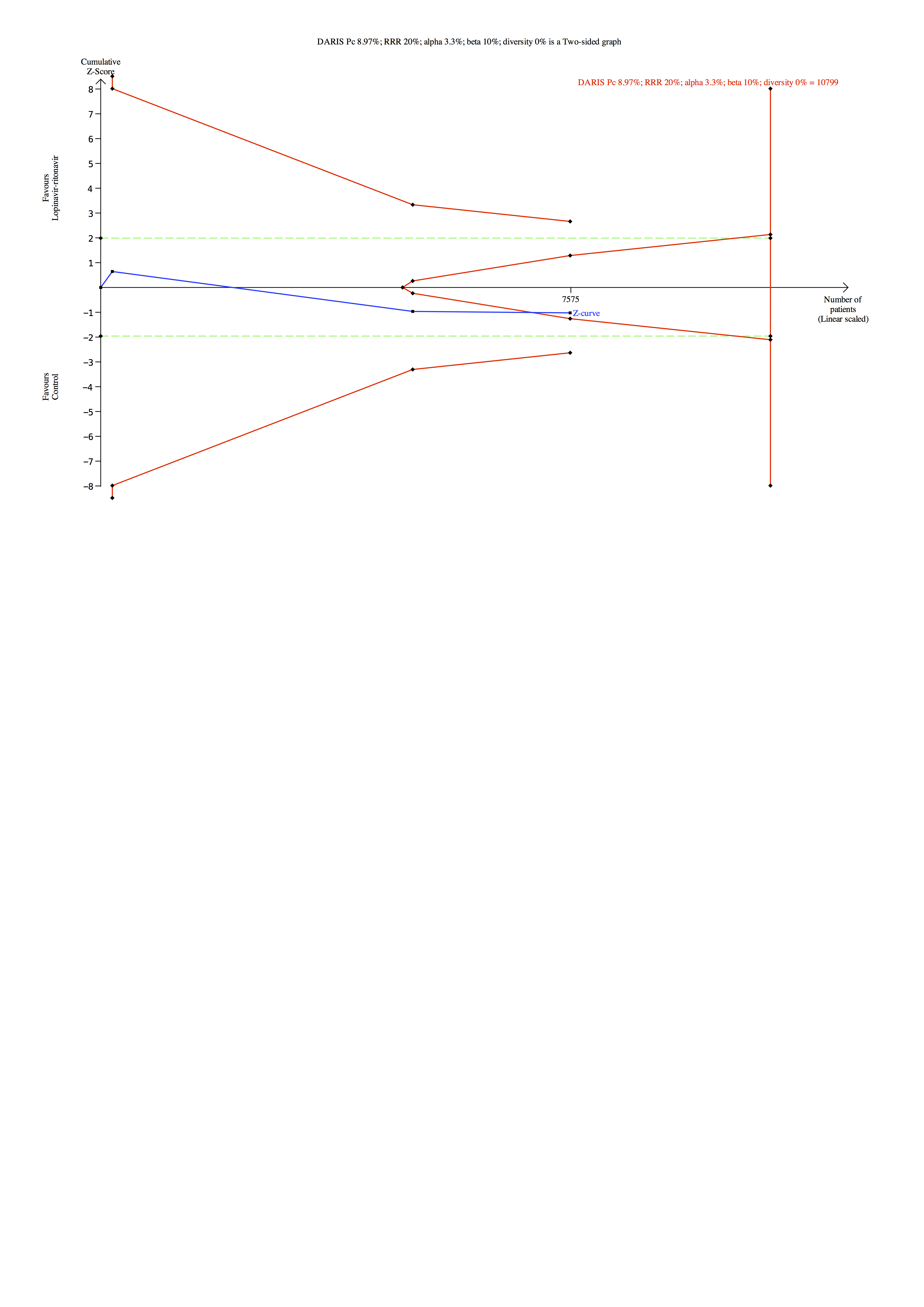

### S32 Fig.tiff

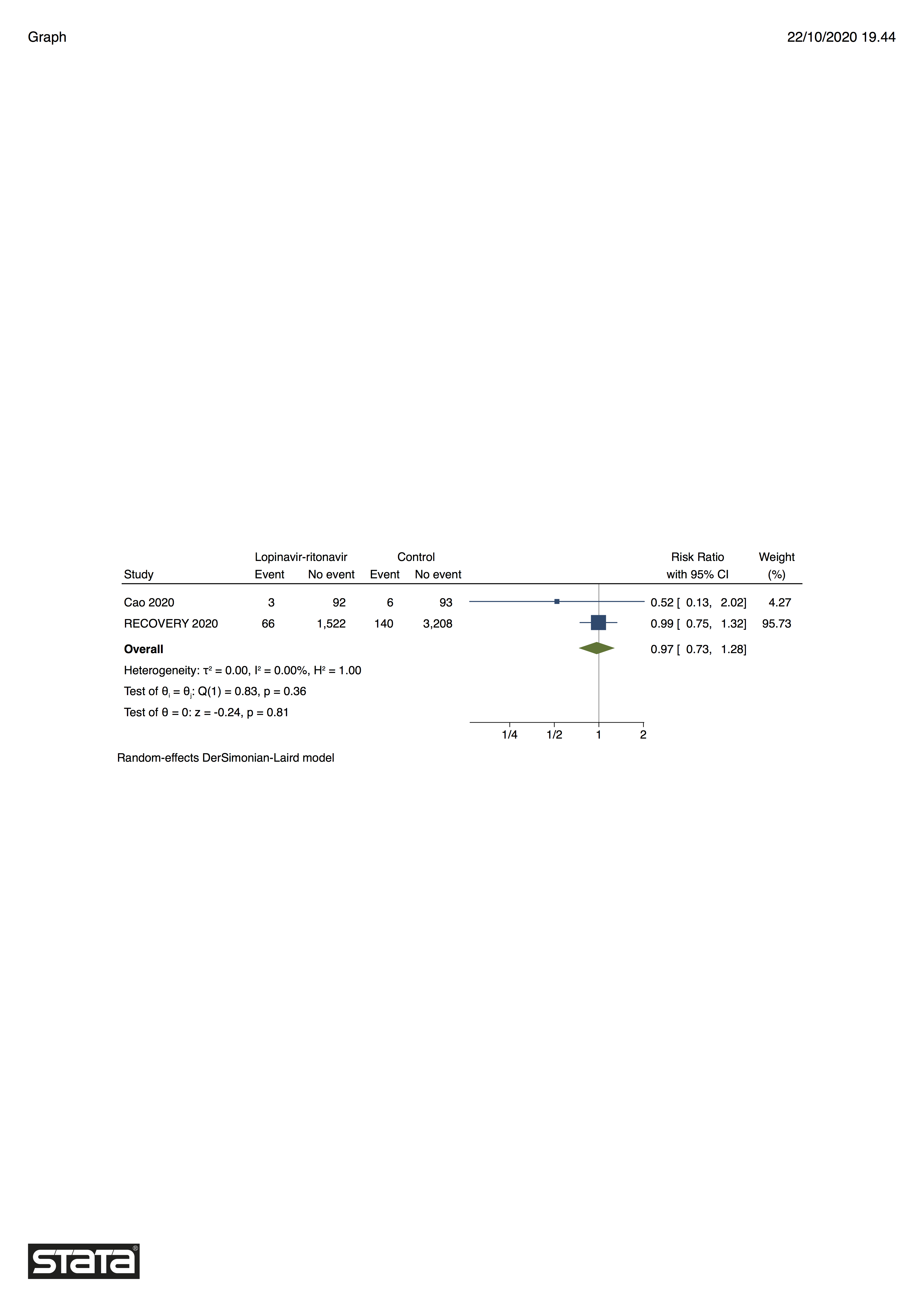

### S33 Fig.tiff

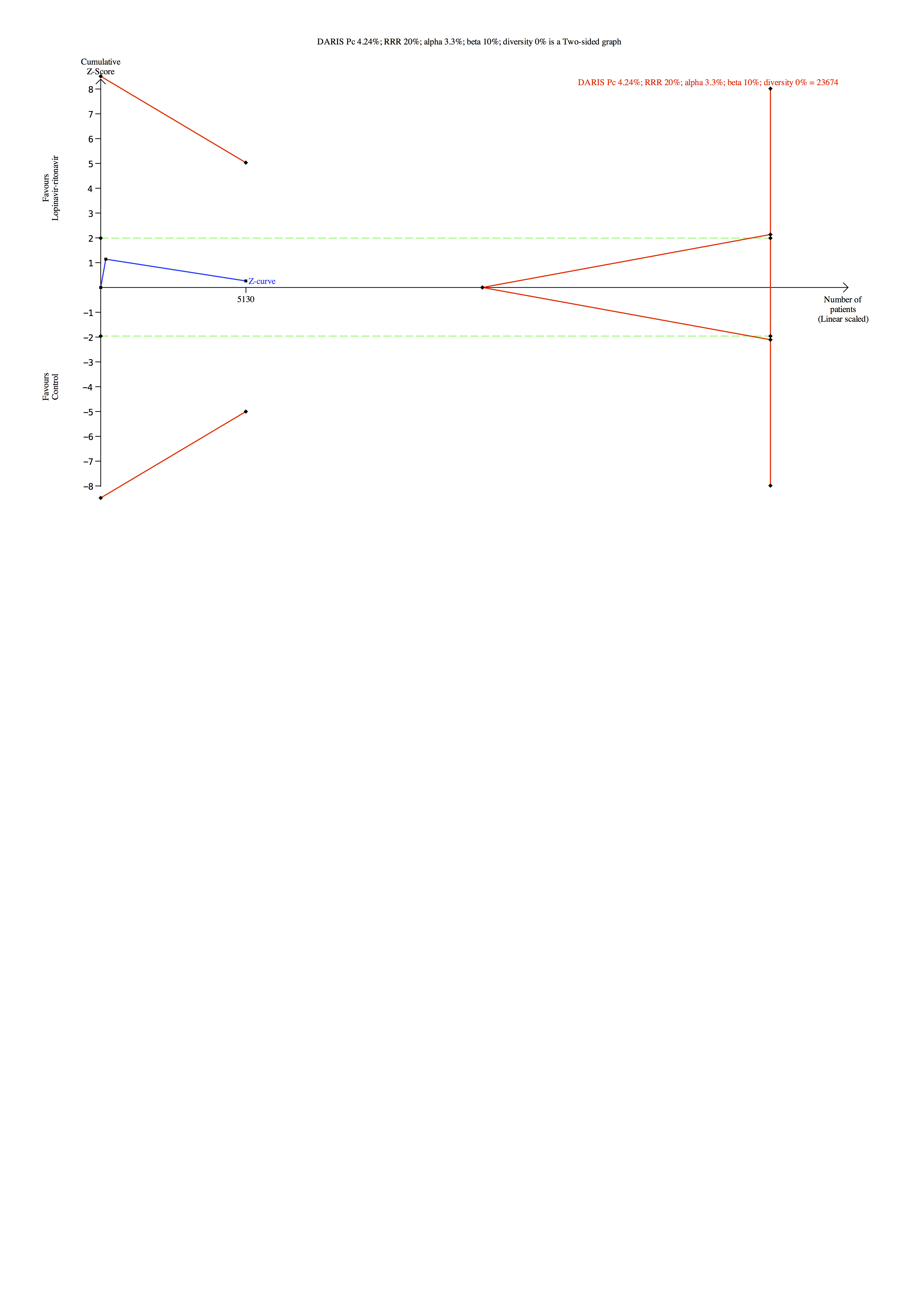

### S34 Fig.tiff

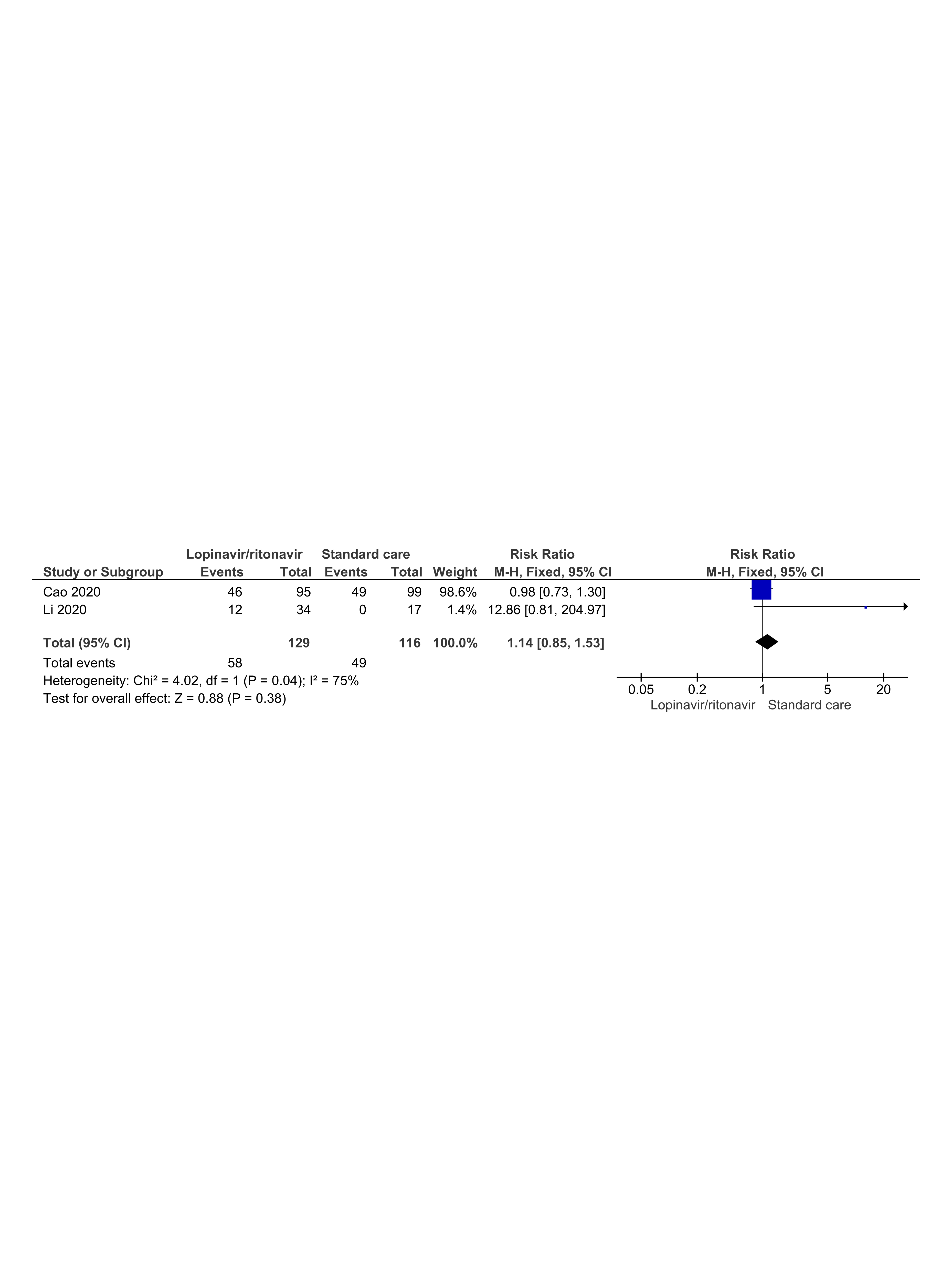

### S35 Fig.tiff

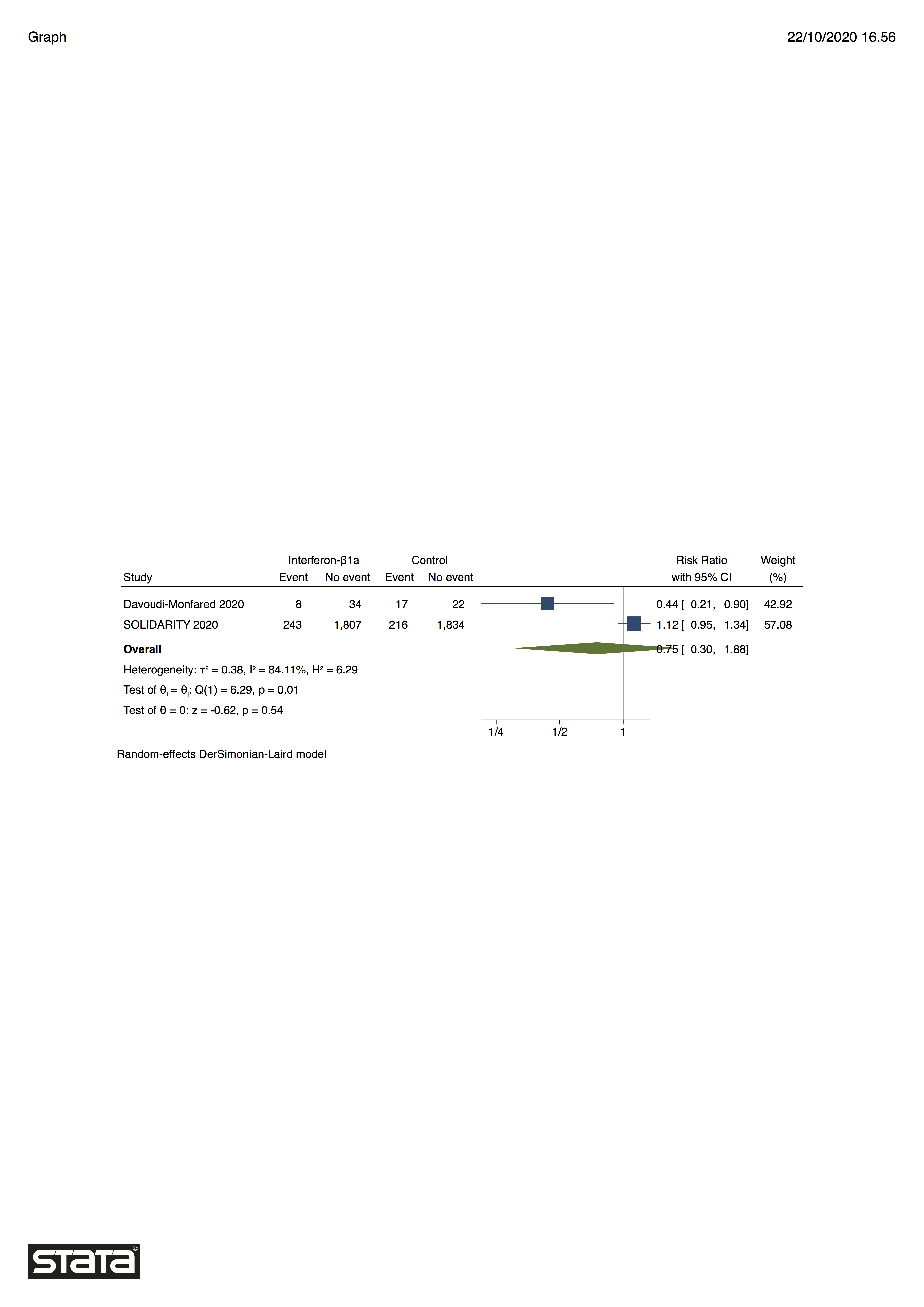

### S38 Fig.pdf

DARIS Pc (16.1)%; RRR 20%; alpha 3.3%; beta 10%; diversity (50.6)% is a Two-sided graph

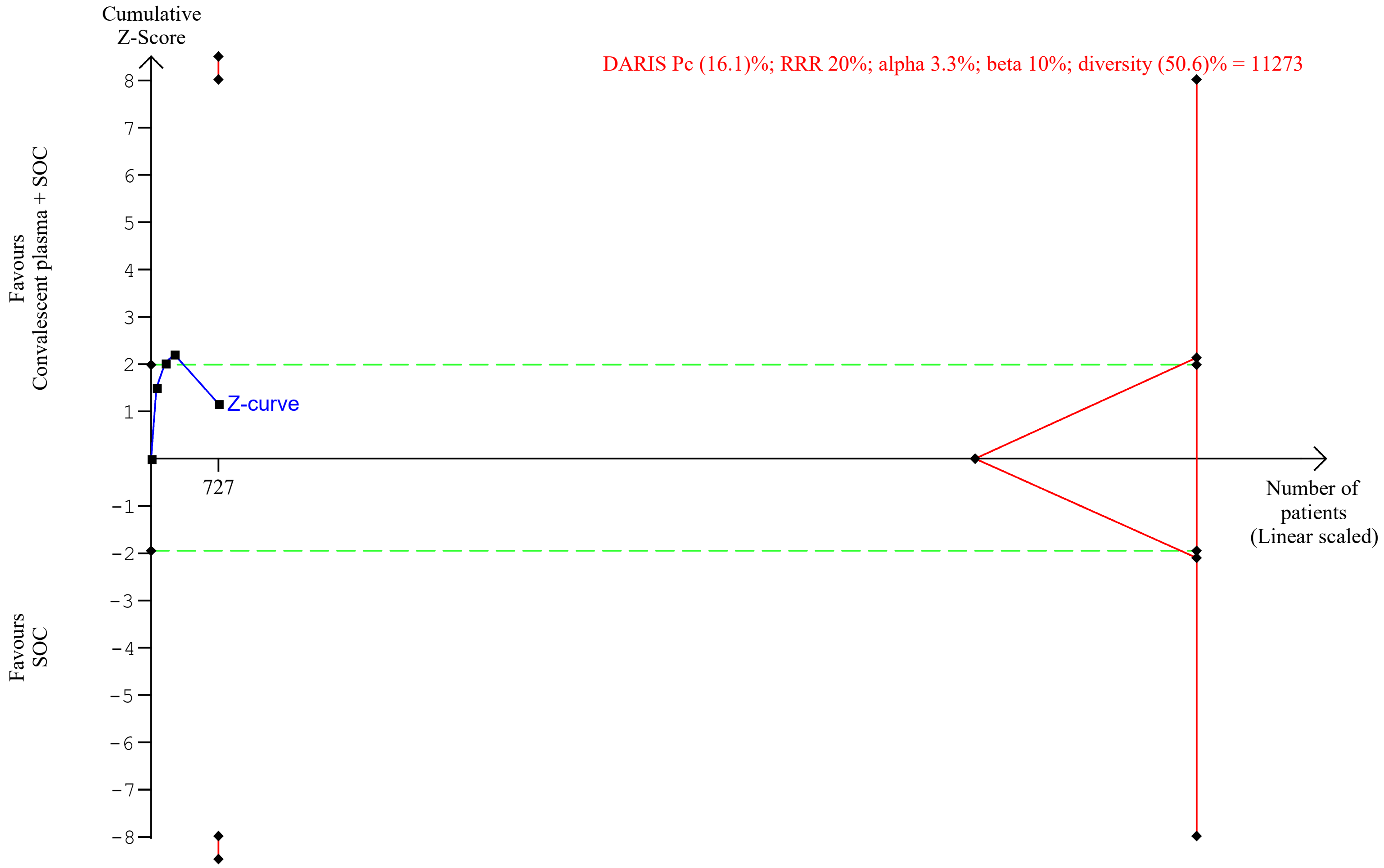

### S40 Fig.pdf

DARIS Pc (15.7)%; RRR 20%; alpha 3.3%; beta 10%; diversity (0.00)% is a Two-sided graph

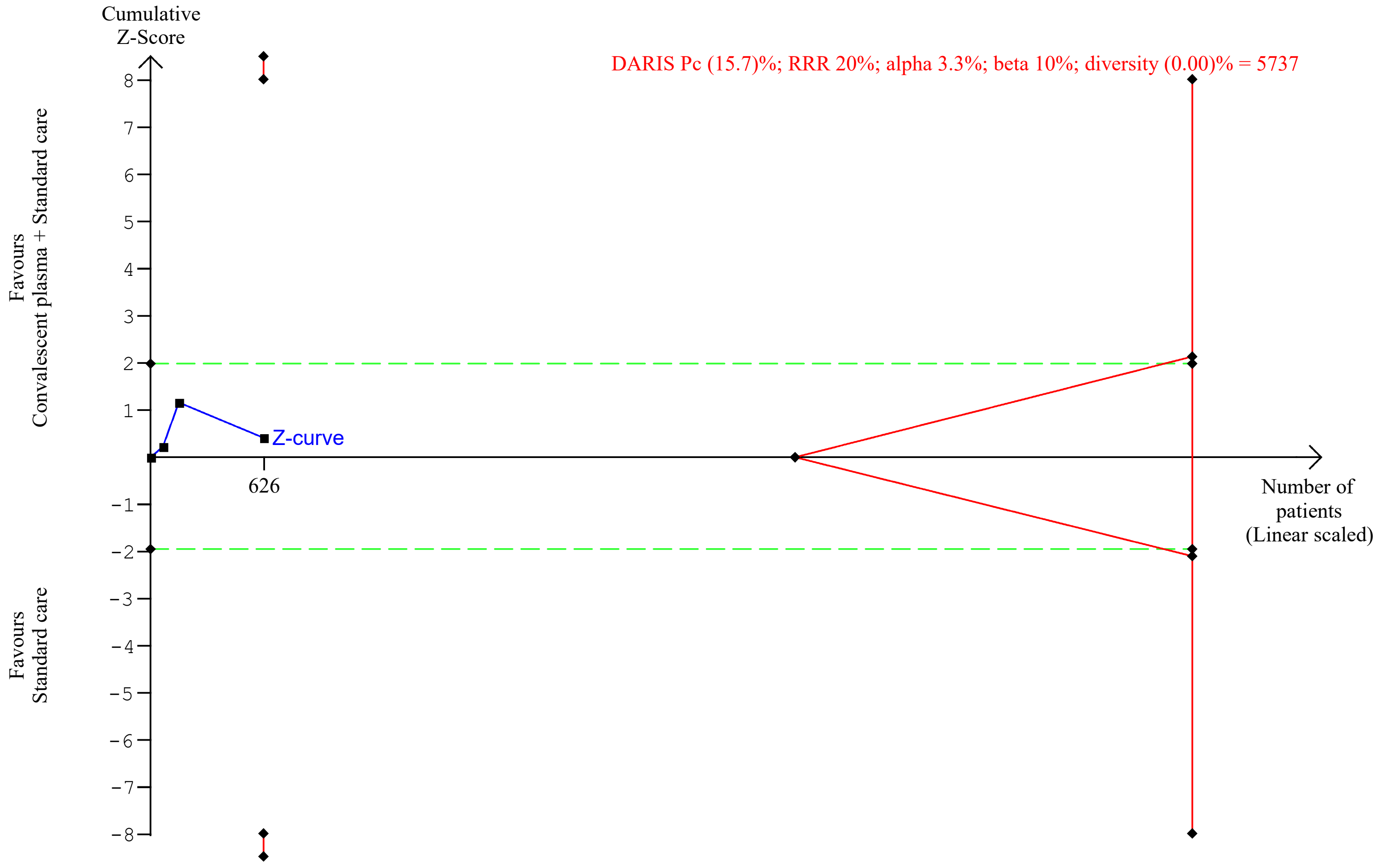

### S41 Fig.pdf

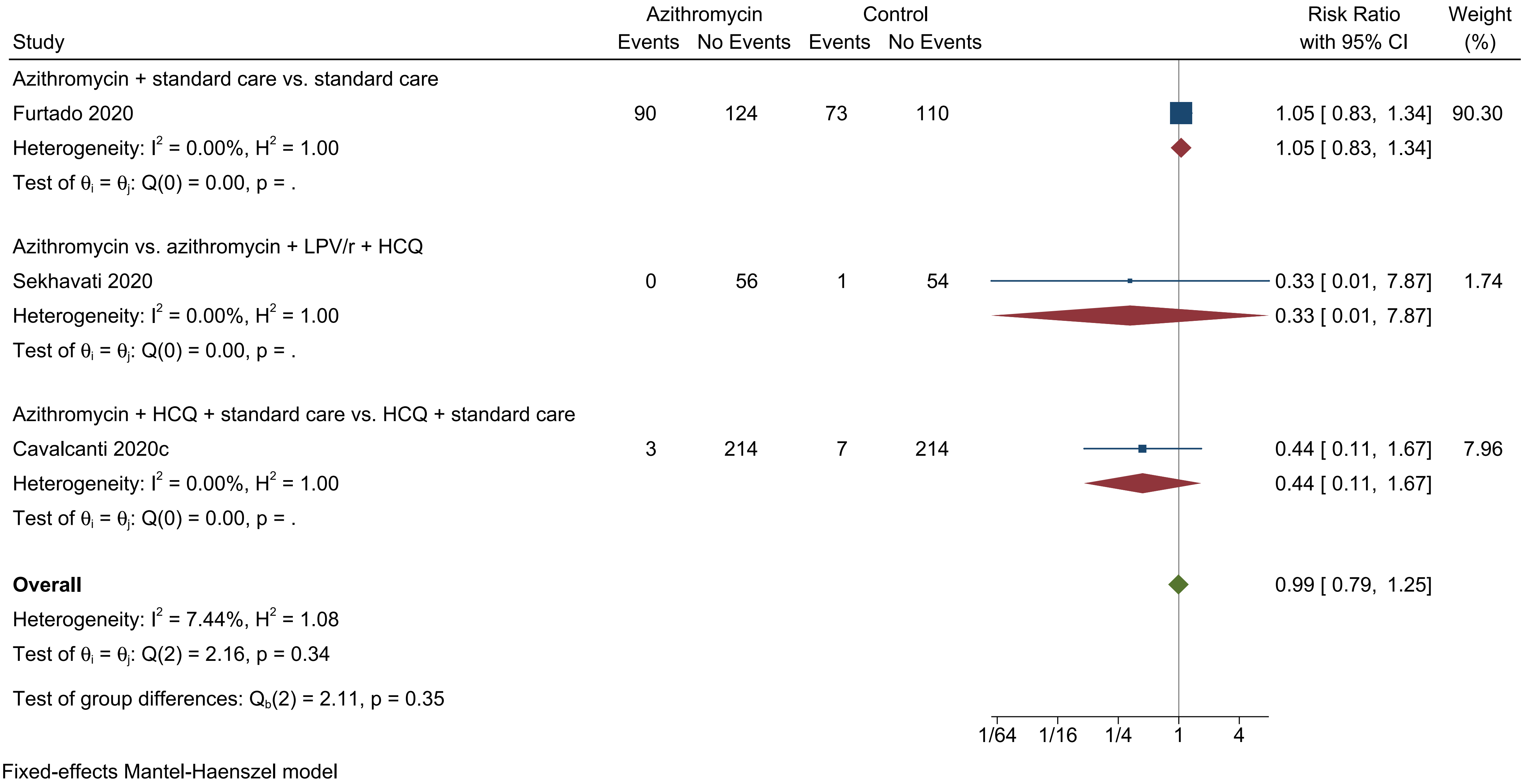

### S42 Fig.pdf

DARIS Pc 17.6%; RRR 20%; alpha 3.3%; beta 10%; diversity 67% is a Two-sided graph

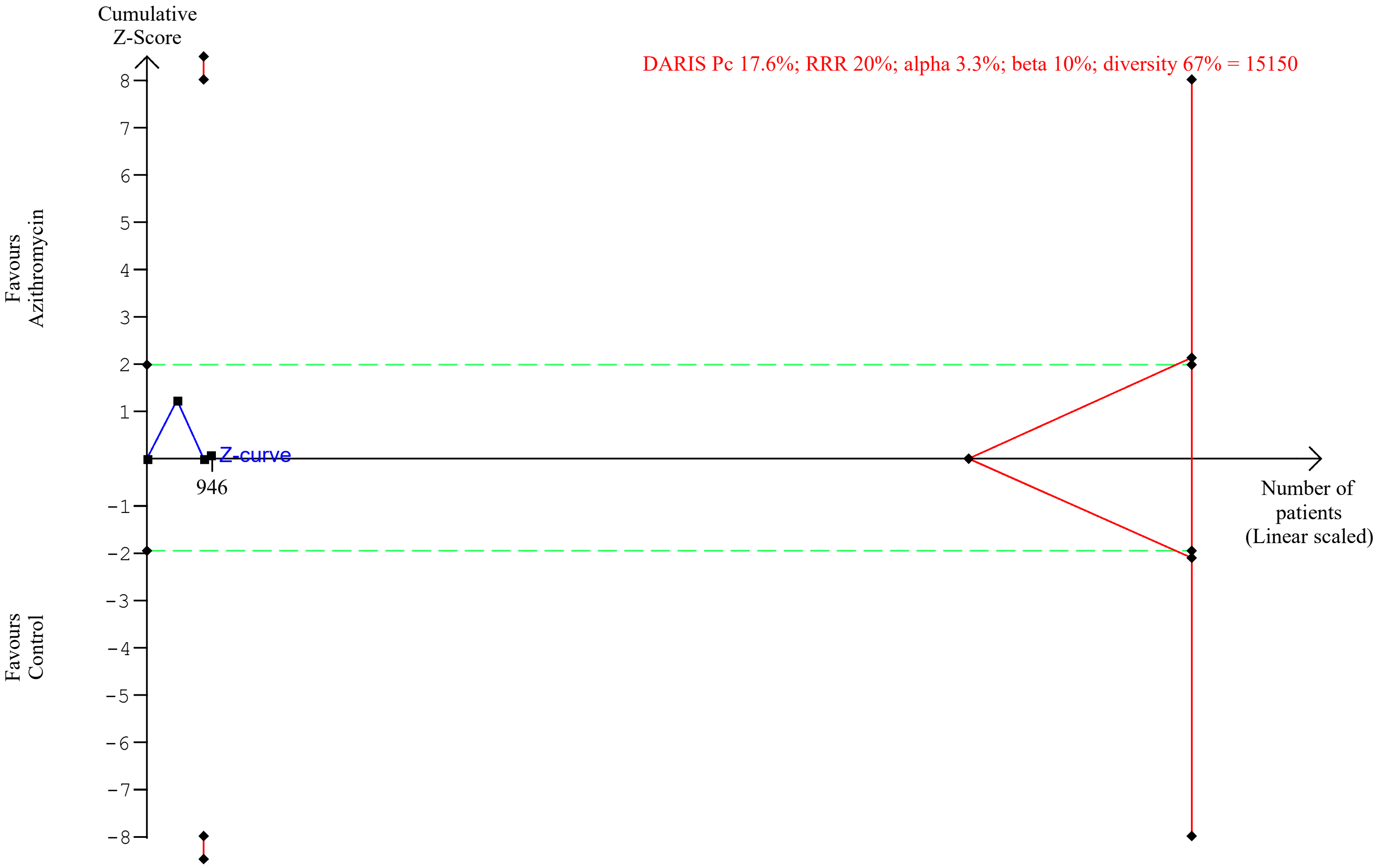

### S43 Fig.pdf

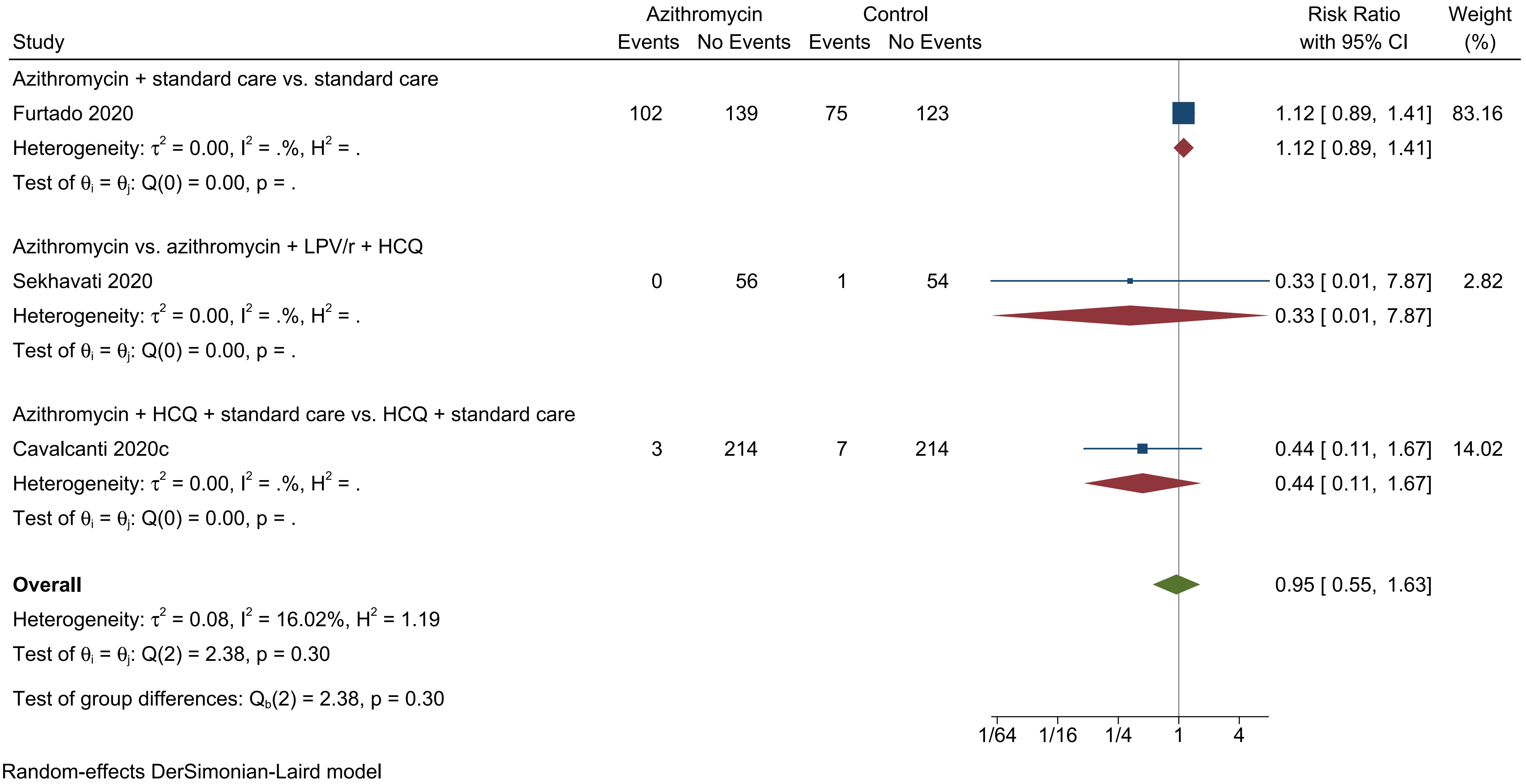

### S45 Fig.pdf

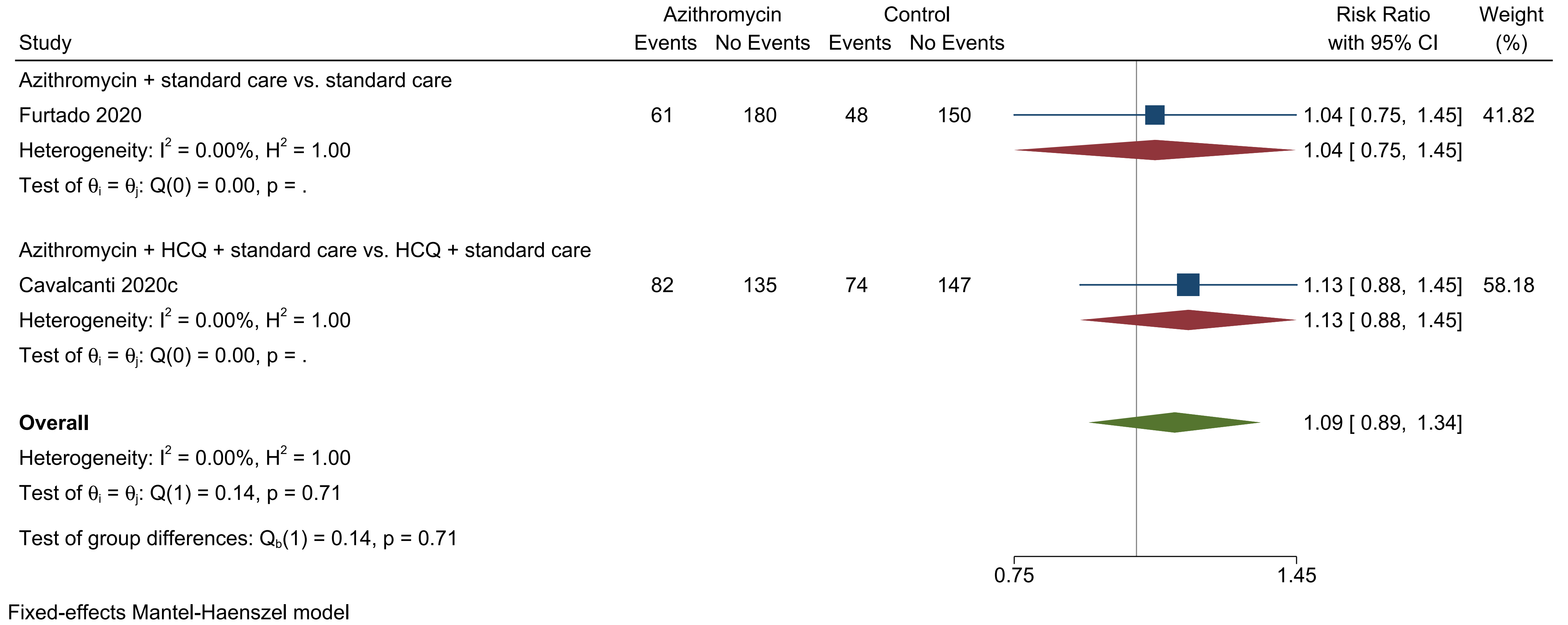

### S46 Fig.pdf

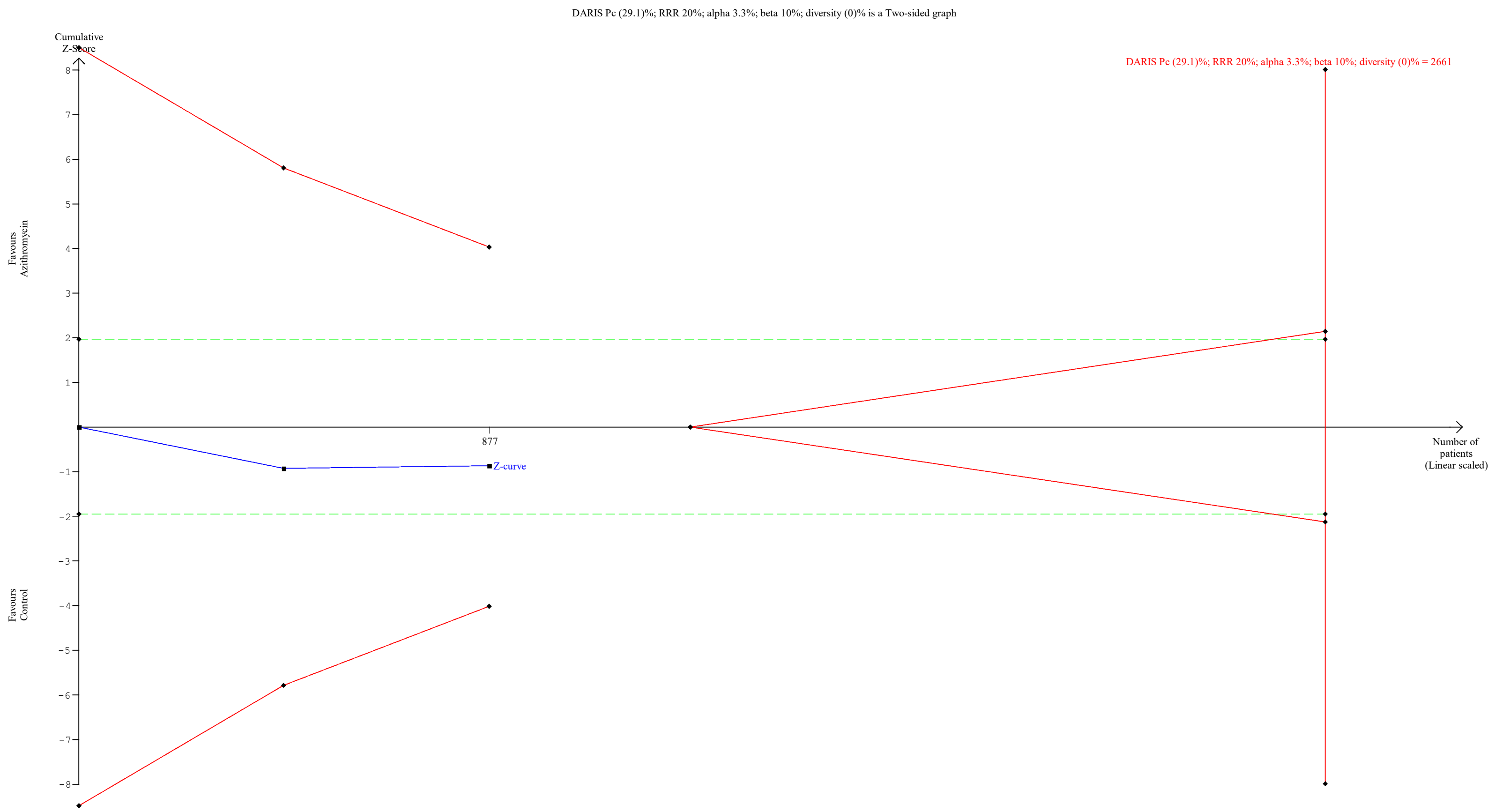

### S48 Fig.pdf

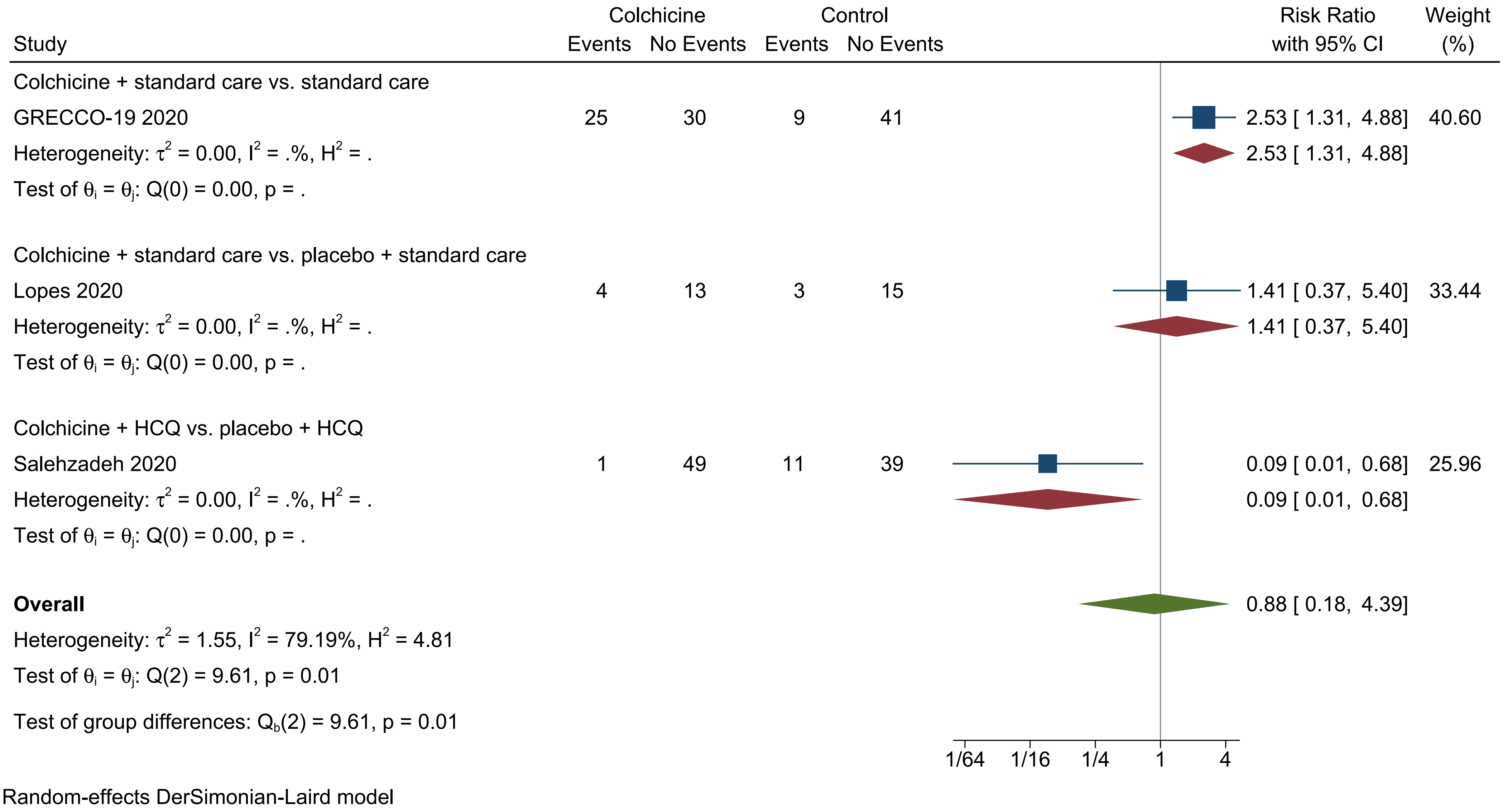

### S50 Fig.pdf

DARIS Pc 10.7%; RRR 20%; alpha 3.3%; beta 10%; diversity 62% is a Two-sided graph

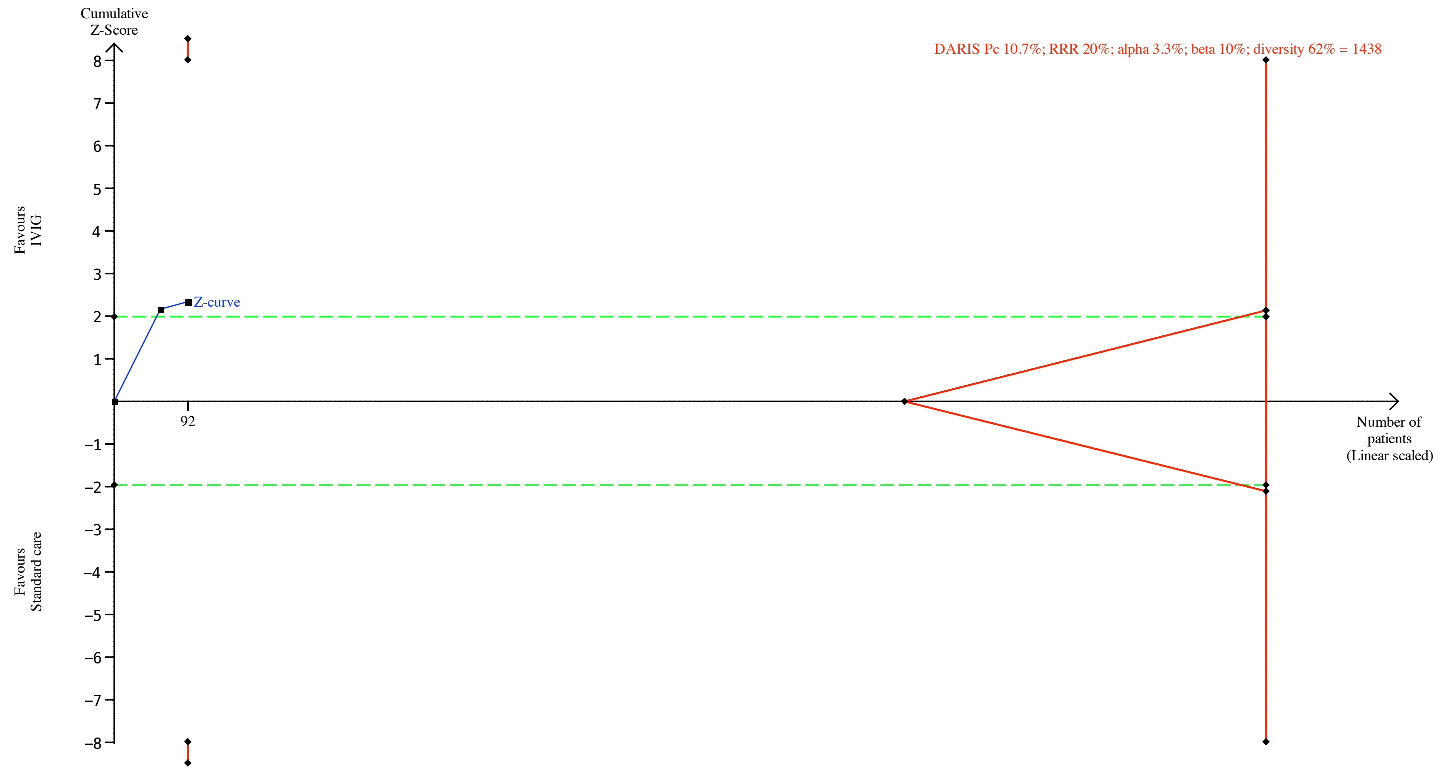

### S53 Fig.tiff

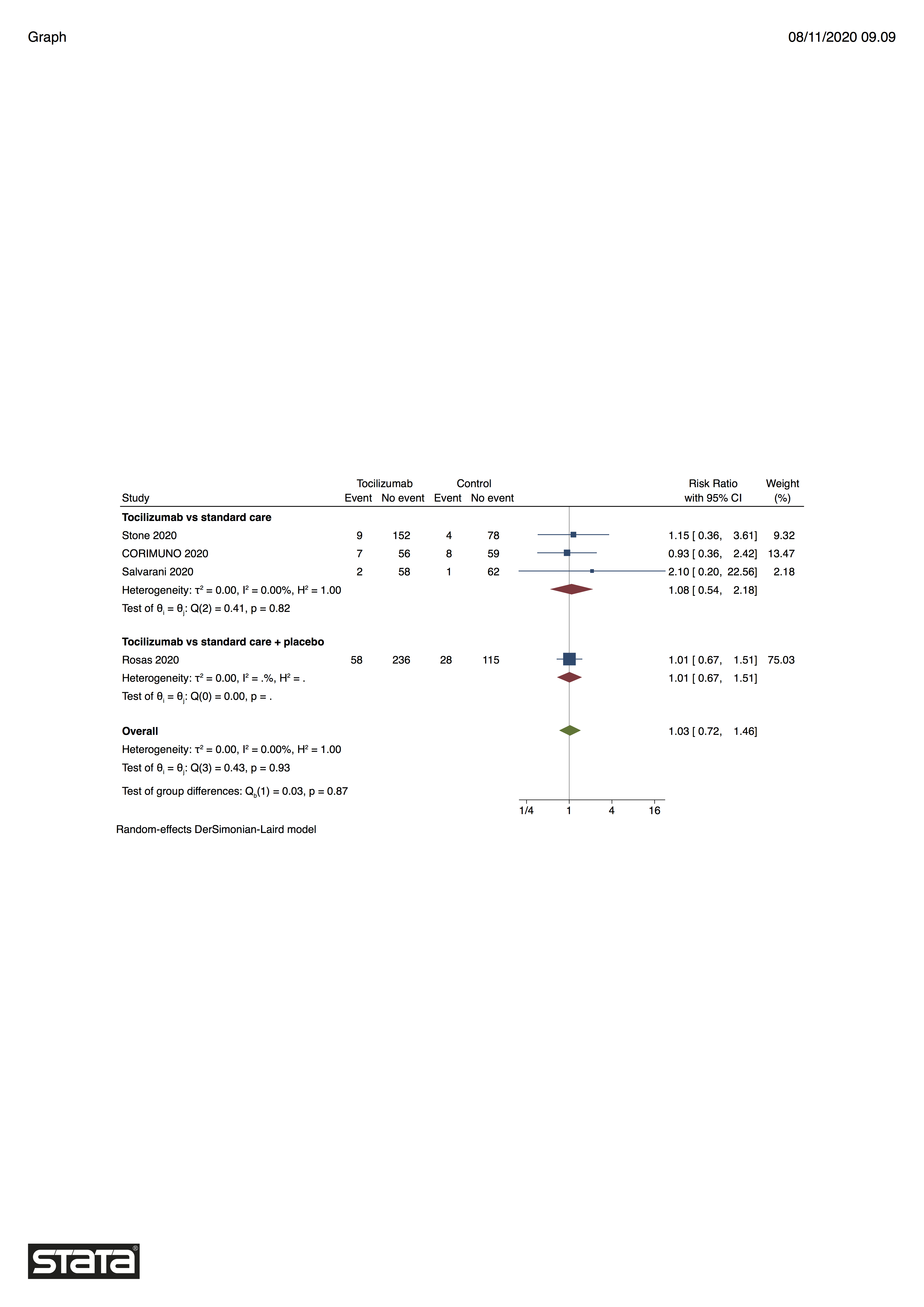

### S54 Fig.tiff

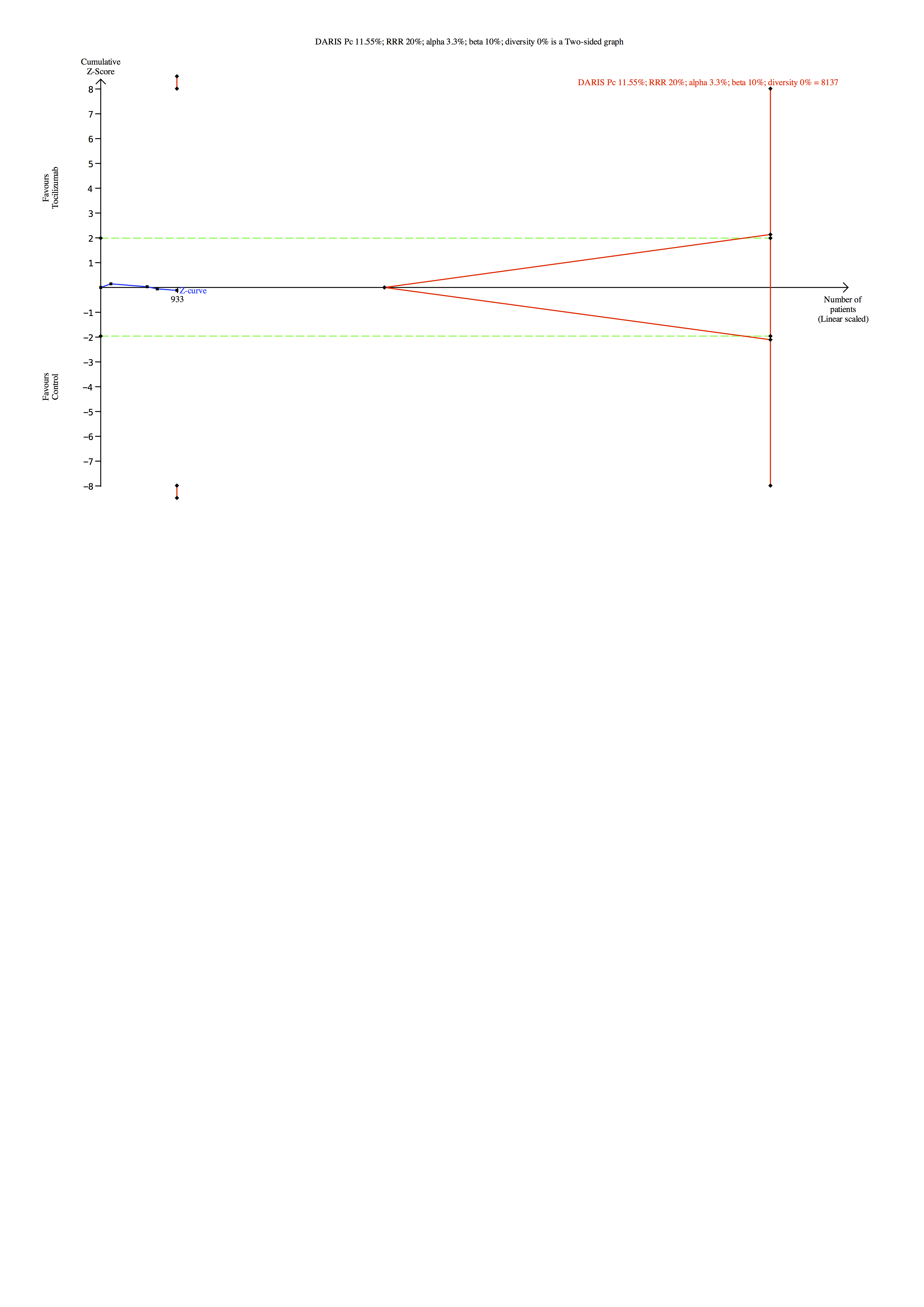

### S55 Fig.tiff

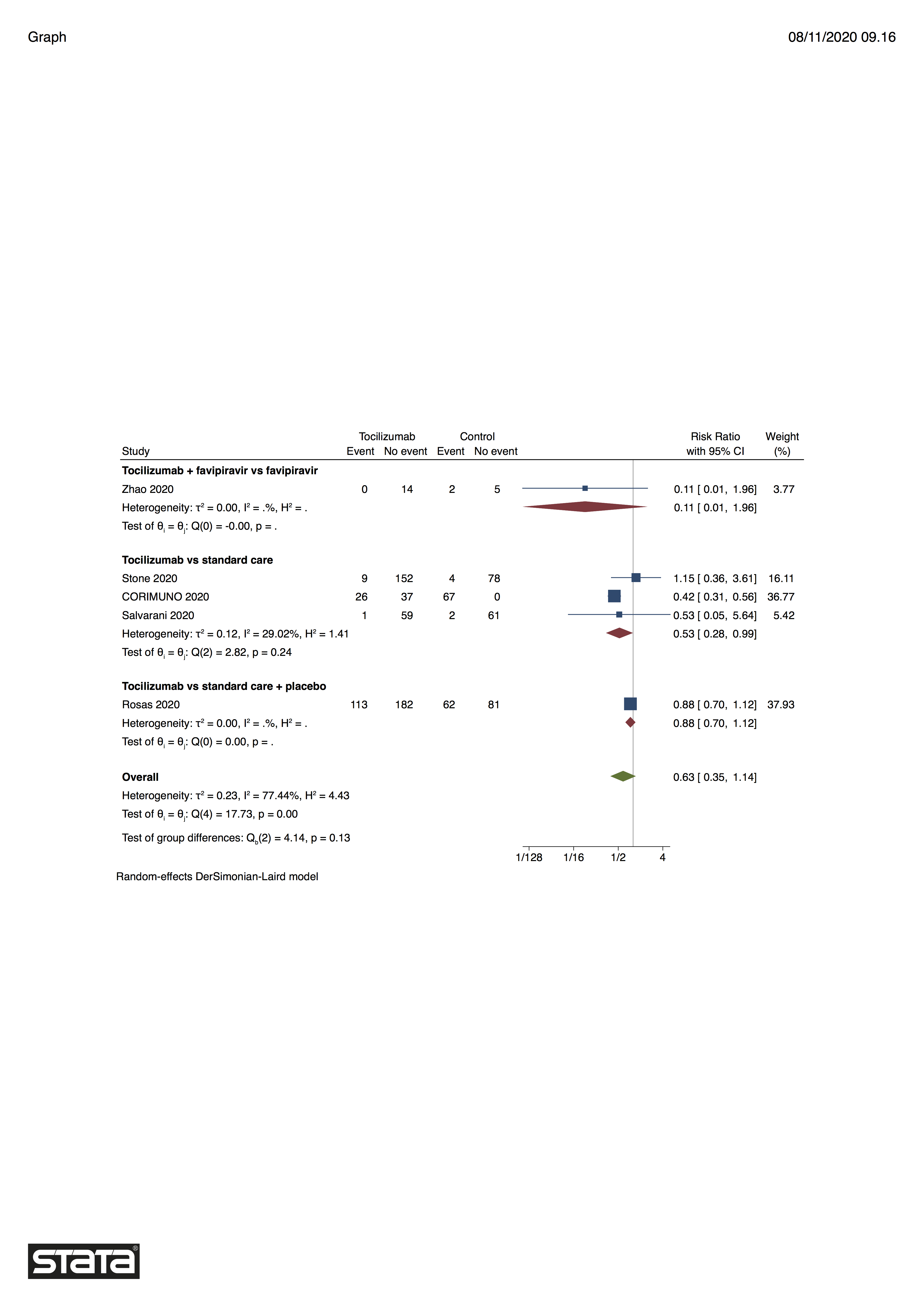

### S56 Fig.tiff

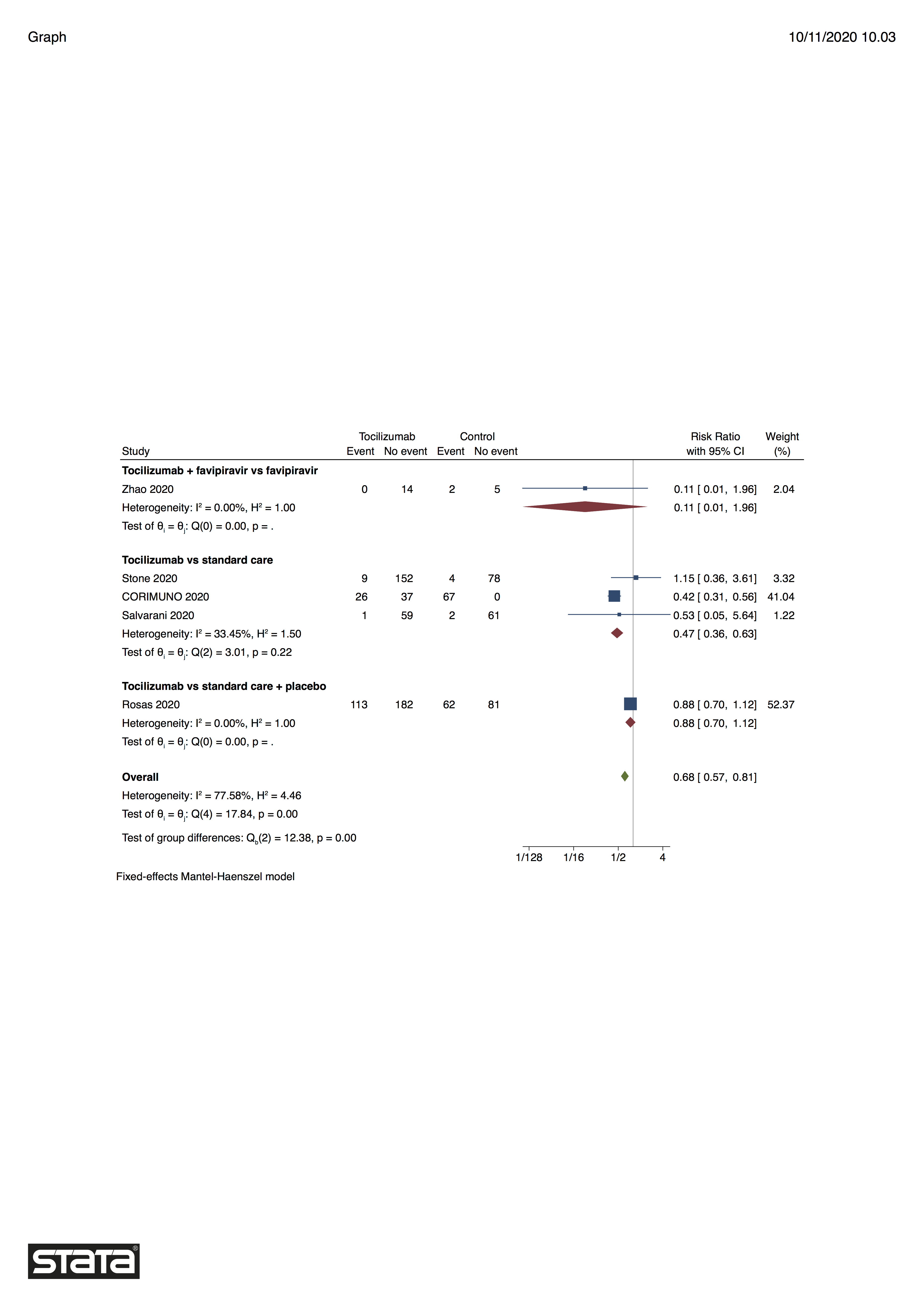

### S57 Fig.tiff

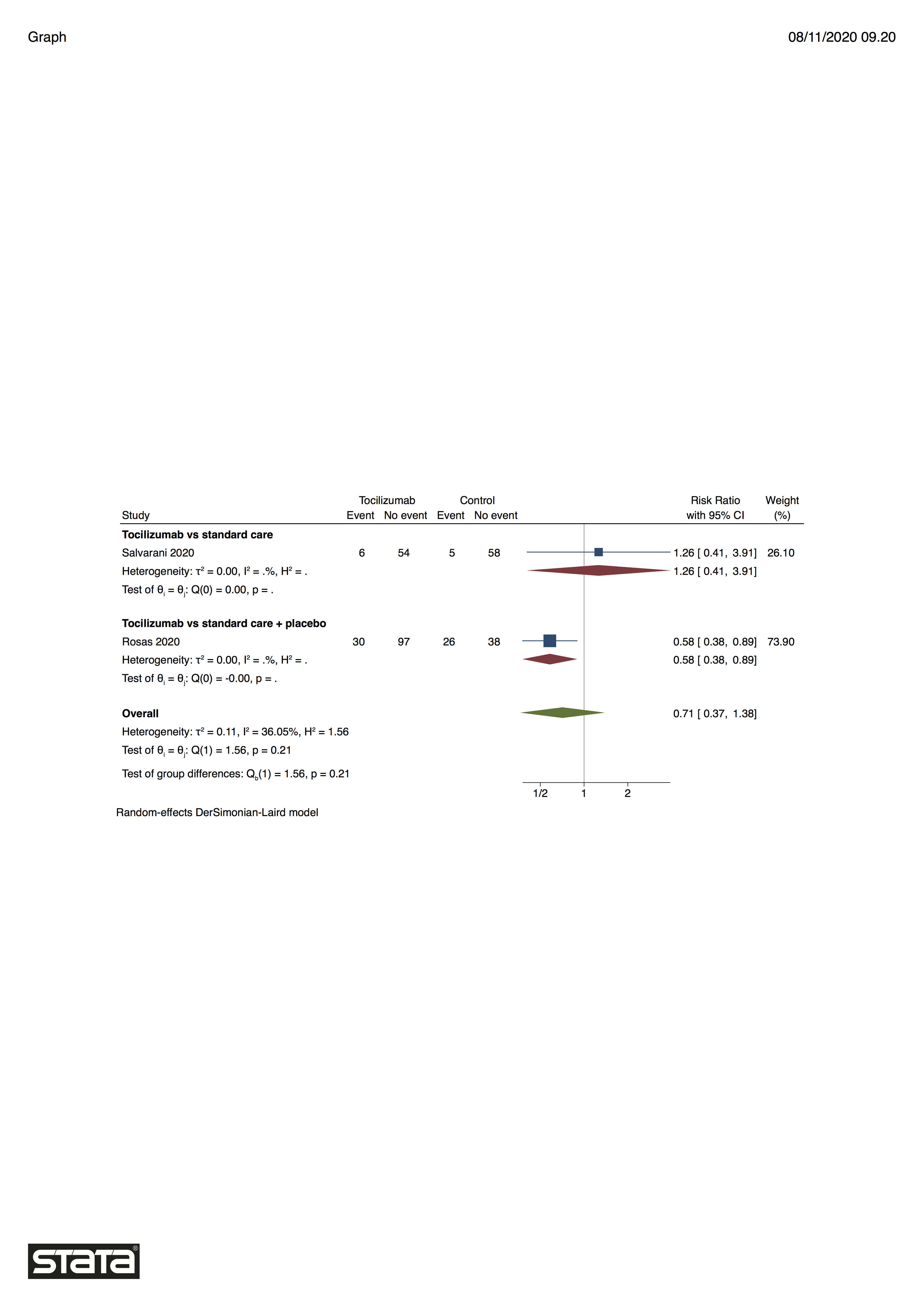

### S58 Fig.tiff

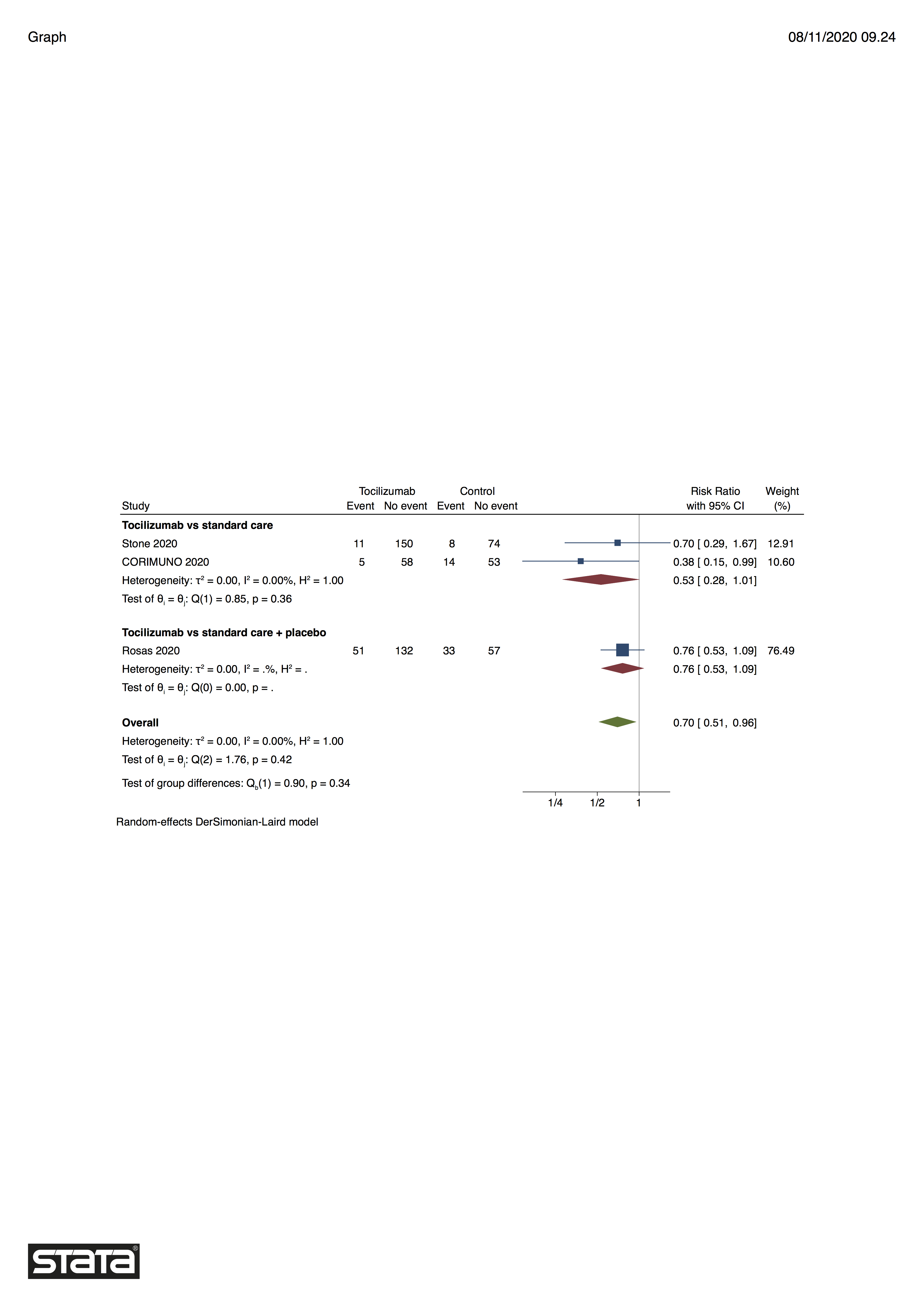

### S59 Fig.tiff

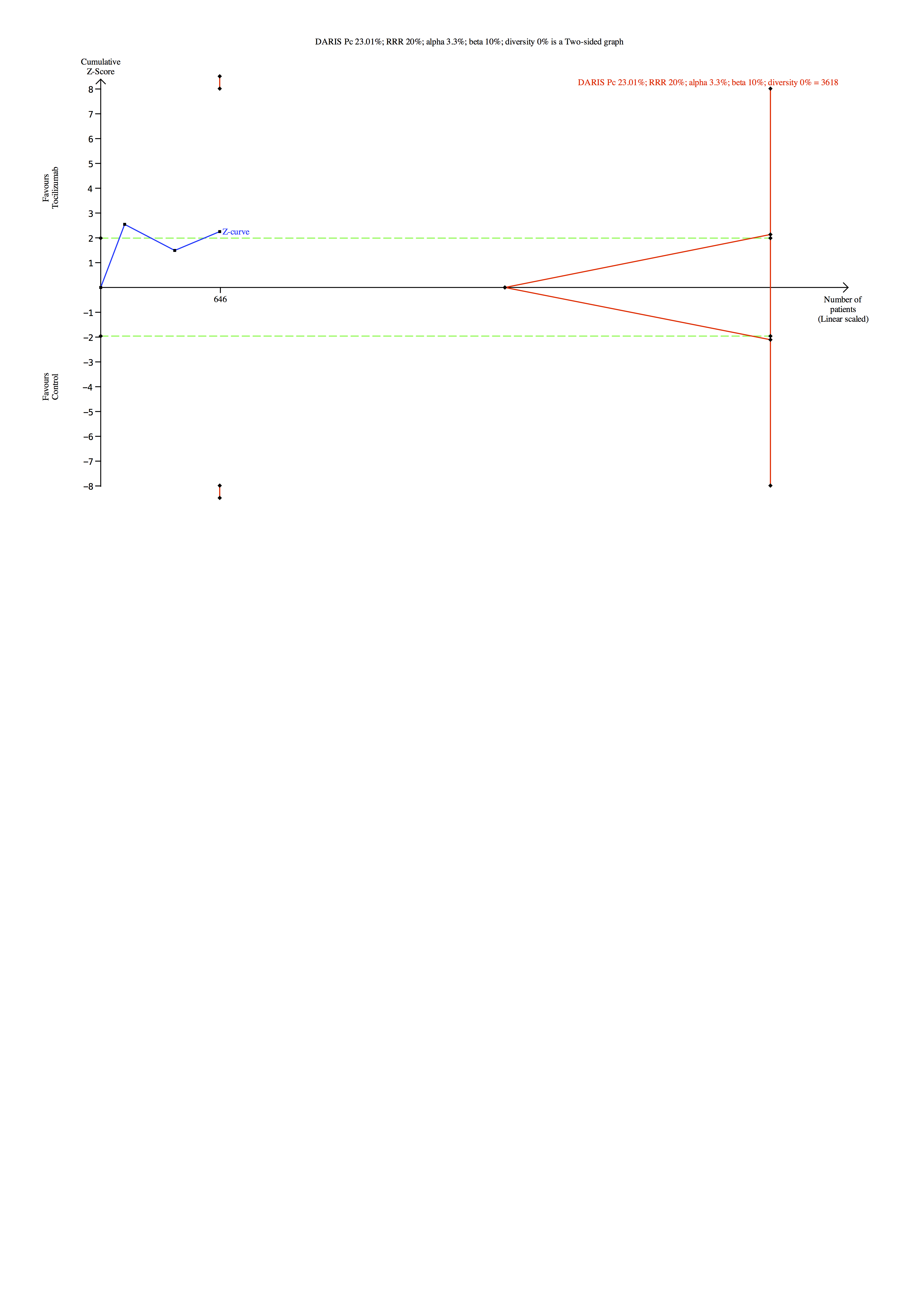

### S60 Fig.tiff

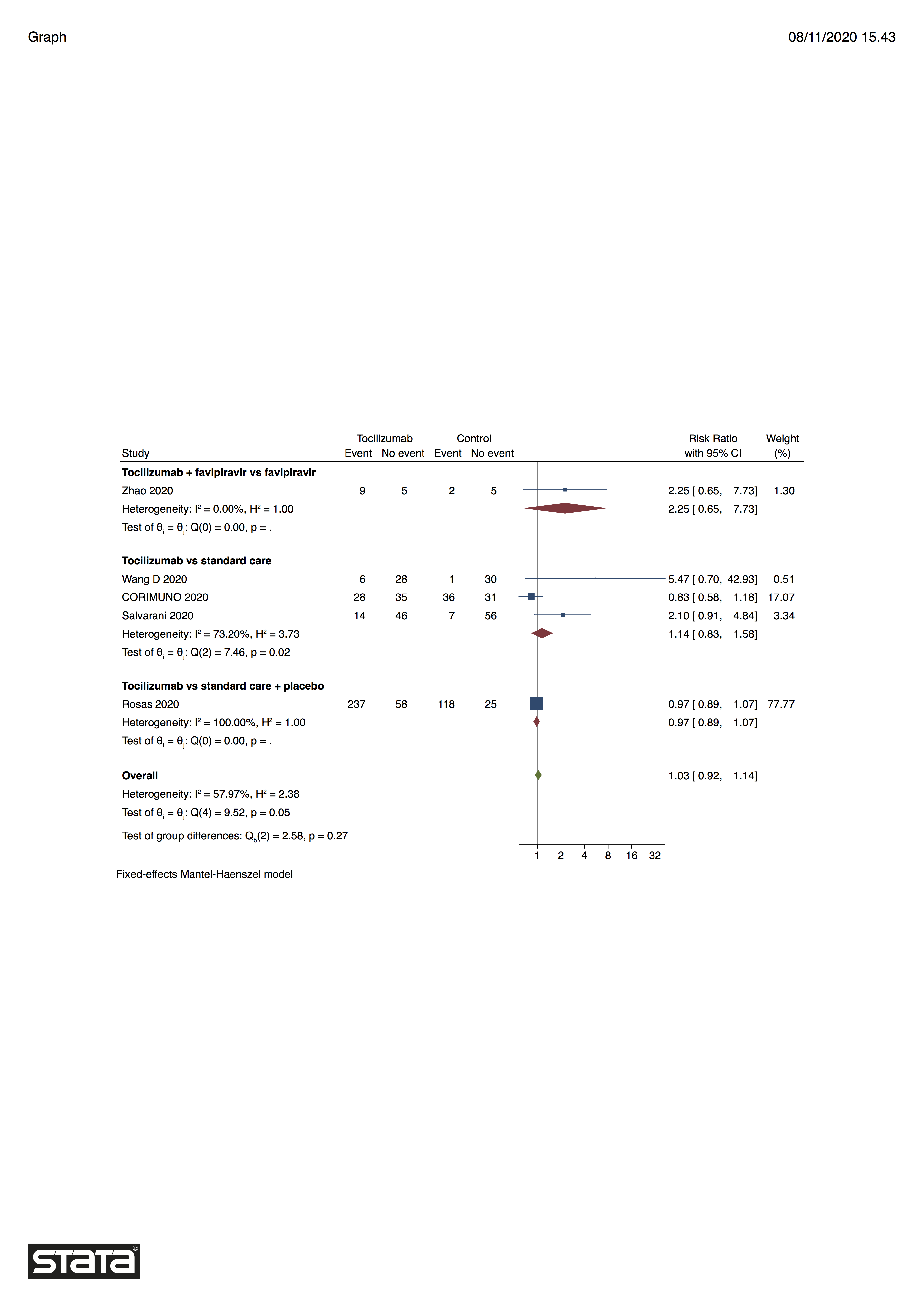
